# Comparative effectiveness of heterologous booster schedules with AZD1222, BNT162b2, or mRNA-1273 vaccines against COVID-19 during omicron predominance in the Nordic countries

**DOI:** 10.1101/2022.11.24.22282651

**Authors:** Niklas Worm Andersson, Emilia Myrup Thiesson, Ulrike Baum, Nicklas Pihlström, Jostein Starrfelt, Kristýna Faksová, Eero Poukka, Lars Christian Lund, Christian Holm Hansen, Mia Aakjær, Jesper Kjær, Catherine Cohet, Mathijs Goossens, Morten Andersen, Jesper Hallas, Hinta Meijerink, Rickard Ljung, Anders Hviid

## Abstract

**Objective:** To investigate the effectiveness of heterologous booster schedules with AZD1222 (Oxford-AstraZeneca, referred to as AZD), BNT162b2 (Pfizer-BioNTech, BNT), and mRNA-1273 (Moderna, MOD) vaccines compared with primary schedules and with homologous mRNA-vaccine booster schedules during a period of omicron predominance.

**Design:** Population-based cohort analyses.

**Setting:** Denmark, Finland, Norway, and Sweden, 27 December 2020 to 28 February 2022.

**Participants:** Adults that had received at least a primary vaccination schedule (ie, two doses) of the AZD, BNT, and/or MOD vaccines during the study period.

**Main outcome measures:** Using the Kaplan-Meier estimator, we compared country-specific risks of SARS-CoV-2 infection and severe COVID-19 outcomes in heterologous booster vaccinated with primary schedule vaccinated (matched analyses) and homologous booster vaccinated (weighted analyses) since emergence of omicron.

**Results:** Heterologous booster schedules improved protection against all outcomes compared with primary schedules, with the largest and most robust effects observed for severe COVID-19. Risk differences for documented infection ranged from -22.4% to -3.1% (comparative vaccine effectiveness [CVE] 9.7% to 60.9%; >63.2% for COVID-19 hospitalisation) across countries for AZD1BNT2BNT3 (AZD as primary dose followed by two doses of BNT) vs AZD1BNT2 and -22.2% to -3.2% (CVE 37.4% to 67.8%; >34.6% for hospitalisation) for BNT1BNT2MOD3 vs BNT1BNT2, the two most common heterologous booster schedules. Heterologous- and homologous booster schedules had comparable effectiveness. Risk differences of documented infection ranged from -0.4% to 4.4% (CVE -20.0% to 2.4%) for AZD1BNT2BNT3 vs BNT1BNT2BNT3 and -19.8% to 1.7% (CVE -14.6% to 53.8%) for BNT1BNT2MOD3 vs BNT1BNT2BNT3; for most comparisons, risk differences for severe COVID-19 outcomes were smaller than 1 per 1000 vaccinated. Previous infection followed by a booster dose conferred the greatest protection.

**Conclusion:** Heterologous booster vaccine schedules are associated with an increased protection against omicron-related COVID-19 outcomes that is comparable to that afforded by homologous booster schedules.

## INTRODUCTION

COVID-19 vaccine-induced immunity against SARS-CoV-2 infection wanes over time.[1] Together with the continued emergence of variants of concern, such as the omicron variant, managing the pandemic through immunisation is a continuously moving target.[2]

Booster doses are recommended to improve immunity in the population, and observational studies have provided evidence of the effectiveness of booster doses where the vaccine used for the booster (ie, the third dose) match the primary vaccination schedule (homologous schedules).[3–5] Additionally, clinical data have shown that use of heterologous vaccination schedules, that is, use of different COVID-19 vaccines for the primary vaccination or booster dose schedules, is just as immunogenic as homologous schedules.[6–9] How these immunologic findings translate into effectiveness is less certain.[10–16] The use of heterologous booster strategies facilitates a more flexible approach for vaccination roll-out and simplifies logistics, but should be supported by evidence of effectiveness including protection against the omicron variant of concern. This evidence is currently lacking.[17–19]

We conducted population-based cohort analyses in Denmark, Finland, Norway, and Sweden. We evaluated the comparative effectiveness of heterologous booster schedules with AZD1222 (Vaxzeria; Oxford-AstraZeneca, referred to as AZD), BNT162b2 (Comirnaty; Pfizer-BioNTech, BNT), and mRNA-1273 (Spikevax; Moderna, MOD) against SARS-CoV-2 infection and severe COVID-19 outcomes during omicron predominance in the Nordic countries. This included comparisons between 1) heterologous booster- and primary course schedules, and 2) comparisons between heterologous- and homologous booster schedules.

## METHODS

### Study design and cohort

Based on nationwide demography- and health care registers with individual-level information on vaccination status, COVID-19-related outcomes, and relevant covariates (such as age, sex, and comorbidity), we constructed country-specific cohorts including all individuals aged 18 years or older (at first vaccination) with known residency within the specific country who had received at least a primary schedule with the AZD, BNT, and/or MOD vaccines during the study period Dec 27, 2020 to Feb 28, 2022. Details on the utilised registers and variables are provided in Supplementary Material (pp 4-12). Individuals were classified according to the vaccine schedule received, and heterologous booster schedules were defined as schedules where not all three vaccine doses matched (eg, AZD as priming dose, followed by BNT as second and booster [third] dose [referred to as AZD1BNT2BNT3] or MOD1MOD2BNT3). For the primary analyses, we excluded individuals with previous documented SARS-CoV-2 infection. Also, heterologous booster schedules with use of all three different vaccines were not analysed as these were rare.

The study was conducted according to ethical and legal requirements of each country (Supplementary p 13).

#### Booster- vs primary course schedule

We assessed the effectiveness of receiving a heterologous booster- as compared with primary vaccine schedule by use of a matched design.[4] On the day an individual received a booster dose, the individual was matched with an individual who had received the same primary schedule but had not yet received a booster dose (ie, unexposed up until this date). The day the booster dose was administered served as the index date for each distinct matched pair. We additionally matched individuals on the calendar month they had received their second vaccine dose, year of birth (in 5-year bins), and a propensity score that included sex, region of residence, vaccination priority group, and selected comorbidities as predictors for receiving the respective studied heterologous booster schedule by logistic regression (Supplementary pp 7-12). Matched unexposed individuals that received a booster dose later than the assigned index date were allowed to re-enter as booster-exposed individuals in additional matched pairs on that given date.

#### Heterologous- vs homologous booster schedules

We assessed the comparative effectiveness of heterologous- and homologous booster schedules by use of inverse probability weights that included: calendar month of the index date, year of birth (in 5-year bins), sex, region of residence, vaccination priority group, and selected comorbidities. The index date was defined by the day of receiving the booster dose. To ensure that the comparison groups were of similar age and booster vaccinated within the same calendar period, we used the interval from the 2.5% to the 97.5% percentiles of the age and vaccination date distribution in the heterologous group as eligibility criteria (ie, both the heterologous and homologous vaccinated individuals had to not be younger or older than these limits and to have received their booster dose within this calendar period). We primarily compared with the larger-sized homologous BNT booster schedule; however, when the heterologous schedule did not involve the BNT vaccine, we compared with the homologous MOD booster schedule.

### Outcomes

The four COVID-19 outcomes of interest were documented SARS-CoV-2 infection (ie, positive PCR test), inpatient hospitalisation for COVID-19 (ascertained by COVID-19 related diagnoses and positive PCR tests), intensive care unit (ICU) admission for COVID-19, and COVID-19 related death (Supplementary pp 10-12). Follow-up was conducted separately for each outcome.

### Statistical analysis

Follow-up began on day 14 after the index date (to ensure full immunisation) and ended on the day of the outcome, day 75 after the start of follow-up (ie, approximately 3 months since the index date), death, emigration, or end of study, whichever occurred first. Specifically for the analyses comparing matched booster- vs primary schedules, we also censored matched pairs if the unexposed individual received a booster dose during follow-up.[4] For the analyses of COVID-19 hospitalisation and ICU admission, we censored individuals with a positive PCR test for these outcomes on day 14 hereafter and after day 30 for COVID-19 related death. The period of omicron predominance was based on national surveillance data and defined as the period when the omicron variant (sublineages BA.1 and BA.2) accounted for more than 90% of all cases: as starting on Dec 28, 2021 in Denmark and Norway, Jan 1, 2022 in Finland, and Jan 3, 2022 in Sweden and until end of study period (Feb 28, 2022). Individuals with follow-up time during these respective calendar periods contributed to the estimation of the cumulative incidences using the Kaplan-Meier estimator according to schedule in the matched analysis or the weighted analysis. Absolute and relative (ie, comparative vaccine effectiveness [CVE]; calculated as 1 – risk ratio) differences were estimated using the cumulative incidences at day 75. We calculated the corresponding 95% confidence intervals (CIs) using the delta method. We truncated the upper 95% CI of the CVE if it exceeded 100% (a computational consequence in some comparisons owed to large standard deviation when number of compared individuals and events was small). The effect estimates across countries were unsuitable for meta-analysis due to significant heterogeneity (data not shown); heterogeneity was evaluated with the Cochran Q test. We calculated the inverse probability weights as ((1-p_0_)/(1-p_c_))/(p_0_ /p_c_); p_0_ equal to the crude probability of the specific schedule and p_c_ equal to the probability of the specific schedule given covariates; these probabilities were computed using logistic regressions.

Additional analyses included comparing the effectiveness of homologous BNT and MOD booster schedules as well as vs primary schedules, and examining the risk of documented SARS-CoV-2 infection with stratification according to previous infection. For the heterologous- vs homologous booster comparisons, we did a sensitivity analysis with adjustment for calendar week of the index date (instead of month; to assess potential residual confounding of calendar time in the main analysis). Moreover, for the booster- vs primary schedule comparisons of documented infection in Denmark, we assessed whether differences in the infection rates modified the effect differences by stratifying calendar period including prior to Dec 28, 2021 (to explore whether the high infection rates in Denmark contributed to the generally observed smaller risk differences relative to the other countries).

### Patient and public involvement

No patients were formally involved in setting the research question, study design, outcome measures, or the conduct of the study.

## RESULTS

### Population

A combined total of 1,851,238 heterologous booster vaccinated individuals were included for the comparisons with primary schedules, and 3,260,410 for the comparisons with homologous booster vaccinated (Supplementary pp 14-15 for numbers of excluded individuals for each comparison owing to history of previous infection and pp 16-32 for distributions of baseline characteristics). The most commonly used heterologous booster schedule among those including AZD was AZD1BNT2BNT3 and BNT1BNT2MOD3 for those schedules with mRNA vaccines only. AZD1AZD2mRNA3 [ie, BNT3/MOD3] schedules were mainly used in Finland and Sweden and these individuals were generally older (mean age, 69 years) relative to recipients of other schedules (mean age was between 35 and 55 years). Individuals vaccinated with AZD only as first dose (ie, AZD1mRNA2mRNA3) were more likely female (≥65%) in Denmark, Norway, and Sweden, but not in Finland. Heterologous booster schedules that included AZD were generally administered earlier (the majority from November 2021 through January 2022) than those heterologous booster schedules that included only mRNA vaccines (December 2021 through February 2022); density plots of age and index date distributions are given in the Supplementary pp 33-42.

### Booster- vs primary course schedule

When compared with matched primary course vaccinated individuals, having received a heterologous booster schedule was in the majority of comparisons associated with lower risk of COVID-19 outcomes. However, estimates varied, particularly for the outcome documented SARS-CoV-2 infection (Figure 1, Figure 2, and Supplementary pp 43-44 and pp 47-48): the cumulative incidences of infection at day 75 ranged from <1% to 45% across countries and comparisons. Some country-specific comparisons did not yield enough cases for analyses and some estimates were imprecise due to low number of cases. The cumulative incidences of hospitalisation for COVID-19 were low and more similar across comparisons and countries in contrast to documented infection (Figure 1, Figure 3, and Supplementary p 45 and pp 49-50; the incidences of ICU admission and death were very low or zero). For schedules where analyses were feasible, boosters reduced the risk of severe COVID-19 outcomes compared with primary schedules. CVE against hospitalisation for AZD1BNT2BNT3 vs AZD1BNT2 was 86.6% (95% CI 65.1%-100.0%) in Finland and 63.2% (12.1%-100.0%) in Sweden (not estimable in Denmark or Norway due to too few cases). CVE against hospitalisation for BNT1BNT2MOD3 vs BNT1BNT2 was 34.6% (−85.8%-100.0%) in Finland, 94.6% (90.0%-99.3%) in Norway, and 88.7% (83.6%-93.8%) in Sweden (not estimable in Denmark). Similar effect estimates were observed for homologous boosters; for hospitalisation, CVE ranged from 77.9% to 88.7 for BNT and 76.5% to 94.4% for MOD (Supplementary pp 54-56).

**Figure 1.**
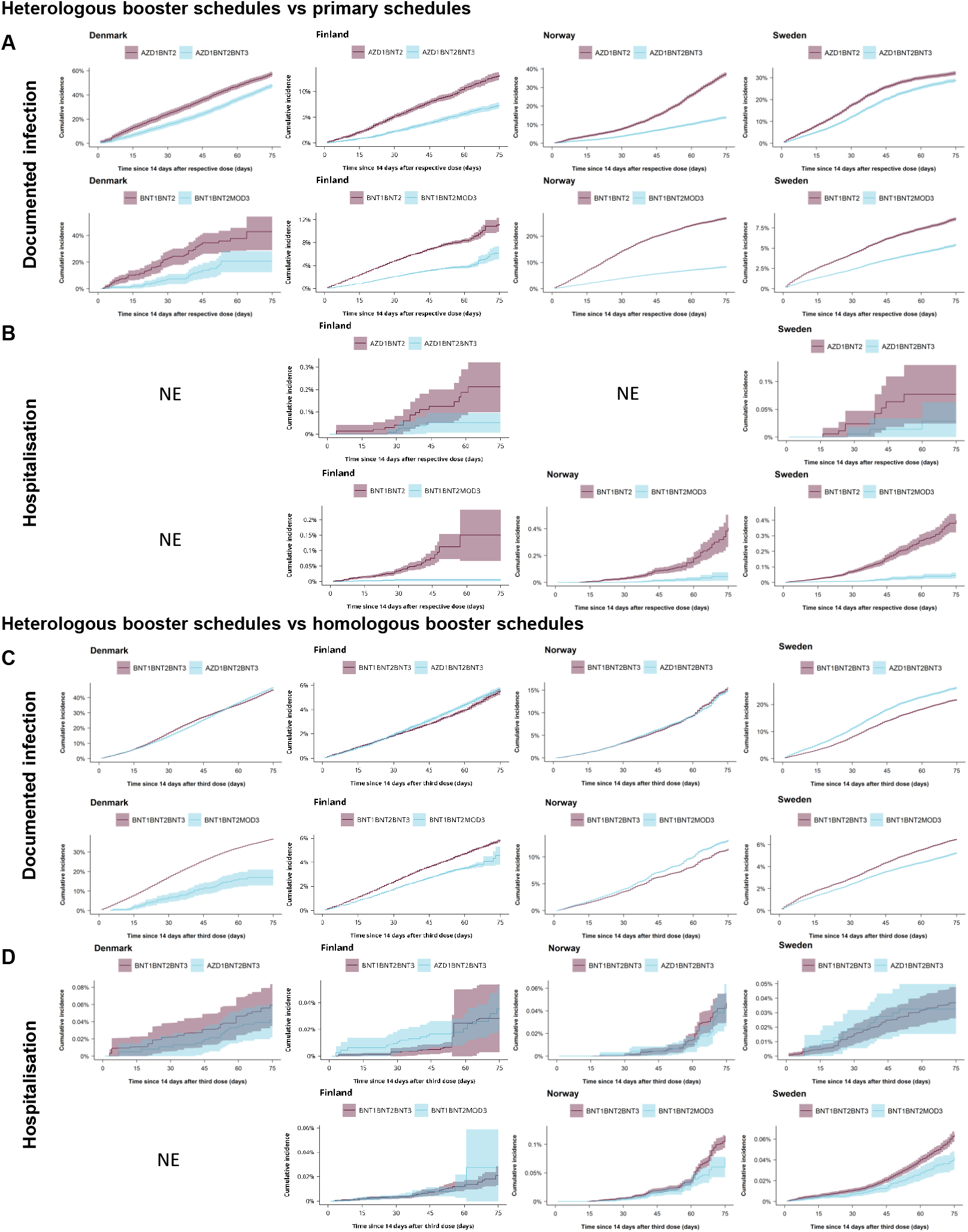
Cumulative incidence curves of documented SARS-CoV-2 infection and hospitalisation for COVID-19 comparing heterologous booster schedules commonly used in the Nordic countries with primary schedules and with homologous booster schedules in each country. Cumulative incidence curves of documented SARS-CoV-2 infection (panel A and C) and hospitalisation for COVID-19 (panel B and D) for the most commonly used heterologous AZD-based and mRNA-only-based booster schedules (blue lines; namely, AZD1BNT2BNT3 [acronym denotes first dose: AZD, second dose: BNT, and third dose: BNT] and BNT1BNT2MOD3) across the four countries (by columns) as compared with primary schedules (panel A and B; matched analyses) and with homologous booster schedules (panel C and D; weighted analyses); comparator group denoted by the red lines (the shaded areas depict the corresponding 95% confidence intervals). Cumulative incidence plots for all matched heterologous booster- vs primary schedule comparisons and all weighted heterologous- vs homologous booster schedule comparisons and including the additional severe COVID-19 outcomes are presented in Supplementary (pp 43-46 and 57-60, respectively). NE denotes not estimated meaning that the cumulative incidence curves could not be generated for this specific country (column) comparison (row).

**Figure 2.**
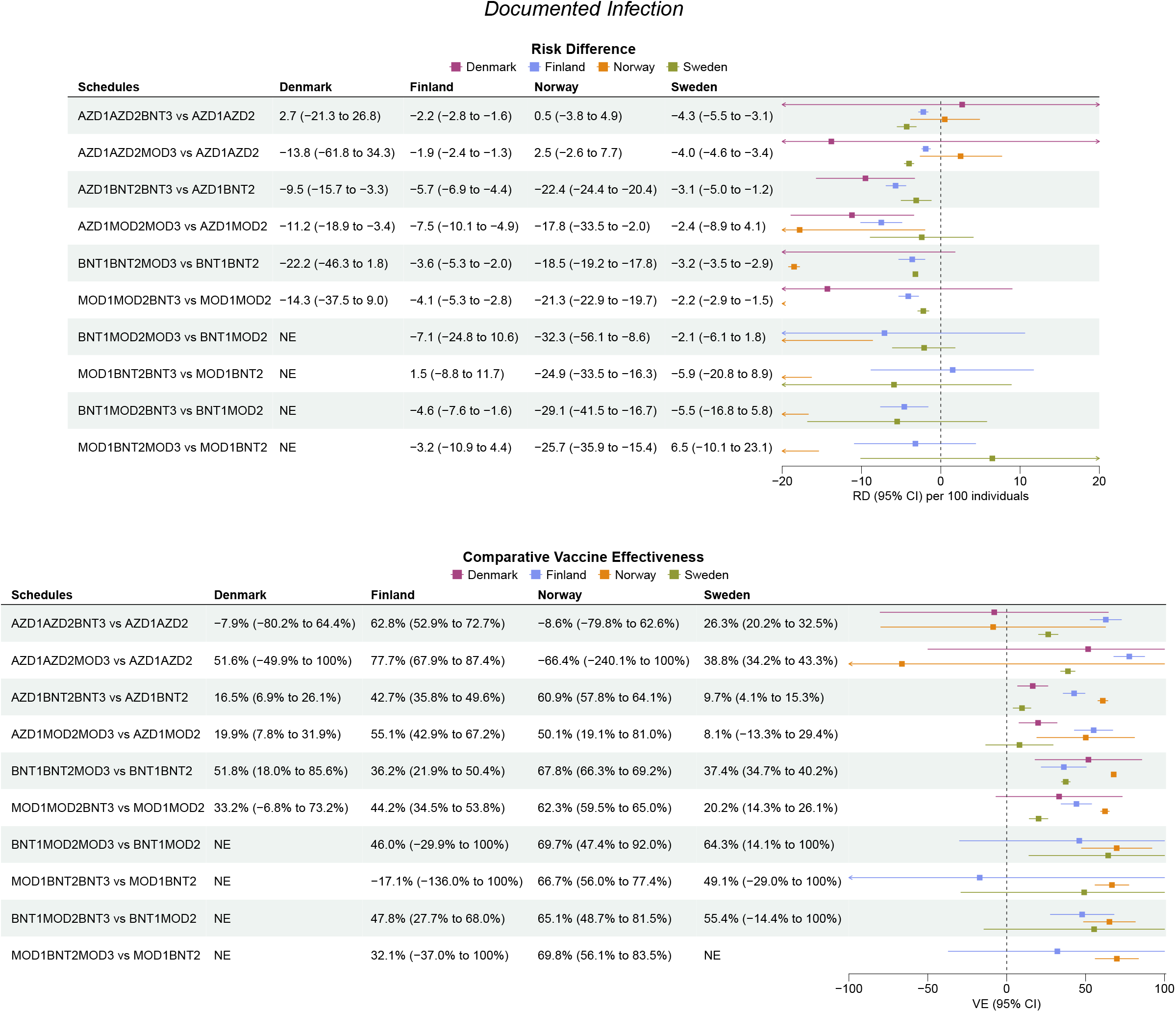
Risk of documented SARS-CoV-2 infection comparing heterologous booster-with primary schedules in each country. Risk difference per 100 individuals and comparative vaccine effectiveness (1 – risk ratio) for documented SARS-CoV-2 infection at day 75 after start of follow-up were adjusted for calendar month of receiving the second vaccine dose, year of birth (5-year bins), sex, region of residence, vaccination priority group, and selected comorbidities through a matched design. Risk estimates are presented together with number of events and total person-years within each comparison in Supplementary (pp 47-48). NE denotes not estimated meaning that the risk estimates could not be generated for this specific country comparison.

**Figure 3.**
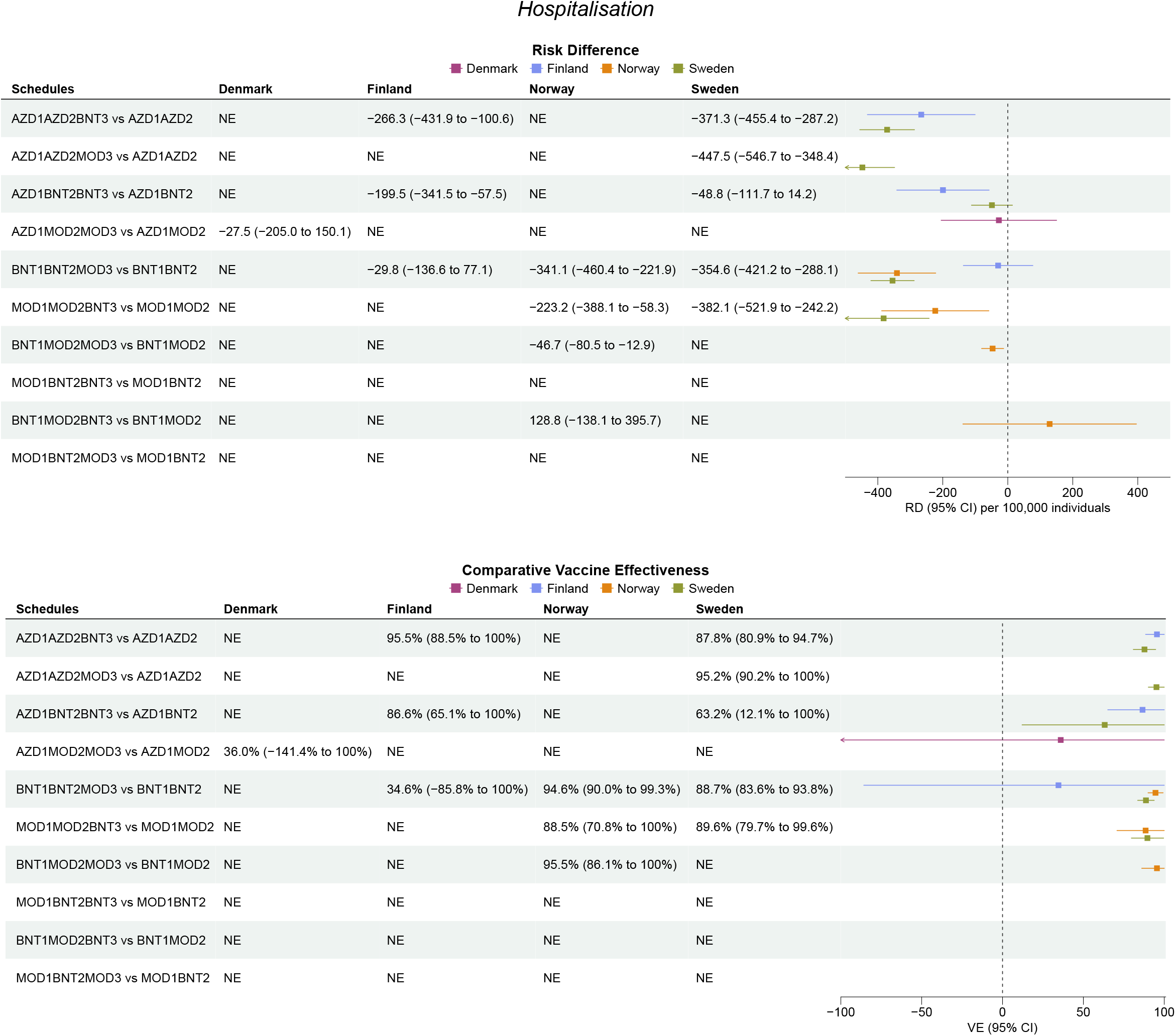
Risk of hospitalisation for COVID-19 comparing heterologous booster-with primary schedules in each country. Risk difference per 100,000 individuals and comparative vaccine effectiveness (1 – risk ratio) for hospitalisation for COVID-19 at day 75 after start of follow-up were adjusted for calendar month of receiving the second vaccine dose, year of birth (5-year bins), sex, region of residence, vaccination priority group, and selected comorbidities through a matched design. Risk estimates are presented together with number of events and total person-years within each comparison in Supplementary (pp 49-50). The other severe COVID-19 outcomes of intensive care unit admission and death were rarer (Supplementary pp 46 and 51-53). NE denotes not estimated meaning that the risk estimates could not be generated for this specific country comparison.

### Heterologous- vs homologous booster schedules

The effectiveness of heterologous booster schedules against COVID-19 outcomes was largely comparable to that of the homologous booster schedules (Figure 1, Figure 4-5, and Supplementary pp 57-67). However, estimates, particularly for infection, varied across comparisons. The number of severe COVID-19 cases were generally low or zero for both the heterologous- and homologous booster schedules in all countries. For those comparisons where analyses were feasible, we observed comparable effectiveness of heterologous- and homologous booster schedules (Supplementary pp 63-67). Risk differences for hospitalisation for COVID-19 were lower than 1 per 1000 individuals in most comparisons; for AZD1BNT2BNT3 vs BNT1BNT2BNT3, CVE was 29.4% (95% CI -15.4%-74.2%) in Denmark, -16.7% (−130.9%-97.4%) in Finland, 6.8% (−42.9%-56.4%) in Norway, 11.7% (−41.8%-65.1%) in Sweden, and for BNT1BNT2MOD3 vs BNT1BNT2BNT3, CVE was -47.6% (−236.3%-100.0%) in Finland, 34.3% (13.5%-55.1%) in Norway, and 34.0% (18.1%-49.8%) in Sweden (not estimable in Denmark). Similarly, our comparisons of homologous BNT and MOD booster schedules showed largely similar effectiveness against severe COVID-19 outcomes (Supplementary pp 68-69).

**Figure 4.**
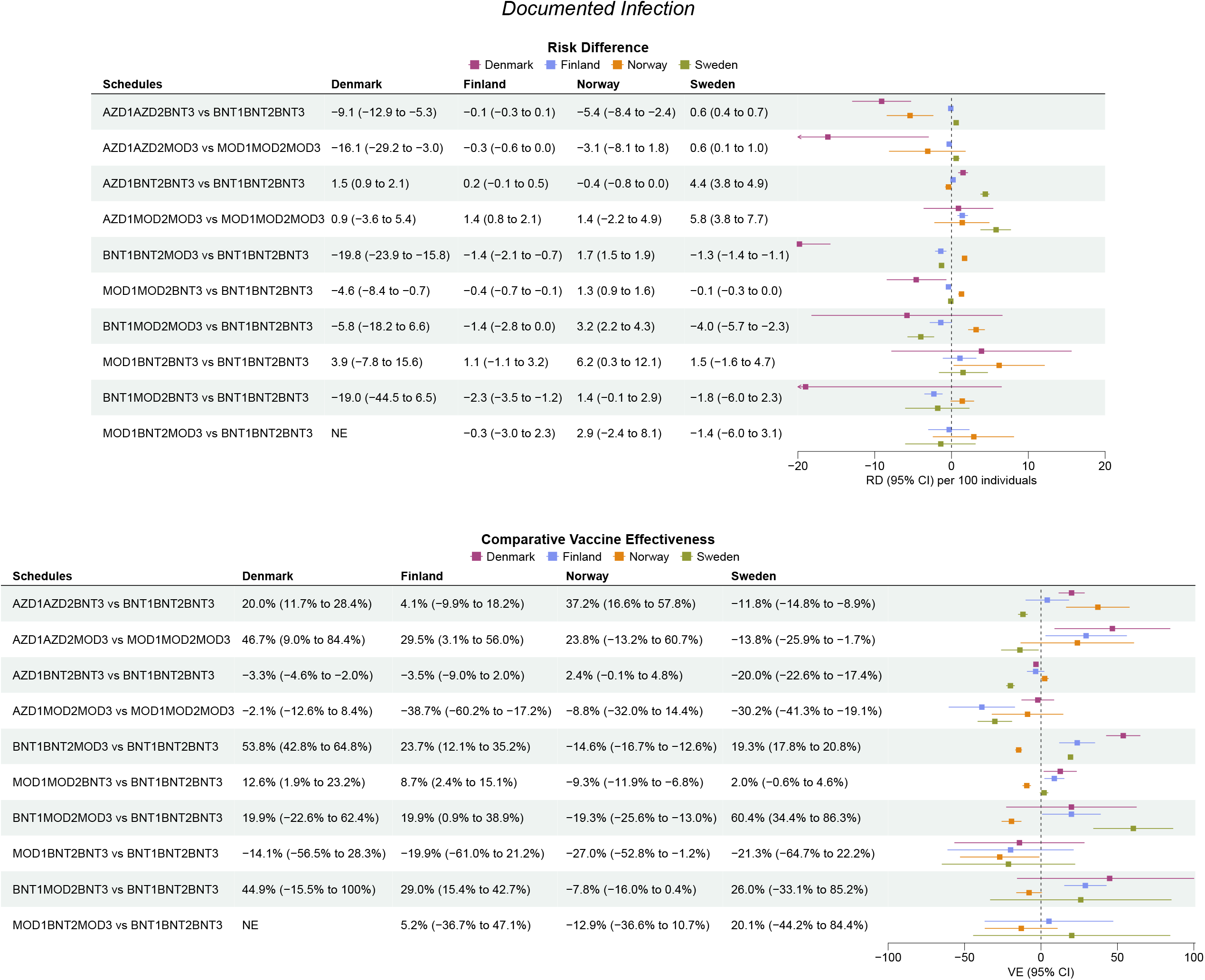
Risk of documented SARS-CoV-2 infection comparing heterologous-with homologous booster schedules in each country. Risk difference per 100 individuals and comparative vaccine effectiveness (1 – risk ratio) for documented SARS-CoV-2 infection at day 75 after start of follow-up were adjusted for calendar month of receiving the booster dose, year of birth (5-year bins), sex, region of residence, vaccination priority group, and selected comorbidities using inverse probability weights. Risk estimates are presented together with number of events and total person-years within each comparison in Supplementary (pp 61-62). NE denotes not estimated meaning that the risk estimates could not be generated for this specific country comparison.

**Figure 5.**
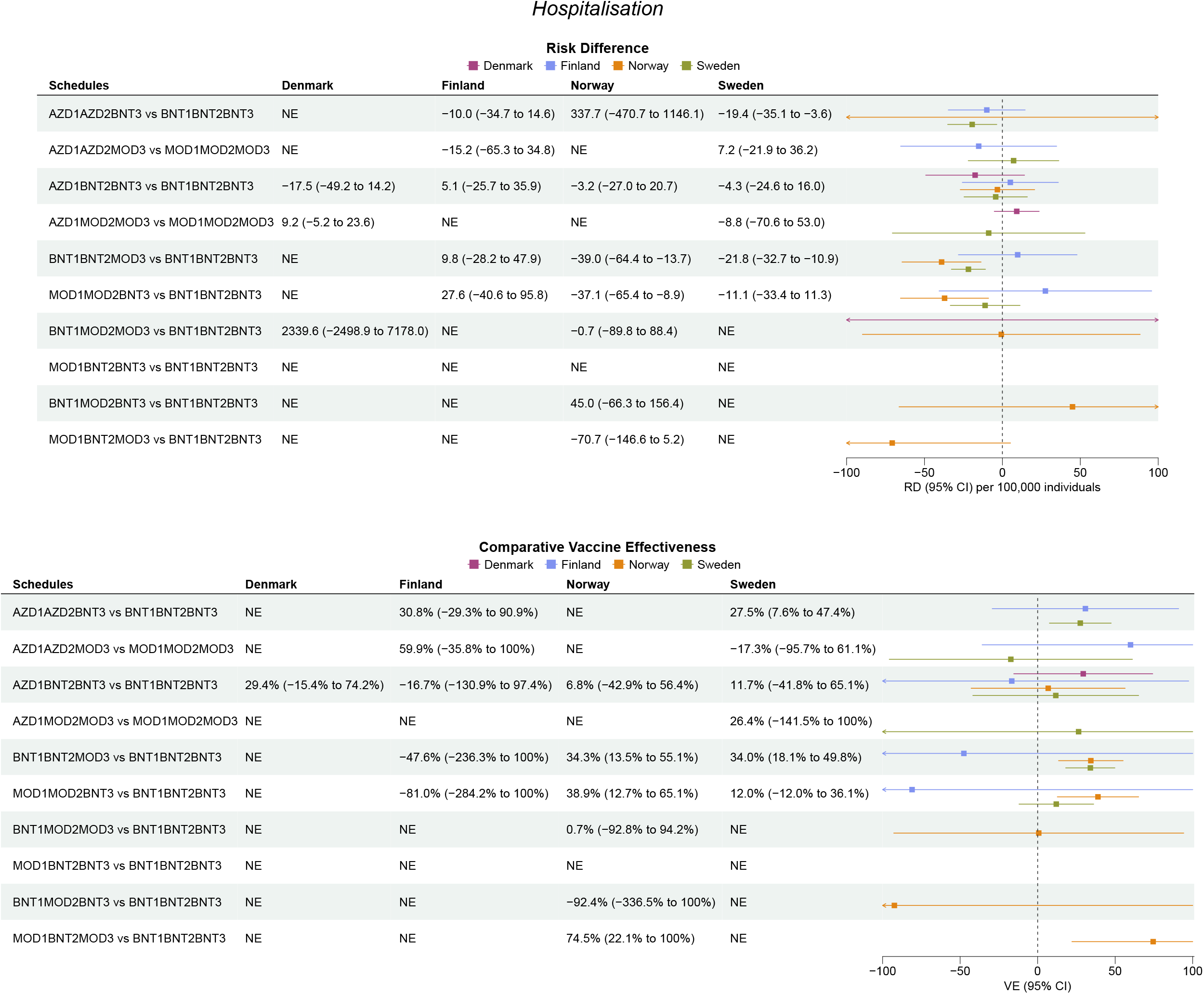
Risk of hospitalisation for COVID-19 comparing heterologous-with homologous booster schedules in each country. Risk difference per 100,000 individuals and comparative vaccine effectiveness (1 – risk ratio) for hospitalisation for COVID-19 at day 75 after start of follow-up were adjusted for calendar month of receiving the booster dose, year of birth (5-year bins), sex, region of residence, vaccination priority group, and selected comorbidities using inverse probability weights. Risk estimates are presented together with number of events and total person-years within each comparison in Supplementary (pp 63-64). The other severe COVID-19 outcomes of intensive care unit admission and death were rarer (Supplementary pp 65-67). NE denotes not estimated meaning that the risk estimates could not be generated for this specific country comparison.

### Additional analyses

Throughout all analyses, we observed no apparent differences in the effectiveness between those heterologous schedules that included AZD and those of mRNA vaccines only. In analyses which included previously SARS-CoV-2 infected individuals (Supplementary pp 70-80), we observed that both heterologous- and homologous booster schedules generally improved protection against documented infection compared with primary schedules, regardless of previous history of infection. Similarly, the comparable effectiveness found between heterologous- and homologous booster schedules, was not affected by previous infection. Moreover, among booster vaccinated individuals, those who had had a previous infection had lower incidences of documented infection compared with those who had not (Supplementary pp 70-80).

The results of our weighted analyses remained unchanged when adjusting for calendar week of receipt of the booster instead of calendar month (Supplementary pp 81-82). When examining the risk of documented infection by periods in Denmark with different infection rates, we observed a tendency towards a decrease in the CVE associated with later calendar periods where omicron infection rates were highest (Supplementary pp 83-84).

## DISCUSSION

We evaluated the effectiveness of heterologous booster schedules with the AZD, BNT, and MOD vaccines as compared with primary schedules and with homologous mRNA-vaccine booster schedules during a period of omicron variant predominance in large nationwide cohorts using register data from the Nordic countries. Overall, we found that the heterologous booster schedules improved protection against both SARS-CoV-2 infection and hospitalisation for COVID-19 when compared with primary schedules. Although risk estimates varied across the evaluated schedules and countries, particularly for infection, the effectiveness against severe COVID-19 outcomes was more consistent. We also found that the heterologous- and homologous booster schedules had comparable effectiveness; for most comparisons, risk differences of severe outcomes were lower than 1 per 1000 vaccinated individuals. Lastly, we found that the comparative effectiveness of heterologous boosters did not differ by previous infection, and hybrid immunity from a booster schedule and previous infection conferred the greatest protection.

Studies have shown higher effectiveness of boosters against infection during periods of delta variant predominance than for omicron periods where estimates are lower.[4,5,17,20,21] However, similar to our results, previous omicron estimates vary: compared with primary schedules, the booster effectiveness against symptomatic infection were 66% and 57% in test-negative studies from the US[20] and Scotland[22], respectively, and 51% against any infection in a cohort study from Spain[19] and 43% in a study from the US[23]. Using unvaccinated individuals as the reference group, a test-negative study from England reported effectiveness estimates of boosters against symptomatic infection at week 2 to 9 from vaccination between 47% and 74% across intervals and schedules.[17] The comparison of observed effectiveness between studies and countries, in particular for the outcome of SARS-CoV-2 infection, is inherently difficult. The most likely reason for the variations in our and other analyses are differences in captured infection rates which can be due to 1) the study period, which highly correlates to the omicron surge, 2) national COVID-19 policies including testing, restriction, and vaccination strategies, 3) individual-level testing and risk behaviour, and 4) population infection rates. These difficulties also highlight the need for both absolute and relative measures to properly interpret effectiveness estimates. Nonetheless and similar to previous data on homologous boosters,[5,21,24–27] the booster schedules we assessed generally provided increased protection against omicron-related COVID-19 outcomes including both infection and hospitalisation when compared with primary schedules.

Data to assess the effectiveness of heterologous booster schedules, however, are sparse and mainly limited to studies conducted during periods of delta variant predominance and/or as compared with unvaccinated.[10–15,17,18] To our knowledge, no study has reported the effectiveness of the individual heterologous booster schedules with the AZD, BNT, and MOD vaccines as compared with primary schedules during an omicron period. When compared with unvaccinated, heterologous and homologous schedules showed similar levels of improved protection against omicron infection[17] and severe COVID-19[18].

However, comparisons to unvaccinated hold concerns of healthy vaccinee bias and fundamental differences in testing and risk behaviour that are likely more extensive when aiming for assessing the effectiveness of boosters. Also, those who have chosen to remain unvaccinated in the current setting are unlikely to be a proper representative sample of the general population, and such comparisons provide little evidence to inform vaccination strategies. Studies during delta predominance periods overall found improved protection of heterologous boosters compared with primary schedules, a protection that was comparable to the homologous boosters when indirectly compared.[10–13] However, the protection against severe COVID-19 was typically not analysed in these studies or the number of cases was very low precluding statistical precision.

Previously, only one study has directly compared the risk of omicron infection between heterologous- and homologous mRNA-boosted individuals; severe COVID-19 was not investigated.[19] This Spanish study reported small risk differences (≤25 per 10,000 individuals), favouring MOD as the booster dose, regardless of the primary mRNA vaccine schedule received.[19] We observed a similar pattern in the BNT1BNT2MOD3 vs BNT1BNT2BNT3 and the MOD1MOD2MOD3 vs BNT1BNT2BNT3 comparisons in Denmark, Finland, and Sweden; however, in Norway, the associations were reversed or insignificant, respectively. Two studies that both combined delta and omicron periods were also suggestive of a greater protection against infection associated with receipt of a MOD booster dose.[28,29] However, a US study, although conducted during a period of delta predominance, found no significant differences in the risk of infection or severe COVID-19 when comparing matched heterologous- and homologous mRNA-boosted individuals;[16] however, sample sizes were likely too small for sufficient statistical power to detect differences with respect to severe outcomes. Our results also agree with those from studies that have reported comparable SARS-CoV-2-binding antibody responses after heterologous AZD, BNT, and/or MOD- and homologous mRNA vaccination.[6–9]

In line with recent work,[30] we found that those with a previous history of SARS-CoV-2 infection had better protection against infection compared to those without. This tendency was consistent regardless of receipt of a booster. Furthermore, hybrid immunity resulting from having been boosted and previously infected conferred the greatest protection. Importantly, the comparative effectiveness of heterologous- and homologous booster schedules did not differ by previous infection. These novel findings have likewise not been previously reported, but provides significant clinical insights to inform vaccination policies further.

Our study should be evaluated in light of potential limitations. First, although the results across the four participating countries are qualitatively similar, quantitative differences in our country-specific results made pooled meta-analyses unfeasible. Given that calendar time was strongly correlated with both the omicron surge and the individual vaccine schedules, our results from the individual comparisons depended on the shifting population infection rates at the time of follow-up. This is particularly relevant for the interpretation of our estimates for documented infection. Throughout most of the study period, the number of daily recorded infections in Denmark was twice as high as in Norway and Sweden and more than 4 times higher as in Finland. To mitigate biases from temporal differences in the COVID-19 endpoint ascertainment between the distinct in-country compared schedules, we used comparative designs and controlled for calendar time. While, our between-booster schedule comparisons were potentially vulnerable to such temporal bias in periods with rapid increases in the infection rates, results were unchanged in the analysis with more stringent calendar time control (week- vs month-adjustments).

Second, as noted, differences in the ascertainment of infection, due to differences in national COVID-19 policies and testing capacities and individual-level testing and risk behaviour as well as changes hereof over time, likely influence an estimation of effectiveness against infection. Compared to the other Nordic countries, Denmark had a proportionally larger testing capacity and registered use of PCR tests for SARS-CoV-2 during the study period (daily tests peaked at approximately 40.2, 8.6, 6.6, and 5.3 per 1000 individuals in Denmark, Finland, Norway, and Sweden, respectively) and PCR tests were freely available to anyone, regardless of symptoms or being in certain key/risk groups. Furthermore, we had no information on home-testing results nor on whether individuals had symptoms. Thus, captured infections likely represent both symptomatic and asymptomatic individuals. Our booster- vs primary schedule comparisons for risk of infection would be more prone to these potential ascertainment biases, but they most likely had no significant influence on our between-booster schedule comparisons. For example, if receipt of a booster dose was associated with greater testing or risk behaviour compared with not having received a booster dose, results could be biased toward the null; and, this bias would likely be amplified by more open public testing policies.

Third, a limitation to our outcome ascertainment of severe COVID-19 is that we may potentially have captured individuals with an outcome not directly related to COVID-19 but where COVID-19 was a contributing factor or co-occurred at the time of hospitalisation or death. Our COVID-19 hospitalisation definitions tried mitigating this by only including inpatient hospitalisation events considered having been due to COVID-19. Moreover, such potential misclassifications are most likely non-differential between vaccinated comparative groups.

Lastly, due to the inherent effectiveness of the booster doses against especially severe COVID-19 outcomes, we utilised a weighted design, in comparison to a matched design, for the between-booster comparisons to avoid a loss in statistical power due to missing matches. However, as we studied each heterologous booster schedule individually to help inform patients, clinicians, and regulatory authorities on the booster schedule-specific effectiveness observed in the Nordic countries and as the relative number of severe cases was low, the statistical precision was reduced for some comparisons. Likewise, analyses could not be performed when the number of vaccinated individuals was too small.

## CONCLUSION

In conclusion, we utilized the health care registers of the Nordic countries to study the vaccine effectiveness of heterologous booster schedules with the AZD, BNT, and MOD vaccines in four nationwide cohorts during a period of omicron predominance. We found that the heterologous booster schedules increased protection against both SARS-CoV-2 infection and hospitalisation for COVID-19 when compared with primary schedules. Additionally, the effectiveness of the heterologous booster schedules against COVID-19 was comparable to that of homologous mRNA-vaccine booster schedules. Hybrid immunity from previous infection and a booster provided the greatest protection.

## Supporting information

ACKNOWLEDGMENT SECTION

## Data Availability

No additional data available. Owing to data privacy regulations in each country, the raw data cannot be shared.

## Contributors

NA, ET, and AH conceptualized the study; NA drafted the manuscript; ET, UB, JS, and NP carried out the statistical analyses. All authors interpreted the results and critically reviewed the manuscript. AH supervised the study. NA and AH are the guarantors. The corresponding author attests that all listed authors meet authorship criteria and that no others meeting the criteria have been omitted.

## Transparency

The lead author (the manuscript’s guarantor) affirms that this manuscript is an honest, accurate, and transparent account of the study being reported; that no important aspects of the study have been omitted; and that any discrepancies from the study as planned (and, if relevant, registered) have been explained.

## Ethics

The Danish study was performed as a surveillance study as part of the governmental institution Statens Serum Institut’s (SSI) advisory tasks for the Danish Ministry of Health. SSI’s purpose is to monitor and fight the spread of disease in accordance with section 222 of the Danish Health Act. According to Danish law, national surveillance activities conducted by SSI do not require approval from an ethics committee. It was approved by the Danish Governmental law firm and SSI’s compliance department that the study is fully compliant with all legal, ethical and IT-security requirements and there are no further approval procedures regarding such studies. For the Finnish study, by Finnish law, the Finnish Institute for Health and Welfare (THL) is the national expert institution to carry out surveillance on the impact of vaccinations in Finland (Communicable Diseases Act, https://www.finlex.fi/en/laki/kaannokset/2016/en20161227.pdf). Neither specific ethical approval (a waiver of ethical approval was received from Professor Mika Salminen, Director of the Department for Health Security Finnish Institute for Health and Welfare) of this study nor informed consent from the participants was needed. The Norwegian study was approved by the Norwegian Regional Committee for Health Research Ethics South East (REK Sør-Øst A, ref 122745), and has conformed to the principles embodied in the Declaration of Helsinki. The emergency preparedness register was established according to the Health Preparedness Act §2-4. Consent to participate was not applicable as this is a register-based study. The Swedish study is approved by the Swedish Ethical Review Authority (2020-06859, 2021-02186) and has conformed to the principles embodied in the Declaration of Helsinki. Consent to participate is not applicable as this is a register-based study. Due to the nature of this research, there was no involvement of patients or members of the public in the design or reporting of this study.

## Disclaimer

This document expresses the opinion of the authors of the paper, and may not be understood or quoted as being made on behalf of, or reflecting the position of the European Medicines Agency or one of its Committees or Working Parties.

## Dissemination to participants and related patient and public communities

Studied participants were anonymised in the utilised data sources; and therefore, direct dissemination to study participants is not possible. The study results will be disseminated to the public and health professionals by a press release written using layman’s terms.

## REFERENCES

1 Chemaitelly H, Tang P, Hasan MR, et al. Waning of BNT162b2 Vaccine Protection against SARS-CoV-2 Infection in Qatar. New England Journal of Medicine 2021;385:e83. doi:10.1056/NEJMoa2114114

2 Tregoning JS, Flight KE, Higham SL, et al. Progress of the COVID-19 vaccine effort: viruses, vaccines and variants versus efficacy, effectiveness and escape. Nat Rev Immunol 2021;21:626–36. doi:10.1038/s41577-021-00592-1

3 Bar-On YM, Goldberg Y, Mandel M, et al. Protection of BNT162b2 Vaccine Booster against Covid-19 in Israel. New England Journal of Medicine. 2021;385:1393–400.

4 Barda N, Dagan N, Cohen C, et al. Effectiveness of a third dose of the BNT162b2 mRNA COVID-19 vaccine for preventing severe outcomes in Israel: an observational study. The Lancet 2021;398:2093– 100. doi:10.1016/S0140-6736(21)02249-2

5 Abu-Raddad LJ, Chemaitelly H, Ayoub HH, et al. Effect of mRNA Vaccine Boosters against SARS-CoV-2 Omicron Infection in Qatar. New England Journal of Medicine 2022;386:1804–16. doi:10.1056/NEJMoa2200797

6 Munro APS, Janani L, Cornelius V, et al. Safety and immunogenicity of seven COVID-19 vaccines as a third dose (booster) following two doses of ChAdOx1 nCov-19 or BNT162b2 in the UK (COV-BOOST): a blinded, multicentre, randomised, controlled, phase 2 trial. The Lancet 2021;398:2258–76. doi:10.1016/S0140-6736(21)02717-3

7 Stuart ASV, Shaw RH, Liu X, et al. Immunogenicity, safety, and reactogenicity of heterologous COVID-19 primary vaccination incorporating mRNA, viral-vector, and protein-adjuvant vaccines in the UK (Com-COV2): a single-blind, randomised, phase 2, non-inferiority trial. Lancet 2022;399:36–49. doi:10.1016/S0140-6736(21)02718-5

8 Borobia AM, Carcas AJ, Pérez-Olmeda M, et al. Immunogenicity and reactogenicity of BNT162b2 booster in ChAdOx1-S-primed participants (CombiVacS): a multicentre, open-label, randomised, controlled, phase 2 trial. Lancet 2021;398:121–30. doi:10.1016/S0140-6736(21)01420-3

9 Parker EPK, Desai S, Marti M, et al. Emerging evidence on heterologous COVID-19 vaccine schedules— To mix or not to mix? The Lancet Infectious Diseases 2022;22:438–40. doi:10.1016/S1473-3099(22)00178-5

10 Andrews N, Stowe J, Kirsebom F, et al. Effectiveness of COVID-19 booster vaccines against COVID-19-related symptoms, hospitalization and death in England. Nat Med 2022;28:831–7. doi:10.1038/s41591-022-01699-1

11 Hulme WJ, Williamson EJ, Horne E, et al. Effectiveness of BNT162b2 booster doses in England: an observational study in OpenSAFELY-TPP. medRxiv. 2022;:2022.06.06.22276026.

12 Tan SHX, Pung R, Wang L-F, et al. Association of Homologous and Heterologous Vaccine Boosters With COVID-19 Incidence and Severity in Singapore. JAMA 2022;327:1181–2. doi:10.1001/jama.2022.1922

13 Marra AR, Miraglia JL, Malheiros DT, et al. Effectiveness of Heterologous Coronavirus Disease 2019 (COVID-19) Vaccine Booster Dosing in Brazilian Healthcare Workers, 2021. Clinical Infectious Diseases 2022;:ciac430. doi:10.1093/cid/ciac430

14 Menni C, May A, Polidori L, et al. COVID-19 vaccine waning and effectiveness and side-effects of boosters: a prospective community study from the ZOE COVID Study. Lancet Infect Dis 2022;22:1002– 10. doi:10.1016/S1473-3099(22)00146-3

15 Starrfelt J, Danielsen AS, Buanes EA, et al. Age and product dependent vaccine effectiveness against SARS-CoV-2 infection and hospitalisation among adults in Norway: a national cohort study, July– November 2021. BMC Medicine 2022;20:278. doi:10.1186/s12916-022-02480-4

16 Mayr FB, Talisa VB, Shaikh O, et al. Effectiveness of Homologous or Heterologous Covid-19 Boosters in Veterans. New England Journal of Medicine 2022;386:1375–7. doi:10.1056/NEJMc2200415

17 Andrews N, Stowe J, Kirsebom F, et al. Covid-19 Vaccine Effectiveness against the Omicron (B.1.1.529) Variant. New England Journal of Medicine 2022;386:1532–46. doi:10.1056/NEJMoa2119451

18 Baum U, Poukka E, Leino T, et al. High vaccine effectiveness against severe Covid-19 in the elderly in Finland before and after the emergence of Omicron. medRxiv. 2022;:2022.03.11.22272140.

19 Monge S, Rojas-Benedicto A, Olmedo C, et al. Effectiveness of mRNA vaccine boosters against infection with the SARS-CoV-2 omicron (B.1.1.529) variant in Spain: a nationwide cohort study. The Lancet Infectious Diseases 2022;22:1313–20. doi:10.1016/S1473-3099(22)00292-4

20 Accorsi EK, Britton A, Fleming-Dutra KE, et al. Association Between 3 Doses of mRNA COVID-19 Vaccine and Symptomatic Infection Caused by the SARS-CoV-2 Omicron and Delta Variants. JAMA 2022;327:639–51. doi:10.1001/jama.2022.0470

21 Tseng HF, Ackerson BK, Luo Y, et al. Effectiveness of mRNA-1273 against SARS-CoV-2 Omicron and Delta variants. Nat Med 2022;28:1063–71. doi:10.1038/s41591-022-01753-y

22 Sheikh A, Kerr S, Woolhouse M, et al. Severity of omicron variant of concern and effectiveness of vaccine boosters against symptomatic disease in Scotland (EAVE II): a national cohort study with nested test-negative design. The Lancet Infectious Diseases 2022;22:959–66. doi:10.1016/S1473-3099(22)00141-4

23 Ioannou GN, Bohnert ASB, O’Hare AM, et al. Effectiveness of mRNA COVID-19 Vaccine Boosters Against Infection, Hospitalization, and Death: A Target Trial Emulation in the Omicron (B.1.1.529) Variant Era. Ann Intern Med Published Online First: 11 October 2022. doi:10.7326/M22-1856

24 Buchan SA, Chung H, Brown KA, et al. Estimated Effectiveness of COVID-19 Vaccines Against Omicron or Delta Symptomatic Infection and Severe Outcomes. JAMA Network Open 2022;5:e2232760. doi:10.1001/jamanetworkopen.2022.32760

25 Stowe J, Andrews N, Kirsebom F, et al. Effectiveness of COVID-19 vaccines against Omicron and Delta hospitalisation, a test negative case-control study. Nat Commun 2022;13:5736. doi:10.1038/s41467-022-33378-7

26 Thompson MG. Effectiveness of a Third Dose of mRNA Vaccines Against COVID-19–Associated Emergency Department and Urgent Care Encounters and Hospitalizations Among Adults During Periods of Delta and Omicron Variant Predominance — VISION Network, 10 States, August 2021–January 2022. MMWR Morb Mortal Wkly Rep 2022;71. doi:10.15585/mmwr.mm7104e3

27 Lauring AS, Tenforde MW, Chappell JD, et al. Clinical severity of, and effectiveness of mRNA vaccines against, covid-19 from omicron, delta, and alpha SARS-CoV-2 variants in the United States: prospective observational study. BMJ 2022;376:e069761. doi:10.1136/bmj-2021-069761

28 Hulme WJ, Horne EM, Parker EP, et al. Comparative effectiveness of BNT162b2 versus mRNA-1273 boosting in England: a cohort study in OpenSAFELY-TPP. medRxiv. 2022;:2022.07.29.22278186.

29 Ono S, Michihata N, Yamana H, et al. Comparative effectiveness of BNT162b2 and mRNA-1273 booster dose after BNT162b2 primary vaccination against the Omicron variants: A retrospective cohort study using large-scale population-based registries in Japan. Clin Infect Dis 2022;:ciac763. doi:10.1093/cid/ciac763

30 Altarawneh HN, Chemaitelly H, Ayoub HH, et al. Effects of Previous Infection and Vaccination on Symptomatic Omicron Infections. New England Journal of Medicine 2022;387:21–34. doi:10.1056/NEJMoa2203965

