## Supplementary material for "Comparative effectiveness of heterologous booster schedules with AZD1222, BNT162b2, or mRNA-1273 vaccines against COVID-19 during omicron predominance in the Nordic countries": ACKNOWLEDGMENT SECTION

### Supplementary Table 1. Description of utilized registers within each country.

| Data source (country) | Details of the individual-level data sources |
|---|---|
| **General points for the data sources in all countries:** The unique nationwide register data available to us allowed for the construction of country-specific cohorts with individual-level information on dates of vaccination and dates of endpoints together with relevant covariate information. All Nordic residents are assigned a unique personal identifier at birth or immigration, enabling unambiguous linkage between registers. Thus, the data from all the Nordic countries were based on individual-level information and we had full data availability during the study period. All countries have universal and tax-financed healthcare systems and reporting to national registers is mandatory, providing near-complete follow-up of all residents over time. | |
| **DENMARK** | |
| The Civil Registration System [1] | The register provides the mandatory unique personal identifier for all permanent residents of Denmark, which allows linkage between all Danish health care services and civil registrations systems. The register has existed since Apr 2, 1968. In addition, it holds general demographic information such as birthdate and sex as well as continuously updated information and dates on historical addresses, immigration and emigration status, and death. |
| The Danish Vaccination Register [2] | The register holds information on all vaccinations given in Denmark including information on vaccination date, type, dose, and product batch number ever since Nov 15, 2015 (where reporting to the register became mandatory). Specifically related to this study, the Danish Health Agency have provided the governmentally assigned COVID-19 vaccine priority groups that were prioritized groups according to the risk of severe infection as well as whether being health and social care workers. |
| The Danish Microbiology Database [3] | Information on positive PCR tests for SARS-CoV-2 were drawn from The Danish Microbiology Database (MiBa) that holds information on all microbiology samples analysed at Danish departments of microbiology, including information on SARS-CoV-2 PCR test results, date of sampling, date of analysis, type of test, and interpretation of test. The SARS-CoV-2 PCR tests have been freely available to all individuals in Denmark regardless of symptoms status throughout the COVID-19 pandemic. |
| The National Patient Register [4] | The register covers all hospital contacts in Denmark with information on the duration of the contact, department of admission and other hospital characteristics. Treating physician-assigned diagnoses have been registered according to ICD-10 codes since 1994. |
| **FINLAND** | |
| Finnish Population Information System [5] | The register is an electronic register including personal data of all permanent residents in Finland. It contains demographic information such as the unique personal identifier in Finland, date of birth, place of residence, date of death, and date of immigration and emigration. The register is held by the Digital and Population Data Services Agency. |
| Register of Social Assistance [6] | The register holds information on individuals in long-term care and/or with need for social assistance including social rehabilitation. This assistance may be given in nursing homes, people’s own homes or other institutions. The register is held by the Finnish Institute for Health and Welfare. |
| Social and Healthcare Professionals Register [7] | The register contains person-level data on rights to act as health care personnel. |
| National Vaccination Register [8] | The register, which is based on the Register of Primary Health Care Visits, holds information on all COVID-19 vaccinations administered in Finland. Data include the date of vaccination, vaccine batch number and trade name. |
| National Infectious Diseases Register [9] | The register contains information on notifiable diseases which must be reported by the laboratories and the physician treating the patient, or performing an autopsy, in accordance with the Finnish Communicable Diseases Act. Thus, the sample dates of all SARS-CoV-2 infections that were laboratory-confirmed in Finland are recorded in this register. The register is held by the Finnish Institute for Health and Welfare. |
| National Care Register for Health Care [10] | The register comprises information on all in-hospital care (since 1969) and outpatient specialist care (since 1998) in Finland, including admission and discharge dates, whether hospitalisation was planned or acute, codes for discharge diagnoses (according to ICD-10) and surgical procedures, whether discharged as deceased, to own private residence or other health care facilities, type of department and hospital. The register is held by Finnish Institute for Health and Welfare. |
| Finnish Intensive Care Consortium's Quality Register for Intensive Care | The register records data on all patients treated in an intensive care unit in Finland. |
| Special Reimbursement Register and Prescription Centre database | These data collections are maintained by the Finnish Social Insurance Institution. The Special Reimbursement Register allows the identification of individuals entitled to special reimbursement for medical expenses. The Prescription Centre database allows the identification of individuals using selected medications of interest. |

| Data source (country) | Details of the individual-level data sources |
|---|---|
| Register of Primary Health Care Visits [11] | The register covers all outpatient primary health care services delivered in Finland. The register is held by Finnish Institute for Health and Welfare. |
| **NORWAY** | |
| The Emergency Preparedness Register for COVID-19 [12] (consisting of the data sources below) | Data for this study were obtained through the Emergency preparedness register for COVID-19 (“Beredt C19”), which is administered by the Norwegian Institute of Public Health, according to the Norwegian Health Preparedness Act §2-4. The register was established in 2020 to provide authorities with up-to-date information on prevalence, causal relationships, and consequences of the COVID-19 epidemic in Norway and includes the total population in Norway. The register includes information already collected in the healthcare system and the national health registries (see the following data sources). |
| Norwegian Population Register | The register holds information on birthdate, immigration and emigration status as well as and death for all residents of Norway. |
| State register of employers and employees (NAV AA register) [13] | The register holds lists of all employment relationships in Norway, and employers and contractors are obliged to report their employees and freelancers to the register. Employees are classified according to the Norwegian Standard Classification of Occupations) and can thus be used to obtain data on health care personnel status. |
| The Norwegian Information System for the Nursing and Care Sector (IPLOS) [14] | The register holds information on the health care services that are provided by municipalities in Norway. Report of applicants and recipients of such services to the register is mandatory for all municipalities. The register includes information on home care service and out-of-hospital institutional care, including short- and long-term nursing home stay. |
| The Norwegian Immunisation Register (SYSVAK) [15] | The register holds information of administered vaccines in Norwegian vaccination programs, including the date of administration and type of vaccine. For the COVID-19 vaccines, reporting to the register have been mandatory. |
| Norwegian Surveillance System for Communicable Diseases (MSIS) | The register holds information on selected infectious diseases for which reporting to the register is mandatory. This includes all COVID-19 tests and the date of testing and test results. |
| The Norwegian Patient Registry (NPR) [16] | The register holds information on all contacts with specialist health-care services in Norway, including admission and discharge dates as well as diagnoses (recorded according to ICD-10) during hospitalisation or outpatient contact. |
| The Norwegian Intensive Care and Pandemic Registry (NIPaR) [17] | The Norwegian Intensive Care and Pandemic Registry (NIPaR) is a national clinical registry that was expanded to include COVID-19 patients in conjunction with the COVID-19 pandemic. In the registry, all patients who have tested positive for SARS-CoV-2 and are admitted to hospital are registered. All Norwegian hospitals report to NIPaR, and reporting is mandatory. NIPaR also includes data on patients who have tested positive for SARS-CoV-2 and are admitted to an intensive care unit (ICU). |
| **SWEDEN** | |
| The Total Population Register [18] | The register contains information on the unique personal identifier for all individuals in Sweden as well as general demographic information such as date of birth, sex, country of birth, place of residence, and date of immigration and emigration. The register is held by Statistics Sweden. |
| The Cause of Death Register [19] | The register contains information place of residence at time of death, date and underlying cause of death and contributing causes of death. |
| The Longitudinal Integrated Database For Health Insurance And Labour Market Studies (LISA) [20] | The database contains a wide range of socioeconomic information including occupation (such as healthcare worker). The register is held by Statistics Sweden. |
| Register On Persons In Nursing Homes [21] | The register holds information on nursing care given to elderly and/or persons with physical, psychiatric or intellectual disabilities at either nursing homes, own homes or other institutions. The register is held by the National Board of Health and Welfare. |
| The National Vaccination Register [22] | The register contains information on administered COVID-19 vaccines including data on date of administration, the specific vaccine products, substance, formulation, batch number and dose number (for repeated doses) since Jan 1, 2021. The register is held by the Public Health Agency of Sweden. |
| Register On Surveillance Of Notifiable Communicable Diseases (Sminet) [23] | The register contains information on notifiable diseases (for which reporting is mandatory) reported by either the analysis performing laboratories, the treating physician or autopsy performing physician, in accordance with the Swedish Communicable Diseases Act. Data include date of disease occurrence, date of testing, date of positive test and diagnoses. The register is held by the Public Health Agency of Sweden. |

| Data source (country) | Details of the individual-level data sources |
|---|---|
| The Swedish Patient Register [24,25] | The register comprises information on all in-hospital (since 1987) and out-patient (since 2001) specialist care in Sweden including data on admission and discharge dates, whether hospitalisation was planned or acute, codes for discharge diagnoses and surgical procedures, whether discharged as deceased, to own private residence or other health care facilities, type of department, and hospital. For the current study period discharge diagnoses were recorded according to the Swedish clinical modification of the ICD-10 (ie, ICD-10-SE). The register is held by the National Board of Health and Welfare. |

**Supplementary Table 2. Detailed description of included variables and data sources in each country.**

| VARIABLE | COUNTRY | DATA SOURCE AND DETAILS | VALUES/CODES |
|---|---|---|---|
| **BASELINE VARIABLES** | | | |
| Age | Denmark | *The Civil Registration System.*<br>Defined as age at first COVID-19 vaccination. | Categorical: 5-year bins |
| | Finland | *The Finnish Population Information System.*<br>Defined as age at first COVID-19 vaccination. | |
| | Norway | *Norwegian Population Register.*<br>Defined as age at first COVID-19 vaccination. | |
| | Sweden | *The Total Population Register.*<br>Defined as age at first COVID-19 vaccination. | |
| Sex | Denmark | *The Civil Registration System.*<br>Defined as biological sex. | Binary: male, female |
| | Finland | *The Finnish Population Information System.*<br>Defined as biological sex. | |
| | Norway | *Norwegian Population Register.*<br>Defined as biological sex. | |
| | Sweden | *The Total Population Register.*<br>Defined as biological sex. | |
| Residency (citizenship) | Denmark | *The Civil Registration System.*<br>Defined as known national resident. | Binary: yes/no |
| | Finland | Not available. | |
| | Norway | *Norwegian Population Register.*<br>Defined as known national resident. | |
| | Sweden | *The Total Population Register.*<br>Defined as known national resident. | |
| Calendar month | Denmark | *The Danish Vaccination Register.*<br>Defined by the date where the respective vaccine dose examined was administered (i.e. second or third dose) and grouped into monthly intervals according to months since start of study period. | Categorical (14 levels): calendar month 1 (Dec 27, 2020 to Jan 31, 2021) to month 14 (February 2022) |
| | Finland | *The National Vaccination Register.*<br>Defined by the date where the respective vaccine dose examined was administered (i.e. second or third dose) and grouped into monthly intervals according to months since start of study period. | |
| | Norway | *The Norwegian Immunisation Register (SYSVAK).*<br>Defined by the date where the respective vaccine dose examined was administered (i.e. second or third dose) and grouped into monthly intervals according to months since start of study period. | |
| | Sweden | *The National Vaccination Register.*<br>Defined by the date where the respective vaccine dose examined was administered (i.e. second or third dose) and grouped into monthly intervals according to months since start of study period. | |
| Region of residency | Denmark | *The Civil Registration System.*<br>Defined by last known address at first vaccination. | Categorical: DK, 5 levels; FI, 5 levels; NO, 5 levels; SE, 9 levels |
| | Finland | *The Finnish Population Information System.*<br>Defined by last known address. | |

| VARIABLE | COUNTRY | DATA SOURCE AND DETAILS | VALUES/CODES |
|---|---|---|---|
| | Norway | *Norwegian Population Register.*<br>Defined by last known address. | |
| | Sweden | *The Total Population Register.*<br>Defined by last known address at first vaccination. | |
| COVID-19 vaccine priority groups[a] | Denmark | *The Danish Vaccination Register.*<br>Defined as governmentally assigned COVID-19 vaccine priority groups, prioritized according to the risk of severe infection as well as whether being health and social care workers (assigned before first COVID-19 vaccination). | Categorical (4 levels): Target risk groups, healthcare personnel, selected relatives of people at high risk, others |
| | Finland | *Register of Social Assistance.*<br>Vulnerable individuals defined as individuals in 24-hours care (binary status per Dec 27, 2020).<br><br>*Social and Healthcare Professionals Register.*<br>Healthcare personnel defined as individuals with the right to act as health care personnel as of Dec 27, 2020. | Categorical (3 levels): Vulnerable individuals, healthcare personnel, others |
| | Norway | *The Norwegian Information System for the Nursing and Care Sector.*<br>Vulnerable individuals defined as nursing home resident (binary status per Dec 27, 2020).<br><br>*State register of employers and employees.*<br>Healthcare personnel defined as binary status per Dec 27, 2020. | Categorical (3 levels): Vulnerable individuals, healthcare personnel, others |
| | Sweden | *Register on persons in nursing homes.*<br>Vulnerable individuals defined as nursing home resident (binary status as of December 2020)<br><br>*The Longitudinal integrated database for health insurance and labour market studies.*<br>Healthcare personnel defined as healthcare worker occupation status as of October 2018 (binary). | Categorical (3 levels): Vulnerable individuals, healthcare personnel, others |
| Comorbidity 1: Chronic pulmonary disease (CPD) | Denmark | *The National Patient Register.*<br>Defined as primary diagnoses regardless of type of hospital contact registered before first COVID-19 vaccination (look-back 3 years). | Binary: yes/no<br><br>ICD-10 codes: J40-J47, J60–J67, J684, J701, J703, J841, J920, J961, J982, J983 |
| Comorbidity 1: CPD | Finland | *Care register for Health Care.*<br>Defined as primary or secondary diagnoses before Dec 27, 2020 (look-back 6 years). | Binary: yes/no<br><br>ICD-10 codes: J41-J44, J47 |
| Comorbidity 1: CPD | Norway | *Norwegian Patient Register.*<br>Defined as any recorded ICD-10 diagnosis during inpatient or outpatient contact in hospital or from private-practicing specialists and before first COVID-19 vaccination (look-back 3 years). | Binary: yes/no<br><br>ICD-10 codes: E84, J41-J47, J701, J703, J84, J98 |
| Comorbidity 1: CPD | Sweden | *National Patient Register.*<br>Defined as any recorded ICD-10 diagnosis during inpatient or outpatient contact and before first COVID-19 vaccination (look-back 3 years). | Binary: yes/no<br><br>ICD-10 codes: E84, J41-J47 J84, J98 |
| Comorbidity 2: Cardiovascular conditions and diabetes (CVD/DM) | Denmark | *The National Patient Register.*<br>Defined as primary diagnoses regardless of type of hospital contact registered before first COVID-19 vaccination (look-back 3 years). | Binary: yes/no<br><br>ICD-10 codes: E10-E11, I110, I130, I132, I20-I23, I420, I426-I429, I48, I500-I503, I508, I509 |

| VARIABLE | COUNTRY | DATA SOURCE AND DETAILS | VALUES/CODES |
|---|---|---|---|
| Comorbidity 2: CVD/DM | Finland | *Care register for Health Care, Register of Primary Health Care Visits, Special Reimbursement Register and Prescription Centre database.*<br>Defined as primary or secondary diagnoses (look-back 6 years) or drug prescriptions (look-back 3 years) before Dec 27, 2020. | Binary: yes/no<br><br>ICD-10 codes: E10, E11, E13, E14, I11–I13, I15, I20–I25<br>ICPC-2 codes: T89, T90<br>ATC codes: A10A, A10B |
| Comorbidity 2: CVD/DM | Norway | *Norwegian Patient Register.*<br>Defined as any recorded ICD-10 diagnosis during inpatient or outpatient contact in hospital or from private-practicing specialists and before first COVID-19 vaccination (look-back 3 years). | Binary: yes/no<br><br>ICD-10 codes: E10-E14<br>I05-I09, I110, I130, I132, I1420, I20-I23, I25-I28, I33-I39, I426-I429, I48, I50 |
| Comorbidity 2: CVD/DM | Sweden | *National Patient Register.*<br>Defined as any recorded ICD-10 diagnosis during inpatient or outpatient contact and before first COVID-19 vaccination (look-back 3 years).<br><br>*Swedish Prescribed Drug Register.*<br>Antidiabetic drugs use defined as ≥2 filled prescriptions during 2020. | Binary: yes/no<br><br>ICD-10 codes: E10-E14, I05-I09, I110, I20-I28, I34-I37, I39, I42, I43, I46, I48-I50<br>ATC code: A10 |
| Comorbidity 3: Autoimmunity related conditions (AIC)[b] | Denmark | *The National Patient Register.*<br>Defined as primary diagnoses regardless of type of hospital contact registered before first COVID-19 vaccination (look-back 3 years). | Binary: yes/no<br><br>ICD-10 codes: D510, D590, D591,<br>D690, D693, D86, E050, E063, E271, E272, G122G, G35, G610, G700, I00, I01, K50, K51, K743, K900, L12, L40, L52, L80, L93, M05, M06, M08, M300, M313, M315, M316, M32, M33, M34, M35, M45 |
| Comorbidity 3: AIC[b] | Finland | *Care register for Health Care, Special Reimbursement Register and Prescription Centre database.*<br>Defined as primary or secondary diagnoses (look-back 6 years) or drug prescriptions (look-back 3 years) before Dec 27, 2020.<br><br>**Only if patient also used one of the listed drugs (marked with **)*<br>***Only if patient also had one of the diagnoses marked with ** | Binary: yes/no<br><br>ICD-10 codes: D7081, D7089, D80–D84, E250, E271, E272, E274, E310, E896, D86*, K50*, K51*, L40*, M02*, M05–M07*, M139*, M45*, M460*, M461*, M469*, M941*<br><br>ATC-codes**: H02AB02, H02AB04, H02AB06, H02AB07, L01BA01, L01XC02, L04AA06, L04AA10, L04AA13, L04AA18, L04AA24, L04AA26, L04AA29, L04AA33, L04AA37, L04AB, L04AC, L04AD01, L04AD02, L04AX01, L04AX03 |
| Comorbidity 3: AIC[b] | Norway | *Norwegian Patient Register.*<br>Defined as any recorded ICD-10 diagnosis during inpatient or outpatient contact in hospital or from private-practicing specialists and before first COVID-19 vaccination (look-back 3 years). | Binary: yes/no<br><br>ICD-10 codes: G35, K50-K51, M05-M09, M13-M14 |
| Comorbidity 3: AIC[b] | Sweden | *National Patient Register.*<br>Defined as any recorded ICD-10 diagnosis during inpatient or outpatient contact and before first COVID-19 vaccination (look-back 3 years). | Binary: yes/no<br><br>ICD-10 codes: D86, G35, K50, K51, L40, M05-M09, M13, M14, M45 |
| Comorbidity 4: Cancer | Denmark | *The National Patient Register.* | Binary: yes/no |

| VARIABLE | COUNTRY | DATA SOURCE AND DETAILS | VALUES/CODES |
|---|---|---|---|
| | | Defined as primary diagnoses regardless of type of hospital contact registered before first COVID-19 vaccination (look-back 3 years). | ICD-10 codes: C00–C85 (without C44), C88, C90-C96 |
| Comorbidity 4: Cancer | Finland | *Care register for Health Care and Special Reimbursement Register.* Defined as primary or secondary diagnoses before Dec 27, 2020 (look-back 6 years). | Binary: yes/no<br><br>ICD-10 codes: C00–C97 (without C44), D051, D39 |
| Comorbidity 4: Cancer | Norway | *Norwegian Patient Register.* Defined as any recorded ICD-10 diagnosis during inpatient or outpatient contact in hospital or from private-practicing specialists and before first COVID-19 vaccination (look-back 3 years). | Binary: yes/no<br><br>ICD-10 codes: C00-C96 (without C44) |
| Comorbidity 4: Cancer | Sweden | *National Patient Register.* Defined as any recorded ICD-10 diagnosis during inpatient or outpatient contact and before first COVID-19 vaccination (look-back 3 years). | Binary: yes/no<br><br>ICD-10 codes: C00-C96 (without C44), D45-D47 |
| Comorbidity 5: Moderate to severe renal disease (CKD) | Denmark | *The National Patient Register.* Defined as primary diagnoses regardless of type of hospital contact registered before first COVID-19 vaccination (look-back 3 years). | Binary: yes/no<br><br>ICD-10 codes: I12, I13, N00–N05, N07, N11, N14, N17–N19, Q61 |
| Comorbidity 5: CKD | Finland | *Care register for Health Care.* Defined as primary or secondary diagnoses before Dec 27, 2020 (look-back 6 years). | Binary: yes/no<br><br>ICD-10 codes: I12, I13, N00–N05, N07, N08, N11, N14, N18, N19, E102, E112, E142 |
| Comorbidity 5: CKD | Norway | *Norwegian Patient Register.* Defined as any recorded ICD-10 diagnosis during inpatient or outpatient contact in hospital or from private-practicing specialists and before first COVID-19 vaccination (look-back 3 years). | Binary: yes/no<br><br>ICD-10 codes: I12-I13, N00-N05, N07, N11, N14, N17-N19, Q61 |
| Comorbidity 5: CKD | Sweden | *National Patient Register.* Defined as any recorded ICD-10 diagnosis during inpatient or outpatient contact and before first COVID-19 vaccination (look-back 3 years). | Binary: yes/no<br><br>ICD-10 codes: I12, I13, N00-N05, N07, N11, N14, N17-N19, Q61 |
| **EXPOSURE AND OUTCOME VARIABLES** | | | |
| Vaccination status[c] | Denmark | *The Danish Vaccination Register.* Defined according to the specific administered COVID-19 vaccines and date of vaccinations. | Categorical (multiple levels): AZD1AZD2BNT3, AZD1BNT2BNT3, AZD1MOD2MOD3, BNT1BNT2BNT3, BNT1BNT2MOD3 etc. |
| | Finland | *The National Vaccination Register.* Defined according to the specific administered COVID-19 vaccines and date of vaccinations. | |
| | Norway | *The Norwegian Immunisation Register (SYSVAK).* Defined according to the specific administered COVID-19 vaccines and date of vaccinations. | |
| | Sweden | *The National Vaccination Register.* Defined according to the specific administered COVID-19 vaccines and date of vaccinations. | |
| Documented SARS-CoV-2 infection | Denmark | *The Danish Microbiology Database.* Defined as the date of registered positive PCR test for SARS-CoV-2. | Binary: yes/no |
| | Finland | *National Infectious Diseases Register.* Defined as the date of registered positive PCR test for SARS-CoV-2. | |

| VARIABLE | COUNTRY | DATA SOURCE AND DETAILS | VALUES/CODES |
|---|---|---|---|
| | Norway | *Norwegian Surveillance System for Communicable Diseases (MSIS).*<br>Defined as the date of registered positive PCR test for SARS CoV-2. | |
| | Sweden | *Register on surveillance of notifiable communicable diseases (SmiNet).*<br>Defined as the date of registered positive PCR test for SARS-CoV-2. | |
| Hospitalisation for COVID-19 | Denmark | *The National Patient Register and the Danish Microbiology Database.*<br>Defined as a hospitalisation on the day of, within 14 days of or in the two days after a PCR positive test for SARS-CoV-2, b) inpatient contact or at least 12 hours of contact, c) a COVID-19 relevant diagnosis code (ICD-10: B342, B342A, B948A, B972, B972A, B972B, B972B1, Z038PA1) | Binary: yes/no |
| | Finland | *National Care Register for Health Care and the National Infectious Diseases Register.*<br>Defined as a hospitalisation on the day of, within 14 days of or in the two days after a PCR positive test for SARS-CoV-2, b) inpatient hospital contact, and c) a COVID-19 relevant main diagnosis (ICD-10: J00-J22, J46, J80-J84, J851, J86, U071, U072). | |
| | Norway | *The Norwegian Intensive Care and Pandemic Registry (NIPaR)*<br>Defined as an individual with a positive PCR test for SARS-CoV-2 who were inpatient hospitalised and where COVID-19 was registered as the main cause of hospitalisation. | |
| | Sweden | *The Swedish Patient Register and the Register on surveillance of notifiable communicable diseases (SmiNet).*<br>Defined as a hospitalisation on the day of, within 14 days of or in the two days after a PCR positive test for SARS-CoV-2, b) inpatient contact or at least 12 hours of contact, c) a COVID-19 relevant diagnosis code (ICD-10: U071, U072, U109) | |
| Intensive care unit admission for COVID-19 | Denmark | *The National Patient Register and the Danish Microbiology Database.*<br>Defined as admission to an intensive care unit facility during hospitalisation for COVID-19. | Binary: yes/no |
| | Finland | *Finnish Intensive Care Consortium's Quality Register for Intensive Care, National Care Register for Health Care and the National Infectious Diseases Register.*<br>Defined as admission to an intensive care unit facility during hospitalisation for COVID-19. | |
| | Norway | *The Norwegian Intensive Care and Pandemic Registry (NIPaR)*<br>Individuals who have tested positive for SARS-CoV-2 and are admitted to an intensive care unit (ICU). Patients are registered as ICU patients if they fulfil one of five categories:<br>1.Length of stay over 24 hours in intensive care<br>2.Require mechanical ventilation<br>3.Are transferred between intensive care wards<br>4.Persistent administration of vasoactive medication<br>5.Length of stay under 24 hours, but passed away during stay in intensive care | |
| | Sweden | *The Swedish Patient Register and the Register on surveillance of notifiable communicable diseases (SmiNet).* | |

| VARIABLE | COUNTRY | DATA SOURCE AND DETAILS | VALUES/CODES |
|---|---|---|---|
| | | Defined as admission to an intensive care unit facility during hospitalisation for COVID-19. | |
| COVID-19 death | Denmark | *The Civil Registration System and the Danish Microbiology Database.* Defined as (the date of) death within 30 days after PCR positive test for SARS-CoV-2. | Binary: yes/no |
| | Finland | *The Finnish Population Information System and the National Infectious Diseases Register.* Defined as (the date of) death within 30 days after PCR positive test for SARS-CoV-2. | |
| | Norway | *Norwegian Population Register and the Norwegian Surveillance System for Communicable Diseases (MSIS).* Defined as (the date of) death with a registered ICD-10 code of U071, U072, U109, or U099 as the main or contributing cause of death. | |
| | Sweden | *The Total Population Register, the Cause of Death Register, and the Swedish Patient Register and the Register on surveillance of notifiable communicable diseases (SmiNet).* Defined as (the date of) death within 30 days after PCR positive test for SARS-CoV-2. | |

[a]To account for the risk of severe COVID-19, we adjusted for vaccine priority groups, specifically established for each country. In Denmark, the COVID-19 vaccine priority groups were governmentally assigned and individuals were prioritized according to the risk of severe infection as well as whether being health and social care workers. In the remaining countries, vulnerable individuals (such as those receiving nursing care or living in nursing homes) and healthcare personnel were identified. [b]Autoimmunity related conditions (AIC) includes diagnoses of disorders such as inflammatory bowel diseases, diseases involving the blood, immune mechanism or endocrine systems, inflammatory rheumatic diseases, psoriasis, lupus erythematosus, multiple sclerosis; subject to country-specific definitions. [c]We only considered receive of a third dose as a booster dose if the third dose was administered at least 90 days after the day of the second vaccine dose of the primary vaccine schedule (as shorter intervals likely reflect an additional dose within the primary schedule among immunocompromised individuals).

**Supplementary Table 3. Description of ethical regulations within each country.**

| Country | Ethical Regulations |
| --- | --- |
| Denmark | The Danish study was performed as a surveillance study as part of the governmental institution Statens Serum Institut's (SSI) advisory tasks for the Danish Ministry of Health. SSI's purpose is to monitor and fight the spread of disease in accordance with section 222 of the Danish Health Act. According to Danish law, national surveillance activities conducted by SSI do not require approval from an ethics committee. It was approved by the Danish Governmental law firm and SSI's compliance department that the study is fully compliant with all legal, ethical and IT-security requirements and there are no further approval procedures regarding such studies. |
| Finland | By Finnish law, the Finnish Institute for Health and Welfare (THL) is the national expert institution to carry out surveillance on the impact of vaccinations in Finland (Communicable Diseases Act, https://www.finlex.fi/en/laki/kaannokset/2016/en20161227.pdf). Neither specific ethical approval (a waiver of ethical approval was received from Professor Mika Salminen, Director of the Department for Health Security Finnish Institute for Health and Welfare) of this study nor informed consent from the participants was needed. |
| Norway | The Norwegian study was approved by the Norwegian Regional Committee for Health Research Ethics South East (REK Sør-Øst A, ref 122745), and has conformed to the principles embodied in the Declaration of Helsinki. The emergency preparedness register was established according to the Health Preparedness Act §2-4. Consent to participate was not applicable as this is a register-based study. |
| Sweden | The Swedish study is approved by the Swedish Ethical Review Authority (2020-06859, 2021-02186) and has conformed to the principles embodied in the Declaration of Helsinki. Consent to participate is not applicable as this is a register-based study. |

**Supplementary Table 4. Number of individuals excluded because of previous history of SARS-CoV-2 infection by comparison (matched analyses) and country.**

| Schedule | Denmark | Finland | Norway | Sweden |
|---|---|---|---|---|
| AZD1AZD2BNT3 vs AZD1AZD2 | 338 | 3090 | 132 | 61874 |
| AZD1AZD2MOD3 vs AZD1AZD2 | 209 | 2596 | 109 | 47851 |
| AZD1BNT2BNT3 vs AZD1BNT2 | 26350 | 4944 | 8113 | 33507 |
| AZD1MOD2MOD3 vs AZD1MOD2 | 9840 | 725 | 652 | 4641 |
| BNT1BNT2MOD3 vs BNT1BNT2 | 368178 | 102006 | 101487 | 864409 |
| MOD1MOD2BNT3 vs MOD1MOD2 | 56236 | 9957 | 22362 | 152384 |
| BNT1MOD2MOD3 vs BNT1MOD2 | 0 | 2428 | 22411 | 5708 |
| MOD1BNT2BNT3 vs MOD1BNT2 | 29 | 2331 | 8571 | 13031 |
| BNT1MOD2BNT3 vs BNT1MOD2 | 0 | 2458 | 22752 | 6222 |
| MOD1BNT2MOD3 vs MOD1BNT2 | 0 | 2309 | 8546 | 12609 |
| BNT1BNT2BNT3 vs BNT1BNT2 | 614648 | 112994 | 109249 | 987077 |
| MOD1MOD2MOD3 vs MOD1MOD2 | 84732 | 10413 | 22461 | 150829 |

**Supplementary Table 5. Number of individuals excluded because of previous history of SARS-CoV-2 infection by comparison (weighted analyses) and country.**

| | Studied schedule | Comparison schedule |
|---|---|---|
| AZD1AZD2BNT3 vs BNT1BNT2BNT3 | | |
| Denmark | 66 (6.6%) | 101498 (5.7%) |
| Norway | 26 (3.4%) | 7372 (0.6%) |
| Finland | 328 (0.3%) | 950 (0.3%) |
| Sweden | 15158 (4.1%) | 84015 (6.5%) |
| AZD1AZD2MOD3 vs MOD1MOD2MOD3 | | |
| Denmark | <5 | 12449 (5.0%) |
| Norway | <5 | 826 (0.6%) |
| Finland | 106 (0.2%) | 96 (0.3%) |
| Sweden | 6232 (4.9%) | 11550 (8.9%) |
| AZD1BNT2BNT3 vs BNT1BNT2BNT3 | | |
| Denmark | 8764 (11.2%) | 81257 (6.1%) |
| Norway | 1074 (1.1%) | 3890 (0.5%) |
| Finland | 578 (0.6%) | 4882 (0.6%) |
| Sweden | 9092 (13.7%) | 134969 (9.3%) |
| AZD1MOD2MOD3 vs MOD1MOD2MOD3 | | |
| Denmark | 3204 (7.4%) | 7889 (4.7%) |
| Norway | 29 (2.6%) | 608 (0.5%) |
| Finland | 73 (0.4%) | 466 (0.4%) |
| Sweden | 758 (14.0%) | 16272 (11.2%) |
| BNT1BNT2MOD3 vs BNT1BNT2BNT3 | | |
| Denmark | 28 (4.8%) | 122192 (5.1%) |
| Norway | 1788 (0.6%) | 6632 (0.5%) |
| Finland | 2535 (0.6%) | 7440 (0.6%) |
| Sweden | 94793 (11.1%) | 142222 (7.6%) |
| MOD1MOD2BNT3 vs BNT1BNT2BNT3 | | |
| Denmark | 46 (4.7%) | 125246 (4.8%) |
| Norway | 814 (0.7%) | 8399 (0.6%) |
| Finland | 370 (0.6%) | 8643 (0.6%) |
| Sweden | 20585 (10.1%) | 178885 (8.3%) |
| BNT1MOD2MOD3 vs BNT1BNT2BNT3 | | |
| Denmark | | 118164 (4.9%) |
| Norway | 291 (0.3%) | 2114 (0.4%) |
| Finland | 65 (0.6%) | 7618 (0.8%) |
| Sweden | 159 (16.5%) | 142842 (8.1%) |
| MOD1BNT2BNT3 vs BNT1BNT2BNT3 | | |
| Denmark | <5 | 127660 (4.9%) |
| Norway | 59 (0.5%) | 2817 (0.5%) |
| Finland | 37 (0.8%) | 8885 (0.7%) |
| Sweden | 465 (17.3%) | 184932 (8.6%) |
| BNT1MOD2BNT3 vs BNT1BNT2BNT3 | | |
| Denmark | | 120238 (5.0%) |
| Norway | 604 (0.3%) | 2472 (0.5%) |
| Finland | 95 (0.9%) | 8004 (0.8%) |
| Sweden | 575 (16.6%) | 158670 (11.1%) |
| MOD1BNT2MOD3 vs BNT1BNT2BNT3 | | |
| Denmark | <5 | 119040 (4.8%) |
| Norway | 39 (0.5%) | 2379 (0.4%) |
| Finland | 18 (0.8%) | 7677 (0.8%) |
| Sweden | 101 (18.9%) | 145104 (7.8%) |
| MOD1MOD2MOD3 vs BNT1BNT2BNT3 | | |
| Denmark | 14886 (4.3%) | 117653 (4.9%) |
| Norway | 816 (0.6%) | 6436 (0.5%) |
| Finland | 630 (0.4%) | 7608 (0.6%) |
| Sweden | 19557 (9.7%) | 144773 (7.5%) |

**Supplementary Table 6. Distribution of age, sex, and calendar period by comparison and country.**

| | Heterologous booster schedule (studied schedule) | | | | Primary schedule or homologous booster schedule (comparison schedule) | | | |
|---|---|---|---|---|---|---|---|---|
| | **Total individuals** | **Age (mean, SD)** | **Female sex (%)** | **Calendar period (min-max)** | **Total individuals** | **Age (mean, SD)** | **Female sex (%)** | **Calendar period (min-max)** |
| **Heterologous booster- vs primary schedules** | | | | | | | | |
| AZD1AZD2BNT3 vs AZD1AZD2 | | | | | | | | |
| Denmark | 218 | 43 (13.5) | 50.9% | 11/11/21 - 28/02/22 | 207 | 43.3 (13.4) | 48.3% | 11/11/21 - 28/02/22 |
| Finland | 46085 | 68.9 (3.8) | 50% | 18/10/21 - 28/02/22 | 46003 | 68.9 (3.8) | 50% | 17/10/21 - 28/02/22 |
| Norway | 565 | 42.5 (15.6) | 44.6% | 14/10/21 - 28/02/22 | 564 | 42.6 (15.6) | 42.9% | 14/10/21 - 28/02/22 |
| Sweden | 63120 | 64.5 (15.5) | 56.4% | 21/10/21 - 28/02/22 | 62243 | 64.5 (15.5) | 52.7% | 21/10/21 - 28/02/22 |
| AZD1AZD2MOD3 vs AZD1AZD2 | | | | | | | | |
| Denmark | 32 | 51.2 (9.8) | 50% | 18/11/21 - 28/02/22 | 32 | 51.8 (9.9) | 31.2% | 18/11/21 - 28/02/22 |
| Finland | 29718 | 69.1 (3.8) | 49.9% | 18/10/21 - 28/02/22 | 29698 | 69.1 (3.8) | 49.4% | 18/10/21 - 28/02/22 |
| Norway | 214 | 48.5 (14.1) | 41.1% | 27/10/21 - 28/02/22 | 213 | 48.3 (14.1) | 41.8% | 27/10/21 - 28/02/22 |
| Sweden | 50634 | 67.5 (11.4) | 52.8% | 21/10/21 - 28/02/22 | 50169 | 67.6 (11.4) | 51.2% | 21/10/21 - 28/02/22 |
| AZD1BNT2BNT3 vs AZD1BNT2 | | | | | | | | |
| Denmark | 6748 | 37.6 (11.9) | 78.2% | 15/10/21 - 28/02/22 | 5765 | 37 (11.7) | 76.6% | 14/10/21 - 28/02/22 |
| Finland | 28175 | 56.5 (13.4) | 53.5% | 18/10/21 - 28/02/22 | 27969 | 56.5 (13.4) | 55.2% | 17/10/21 - 28/02/22 |
| Norway | 21069 | 37.4 (12.9) | 80.3% | 14/10/21 - 28/02/22 | 21032 | 37.3 (12.8) | 77.3% | 14/10/21 - 28/02/22 |
| Sweden | 26365 | 42.8 (14.5) | 75.3% | 21/10/21 - 28/02/22 | 26083 | 42.8 (14.5) | 72.6% | 21/10/21 - 28/02/22 |
| AZD1MOD2MOD3 vs AZD1MOD2 | | | | | | | | |
| Denmark | 4329 | 38.5 (12.1) | 81.7% | 14/10/21 - 28/02/22 | 3912 | 38.2 (12) | 79.2% | 14/10/21 - 28/02/22 |
| Finland | 6806 | 55.7 (13.4) | 56.9% | 18/10/21 - 28/02/22 | 6796 | 55.7 (13.4) | 54.9% | 17/10/21 - 28/02/22 |
| Norway | 438 | 48.5 (13.5) | 65.5% | 26/10/21 - 28/02/22 | 438 | 48.2 (13.4) | 62.3% | 26/10/21 - 28/02/22 |
| Sweden | 2450 | 48 (12.6) | 74.7% | 25/10/21 - 28/02/22 | 2428 | 47.8 (12.7) | 73.8% | 25/10/21 - 28/02/22 |
| BNT1BNT2MOD3 vs BNT1BNT2 | | | | | | | | |
| Denmark | 334 | 49.2 (16.8) | 52.4% | 26/10/21 - 28/02/22 | 330 | 49.3 (16.7) | 49.1% | 26/10/21 - 28/02/22 |
| Finland | 348639 | 52.8 (15) | 54% | 18/10/21 - 28/02/22 | 348641 | 52.7 (15) | 48.4% | 17/10/21 - 28/02/22 |
| Norway | 238921 | 49.8 (12.2) | 49.3% | 14/10/21 - 28/02/22 | 237807 | 49.8 (12.3) | 48.2% | 14/10/21 - 28/02/22 |
| Sweden | 504359 | 52.3 (12.8) | 47.8% | 21/10/21 - 28/02/22 | 501571 | 52.2 (12.8) | 46.3% | 21/10/21 - 28/02/22 |

| | Heterologous booster schedule (studied schedule) | | | | Primary schedule or homologous booster schedule (comparison schedule) | | | |
|---|---|---|---|---|---|---|---|---|
| | **Total individuals** | **Age (mean, SD)** | **Female sex (%)** | **Calendar period (min-max)** | **Total individuals** | **Age (mean, SD)** | **Female sex (%)** | **Calendar period (min-max)** |
| MOD1MOD2BNT3 vs MOD1MOD2 | | | | | | | | |
| Denmark | 600 | 49.6 (21.3) | 54.8% | 17/10/21 - 28/02/22 | 588 | 49.4 (21.1) | 50% | 17/10/21 - 28/02/22 |
| Finland | 50415 | 52.2 (18.3) | 48.7% | 18/10/21 - 28/02/22 | 50384 | 52.2 (18.3) | 49.4% | 17/10/21 - 28/02/22 |
| Norway | 64393 | 40.9 (15.2) | 49.6% | 14/10/21 - 28/02/22 | 63940 | 40.9 (15.1) | 48.3% | 14/10/21 - 28/02/22 |
| Sweden | 105725 | 44 (18) | 50.2% | 21/10/21 - 28/02/22 | 105134 | 43.9 (17.9) | 47.9% | 21/10/21 - 28/02/22 |
| BNT1MOD2MOD3 vs BNT1MOD2 | | | | | | | | |
| Denmark | <5 | 45 (37.5) | 33.3% | 02/11/21 - 20/02/22 | <5 | 45.7 (37.9) | 0% | 02/11/21 - 08/02/22 |
| Finland | 7393 | 42 (13.9) | 60.4% | 20/10/21 - 28/02/22 | 7391 | 41.9 (13.9) | 46.4% | 20/10/21 - 28/02/22 |
| Norway | 71546 | 43.4 (9.5) | 45.2% | 02/11/21 - 28/02/22 | 71035 | 43.3 (9.7) | 42.4% | 02/11/21 - 28/02/22 |
| Sweden | 609 | 43.6 (10.7) | 43.7% | 23/11/21 - 28/02/22 | 608 | 43.5 (10.8) | 42.9% | 23/11/21 - 28/02/22 |
| MOD1BNT2BNT3 vs MOD1BNT2 | | | | | | | | |
| Denmark | 10 | 40 (16.4) | 40% | 08/12/21 - 28/02/22 | 10 | 41 (15.9) | 40% | 08/12/21 - 28/02/22 |
| Finland | 3087 | 45.6 (18.8) | 46.2% | 18/10/21 - 28/02/22 | 3084 | 45.6 (18.8) | 44.7% | 18/10/21 - 28/02/22 |
| Norway | 7859 | 32.8 (11.3) | 45.9% | 27/10/21 - 28/02/22 | 7793 | 32.8 (11.2) | 44.5% | 27/10/21 - 28/02/22 |
| Sweden | 1586 | 33.7 (14.6) | 49.4% | 24/10/21 - 28/02/22 | 1583 | 33.7 (14.6) | 41.8% | 24/10/21 - 28/02/22 |
| BNT1MOD2BNT3 vs BNT1MOD2 | | | | | | | | |
| Denmark | | | | | | | | |
| Finland | 8565 | 38.1 (14.2) | 46.7% | 26/10/21 - 28/02/22 | 8562 | 38.1 (14.2) | 46.7% | 24/10/21 - 28/02/22 |
| Norway | 141657 | 34.4 (11.8) | 45.2% | 02/11/21 - 28/02/22 | 140531 | 34.4 (11.8) | 42.6% | 02/11/21 - 28/02/22 |
| Sweden | 2243 | 34.2 (12.7) | 48% | 15/11/21 - 28/02/22 | 2240 | 34.2 (12.6) | 44.7% | 15/11/21 - 28/02/22 |
| MOD1BNT2MOD3 vs MOD1BNT2 | | | | | | | | |
| Denmark | | | | | | | | |
| Finland | 1755 | 46.9 (15.6) | 57.5% | 19/10/21 - 28/02/22 | 1755 | 46.9 (15.7) | 46.7% | 19/10/21 - 28/02/22 |
| Norway | 4249 | 40.9 (10.4) | 46.4% | 23/11/21 - 28/02/22 | 4205 | 40.7 (10.6) | 45.8% | 23/11/21 - 28/02/22 |
| Sweden | 327 | 48.5 (11.4) | 47.4% | 21/10/21 - 28/02/22 | 327 | 48.4 (11.6) | 43.7% | 21/10/21 - 28/02/22 |
| BNT1BNT2BNT3 vs BNT1BNT2 | | | | | | | | |
| Denmark | 430713 | 42.6 (16.1) | 49.6% | 14/10/21 - 28/02/22 | 422554 | 42.3 (15.9) | 45.9% | 14/10/21 - 28/02/22 |

| | Heterologous booster schedule (studied schedule) | | | | Primary schedule or homologous booster schedule (comparison schedule) | | | |
|---|---|---|---|---|---|---|---|---|
| | **Total individuals** | **Age (mean, SD)** | **Female sex (%)** | **Calendar period (min-max)** | **Total individuals** | **Age (mean, SD)** | **Female sex (%)** | **Calendar period (min-max)** |
| Finland | 680361 | 51.7 (17.3) | 52.4% | 18/10/21 - 28/02/22 | 679394 | 51.6 (17.3) | 50% | 17/10/21 - 28/02/22 |
| Norway | 482290 | 45.5 (16.7) | 50.9% | 14/10/21 - 28/02/22 | 478944 | 45.4 (16.5) | 48.9% | 14/10/21 - 28/02/22 |
| Sweden | 791382 | 48.1 (16.4) | 49.9% | 21/10/21 - 28/02/22 | 784699 | 47.9 (16.3) | 49% | 21/10/21 - 28/02/22 |
| MOD1MOD2MOD3 vs MOD1MOD2 | | | | | | | | |
| Denmark | 98164 | 37.5 (12.9) | 47.6% | 14/10/21 - 28/02/22 | 96827 | 37.3 (12.7) | 45.6% | 14/10/21 - 28/02/22 |
| Finland | 81314 | 54.4 (16.8) | 54.9% | 18/10/21 - 28/02/22 | 81233 | 54.3 (16.8) | 49.3% | 17/10/21 - 28/02/22 |
| Norway | 63974 | 46.3 (12.9) | 48.5% | 14/10/21 - 28/02/22 | 63591 | 46.1 (12.9) | 47.5% | 14/10/21 - 28/02/22 |
| Sweden | 93241 | 51.2 (13.3) | 48% | 21/10/21 - 28/02/22 | 92640 | 51 (13.4) | 46.2% | 21/10/21 - 28/02/22 |
| **Heterologous- vs homologous booster schedules** | | | | | | | | |
| AZD1AZD2BNT3 vs BNT1BNT2BNT3 | | | | | | | | |
| Denmark | 930 | 45.8 (12.5) | 62.7% | 29/10/21 - 22/01/22 | 1686091 | 50.4 (12.7) | 48.9% | 29/10/21 - 22/01/22 |
| Finland | 113620 | 68.4 (2.6) | 49.4% | 09/11/21 - 25/01/22 | 343376 | 72.4 (3.1) | 55.3% | 09/11/21 - 25/01/22 |
| Norway | 740 | 43.9 (15.6) | 43.9% | 04/11/21 - 09/02/22 | 1309526 | 54 (15.4) | 51% | 04/11/21 - 09/02/22 |
| Sweden | 355948 | 70.5 (7.8) | 52.4% | 11/11/21 - 17/01/22 | 1209728 | 63.6 (10.9) | 52.6% | 11/11/21 - 17/01/22 |
| AZD1AZD2MOD3 vs MOD1MOD2MOD3 | | | | | | | | |
| Denmark | 60 | 50.8 (9.7) | 51.7% | 09/11/21 - 06/02/22 | 237355 | 41.5 (11.5) | 46.4% | 09/11/21 - 06/02/22 |
| Finland | 47168 | 68.7 (2.9) | 49.8% | 19/11/21 - 29/01/22 | 37383 | 71.9 (3.3) | 52.9% | 19/11/21 - 29/01/22 |
| Norway | 246 | 50.6 (13.4) | 40.7% | 14/10/21 - 05/02/22 | 144180 | 51.8 (12.4) | 48.1% | 14/10/21 - 05/02/22 |
| Sweden | 120874 | 69.7 (7.4) | 51.2% | 11/11/21 - 27/01/22 | 118024 | 59.7 (11.9) | 49.3% | 11/11/21 - 27/01/22 |
| AZD1BNT2BNT3 vs BNT1BNT2BNT3 | | | | | | | | |
| Denmark | 69558 | 44.9 (12) | 79.5% | 08/10/21 - 05/01/22 | 1254153 | 48.9 (10.6) | 49.9% | 08/10/21 - 05/01/22 |
| Finland | 101326 | 58.4 (9.2) | 52.5% | 15/10/21 - 22/01/22 | 800650 | 53.6 (11.8) | 53.8% | 15/10/21 - 22/01/22 |
| Norway | 96725 | 44.7 (12.6) | 78% | 16/11/21 - 19/01/22 | 752266 | 47.9 (12.5) | 51.7% | 16/11/21 - 19/01/22 |
| Sweden | 57041 | 47 (13.3) | 77.8% | 15/11/21 - 01/02/22 | 1318686 | 54.9 (13.7) | 51.7% | 15/11/21 - 01/02/22 |
| AZD1MOD2MOD3 vs MOD1MOD2MOD3 | | | | | | | | |
| Denmark | 40069 | 46.6 (11.4) | 82.2% | 07/10/21 - 07/01/22 | 159710 | 43.8 (12) | 46.9% | 07/10/21 - 07/01/22 |

| | Heterologous booster schedule (studied schedule) | | | | Primary schedule or homologous booster schedule (comparison schedule) | | | |
|---|---|---|---|---|---|---|---|---|
| | **Total individuals** | **Age (mean, SD)** | **Female sex (%)** | **Calendar period (min-max)** | **Total individuals** | **Age (mean, SD)** | **Female sex (%)** | **Calendar period (min-max)** |
| Finland | 18738 | 58.1 (9.8) | 53.2% | 14/10/21 - 25/01/22 | 104355 | 55 (11.4) | 50.8% | 14/10/21 - 25/01/22 |
| Norway | 1077 | 51.5 (11.5) | 64.8% | 17/10/21 - 31/01/22 | 114926 | 48.3 (10.5) | 47.7% | 17/10/21 - 31/01/22 |
| Sweden | 4656 | 50 (10.9) | 78.1% | 15/11/21 - 03/02/22 | 128714 | 52.9 (11.6) | 48.3% | 15/11/21 - 03/02/22 |
| BNT1BNT2MOD3 vs BNT1BNT2BNT3 | | | | | | | | |
| Denmark | 553 | 49.8 (15.5) | 52.1% | 06/10/21 - 01/02/22 | 2282795 | 55.3 (15.1) | 50.3% | 01/10/21 - 01/02/22 |
| Finland | 443176 | 53.5 (13.2) | 52.2% | 25/11/21 - 04/02/22 | 1203555 | 57.1 (15.4) | 53% | 25/11/21 - 04/02/22 |
| Norway | 320119 | 51.8 (11.2) | 48.4% | 26/10/21 - 01/02/22 | 1248035 | 58.1 (13.1) | 50.9% | 26/10/21 - 01/02/22 |
| Sweden | 756029 | 54.4 (12.9) | 47.9% | 21/10/21 - 10/02/22 | 1720151 | 62.7 (13.5) | 52.3% | 21/10/21 - 10/02/22 |
| MOD1MOD2BNT3 vs BNT1BNT2BNT3 | | | | | | | | |
| Denmark | 936 | 51.2 (20.8) | 53.3% | 01/10/21 - 31/01/22 | 2461534 | 57.5 (16.5) | 50.9% | 01/10/21 - 31/01/22 |
| Finland | 61418 | 54.5 (17.7) | 49.5% | 17/11/21 - 09/02/22 | 1367687 | 58 (17.2) | 53.5% | 17/11/21 - 09/02/22 |
| Norway | 120544 | 47.9 (18.3) | 53.9% | 19/10/21 - 08/02/22 | 1497941 | 57.2 (16.5) | 51.4% | 19/10/21 - 08/02/22 |
| Sweden | 183788 | 53.6 (20.5) | 52.3% | 19/10/21 - 10/02/22 | 1975964 | 59.1 (17.6) | 52.7% | 19/10/21 - 10/02/22 |
| BNT1MOD2MOD3 vs BNT1BNT2BNT3 | | | | | | | | |
| Denmark | 51 | 57.1 (23.1) | 51% | 25/10/21 - 26/01/22 | 2316309 | 57.3 (15.8) | 50.5% | 25/10/21 - 26/01/22 |
| Finland | 10277 | 43.5 (12.1) | 57.1% | 19/12/21 - 10/02/22 | 940130 | 50.9 (14.6) | 51.2% | 19/12/21 - 10/02/22 |
| Norway | 97079 | 44.2 (8.4) | 44% | 20/12/21 - 09/02/22 | 488451 | 47 (9.4) | 48.2% | 20/12/21 - 09/02/22 |
| Sweden | 806 | 45.6 (11.5) | 43.3% | 31/10/21 - 12/02/22 | 1627528 | 60.8 (13) | 52% | 27/10/21 - 12/02/22 |
| MOD1BNT2BNT3 vs BNT1BNT2BNT3 | | | | | | | | |
| Denmark | 90 | 62.3 (21.9) | 38.9% | 04/10/21 - 02/02/22 | 2488348 | 57.1 (16.8) | 50.9% | 01/10/21 - 02/02/22 |
| Finland | 4504 | 50.9 (18.7) | 48% | 24/11/21 - 11/02/22 | 1324944 | 56.6 (16.9) | 53.1% | 24/11/21 - 11/02/22 |
| Norway | 12620 | 32.4 (10.7) | 48.8% | 20/12/21 - 11/02/22 | 588157 | 41.9 (12.5) | 48.8% | 20/12/21 - 11/02/22 |
| Sweden | 2238 | 37.9 (17.9) | 51.8% | 20/10/21 - 14/02/22 | 1977303 | 58.1 (17.7) | 52.5% | 20/10/21 - 14/02/22 |
| BNT1MOD2BNT3 vs BNT1BNT2BNT3 | | | | | | | | |
| Denmark | 24 | 38.3 (20.1) | 50% | 01/11/21 - 09/02/22 | 2290559 | 56 (16) | 49.9% | 01/11/21 - 09/02/22 |
| Finland | 10148 | 38.9 (13.1) | 46.5% | 14/12/21 - 11/02/22 | 1018958 | 51.1 (14.7) | 51.7% | 14/12/21 - 11/02/22 |
| Norway | 194343 | 34.1 (11.2) | 45.8% | 28/12/21 - 10/02/22 | 513894 | 40 (12.7) | 48.6% | 28/12/21 - 10/02/22 |
| Sweden | 2884 | 35.1 (12.6) | 49% | 30/11/21 - 12/02/22 | 1267739 | 49.7 (14.9) | 50.6% | 30/11/21 - 12/02/22 |

| | Heterologous booster schedule (studied schedule) | | | | Primary schedule or homologous booster schedule (comparison schedule) | | | |
|---|---|---|---|---|---|---|---|---|
| | Total individuals | Age (mean, SD) | Female sex (%) | Calendar period (min-max) | Total individuals | Age (mean, SD) | Female sex (%) | Calendar period (min-max) |
| MOD1BNT2MOD3 vs BNT1BNT2BNT3 | | | | | | | | |
| Denmark | 14 | 59.2 (23) | 21.4% | 20/10/21 - 26/01/22 | 2370020 | 58.2 (15.4) | 50.8% | 01/10/21 - 26/01/22 |
| Finland | 2102 | 48.2 (14.4) | 55.3% | 19/12/21 - 10/02/22 | 961231 | 51.7 (15) | 51.3% | 19/12/21 - 10/02/22 |
| Norway | 7458 | 42.7 (8.9) | 45.9% | 21/12/21 - 10/02/22 | 532352 | 45.4 (11) | 48.2% | 21/12/21 - 10/02/22 |
| Sweden | 433 | 50.1 (12.5) | 49.2% | 21/10/21 - 13/02/22 | 1709913 | 61.9 (13.5) | 52.2% | 21/10/21 - 13/02/22 |
| MOD1MOD2MOD3 vs BNT1BNT2BNT3 | | | | | | | | |
| Denmark | 332127 | 49.2 (18.6) | 48.3% | 28/10/21 - 28/01/22 | 2287611 | 56.9 (15.7) | 50.2% | 28/10/21 - 28/01/22 |
| Finland | 150242 | 59.6 (15.2) | 52.7% | 17/11/21 - 05/02/22 | 1312426 | 59.6 (16.2) | 53.8% | 17/11/21 - 05/02/22 |
| Norway | 141956 | 52.4 (12.2) | 47.8% | 12/11/21 - 04/02/22 | 1218904 | 57.1 (13.3) | 50.7% | 12/11/21 - 04/02/22 |
| Sweden | 182263 | 58.9 (15.3) | 49.4% | 12/10/21 - 10/02/22 | 1782303 | 63.3 (13.8) | 52.5% | 12/10/21 - 10/02/22 |

NE denotes not estimated and SD standard deviation.

**Supplementary Table 7. Distribution of vaccine priority groups and comorbidities by comparison in Denmark.**

| | Total individuals | Priority groups | | | | Comorbidities | | | | |
|---|---|---|---|---|---|---|---|---|---|---|
| | | Age priority | At risk | Close contact | Healthcare workers | AIC | Cancer | CPD | CVD/DM | CKD |
| **Heterologous booster- vs primary schedules** | | | | | | | | | | |
| AZD1AZD2BNT3 vs AZD1AZD2 | | | | | | | | | | |
| Studied schedule | 218 | 151 (69.3%) | 0 | 0 | 67 (30.7%) | <5 | 144 (66.1%) | <5 | 0 | 0 |
| Comparison schedule | 207 | 150 (72.5%) | 0 | 0 | 57 (27.5%) | <5 | 121 (58.5%) | <5 | <5 | 0 |
| AZD1AZD2MOD3 vs AZD1AZD2 | | | | | | | | | | |
| Studied schedule | 32 | 27 (84.4%) | 0 | 0 | 5 (15.6%) | 0 | 17 (53.1%) | 0 | 0 | 0 |
| Comparison schedule | 32 | NE | NE | NE | NE | 0 | 17 (53.1%) | 0 | 0 | 0 |
| AZD1BNT2BNT3 vs AZD1BNT2 | | | | | | | | | | |
| Studied schedule | 6748 | *(15.8%) | <5 | 0 | *(84.2%) | 189 (2.8%) | 5015 (74.3%) | 86 (1.3%) | 84 (1.2%) | 7 (0.1%) |
| Comparison schedule | 5765 | 890 (15.4%) | 8 (0.1%) | 0 | 4867 (84.4%) | 117 (2.0%) | 4357 (75.6%) | 69 (1.2%) | 74 (1.3%) | 8 (0.1%) |
| AZD1MOD2MOD3 vs AZD1MOD2 | | | | | | | | | | |
| Studied schedule | 4329 | *(11.1%) | <5 | 0 | *(88.8%) | 129 (3.0%) | 3312 (76.5%) | 59 (1.4%) | 66 (1.5%) | 8 (0.2%) |
| Comparison schedule | 3912 | 469 (12.0%) | 7 (0.2%) | 0 | 3436 (87.8%) | 103 (2.6%) | 3031 (77.5%) | 58 (1.5%) | 50 (1.3%) | 8 (0.2%) |
| BNT1BNT2MOD3 vs BNT1BNT2 | | | | | | | | | | |
| Studied schedule | 334 | 314 (94.0%) | 15 (4.5%) | 0 | 5 (1.5%) | 7 (2.1%) | 206 (61.7%) | <5 | 9 (2.7%) | <5 |
| Comparison schedule | 330 | 295 (89.4%) | 14 (4.2%) | 0 | 21 (6.4%) | 22 (6.7%) | 237 (71.8%) | 12 (3.6%) | 24 (7.3%) | 6 (1.8%) |
| MOD1MOD2BNT3 vs MOD1MOD2 | | | | | | | | | | |
| Studied schedule | 600 | 563 (93.8%) | 15 (2.5%) | 0 | 22 (3.7%) | 19 (3.2%) | 430 (71.7%) | 12 (2.0%) | 34 (5.7%) | <5 |
| Comparison schedule | 588 | 541 (92.0%) | 15 (2.6%) | 0 | 32 (5.4%) | 17 (2.9%) | 397 (67.5%) | 14 (2.4%) | 31 (5.3%) | <5 |
| BNT1MOD2MOD3 vs BNT1MOD2 | | | | | | | | | | |
| Studied schedule | <5 | <5 | 0 | 0 | 0 | 0 | <5 | 0 | 0 | 0 |
| Comparison schedule | <5 | <5 | 0 | 0 | 0 | 0 | <5 | 0 | 0 | 0 |
| MOD1BNT2BNT3 vs MOD1BNT2 | | | | | | | | | | |
| Studied schedule | 10 | 10 (100.0%) | 0 | 0 | 0 | 0 | <5 | 0 | <5 | 0 |
| Comparison schedule | 10 | 10 (100.0%) | 0 | 0 | 0 | 0 | 5 (50.0%) | 0 | 0 | 0 |
| BNT1MOD2BNT3 vs BNT1MOD2 | | | | | | | | | | |
| Studied schedule | 0 | 0 | 0 | 0 | 0 | 0 | 0 | 0 | 0 | 0 |

| | Total individuals | Priority groups | | | | Comorbidities | | | | |
|---|---|---|---|---|---|---|---|---|---|---|
| | | Age priority | At risk | Close contact | Healthcare workers | AIC | Cancer | CPD | CVD/DM | CKD |
| Comparison schedule | 0 | 0 | 0 | 0 | 0 | 0 | 0 | 0 | 0 | 0 |
| MOD1BNT2MOD3 vs MOD1BNT2 | | | | | | | | | | |
| Studied schedule | 0 | 0 | 0 | 0 | 0 | 0 | 0 | 0 | 0 | 0 |
| Comparison schedule | 0 | 0 | 0 | 0 | 0 | 0 | 0 | 0 | 0 | 0 |
| BNT1BNT2BNT3 vs BNT1BNT2 | | | | | | | | | | |
| Studied schedule | 430713 | *(94.4%) | *(1.2%) | <5 | *(4.4%) | 12207 (2.8%) | 300969 (69.9%) | 6813 (1.6%) | 14460 (3.4%) | 1901 (0.4%) |
| Comparison schedule | 422554 | 393688 (93.2%) | 7482 (1.8%) | 0 | 21384 (5.1%) | 9769 (2.3%) | 289015 (68.4%) | 6736 (1.6%) | 13102 (3.1%) | 1887 (0.4%) |
| MOD1MOD2MOD3 vs MOD1MOD2 | | | | | | | | | | |
| Studied schedule | 98164 | 94258 (96.0%) | 240 (0.2%) | 0 | 3666 (3.7%) | 2100 (2.1%) | 64319 (65.5%) | 1044 (1.1%) | 1997 (2.0%) | 271 (0.3%) |
| Comparison schedule | 96827 | 92036 (95.1%) | 450 (0.5%) | 0 | 4341 (4.5%) | 1716 (1.8%) | 63816 (65.9%) | 977 (1.0%) | 1824 (1.9%) | 250 (0.3%) |
| **Heterologous- vs homologous booster schedules** | | | | | | | | | | |
| AZD1AZD2BNT3 vs BNT1BNT2BNT3 | | | | | | | | | | |
| Studied schedule | 930 | NE | NE | NE | NE | 26 (2.8%) | 680 (73.1%) | 12 (1.3%) | 20 (2.2%) | <5 |
| Comparison schedule | 1686091 | 1556480 (92.3%) | 22258 (1.3%) | 9 (0.0%) | 107344 (6.4%) | 53699 (3.2%) | 1261062 (74.8%) | 31043 (1.8%) | 67961 (4.0%) | 7600 (0.5%) |
| AZD1AZD2MOD3 vs MOD1MOD2MOD3 | | | | | | | | | | |
| Studied schedule | 60 | 45 (75.0%) | 0 | 0 | 15 (25.0%) | 0 | 32 (53.3%) | <5 | <5 | 0 |
| Comparison schedule | 237355 | 226446 (95.4%) | 598 (0.3%) | 0 | 10311 (4.3%) | 5216 (2.2%) | 162348 (68.4%) | 2606 (1.1%) | 5062 (2.1%) | 552 (0.2%) |
| AZD1BNT2BNT3 vs BNT1BNT2BNT3 | | | | | | | | | | |
| Studied schedule | 69558 | 8134 (11.7%) | 31 (0.0%) | 0 | 61393 (88.3%) | 1902 (2.7%) | 54686 (78.6%) | 929 (1.3%) | 1349 (1.9%) | 140 (0.2%) |
| Comparison schedule | 1254153 | 1105827 (88.2%) | 22635 (1.8%) | 14 (0.0%) | 125677 (10.0%) | 44815 (3.6%) | 936430 (74.7%) | 24138 (1.9%) | 47036 (3.8%) | 5655 (0.5%) |
| AZD1MOD2MOD3 vs MOD1MOD2MOD3 | | | | | | | | | | |
| Studied schedule | 40069 | 3843 (9.6%) | 15 (0.0%) | 0 | 36211 (90.4%) | 1230 (3.1%) | 32776 (81.8%) | 555 (1.4%) | 920 (2.3%) | 106 (0.3%) |
| Comparison schedule | 159710 | 151665 (95.0%) | 587 (0.4%) | 0 | 7458 (4.7%) | 3936 (2.5%) | 111485 (69.8%) | 2070 (1.3%) | 4130 (2.6%) | 435 (0.3%) |
| BNT1BNT2MOD3 vs BNT1BNT2BNT3 | | | | | | | | | | |
| Studied schedule | 553 | 519 (93.9%) | 22 (4.0%) | 0 | 12 (2.2%) | 17 (3.1%) | 353 (63.8%) | 8 (1.4%) | 20 (3.6%) | 5 (0.9%) |
| Comparison schedule | 2282795 | 2053444 (90.0%) | 73217 (3.2%) | 28 (0.0%) | 156106 (6.8%) | 83328 (3.7%) | 1764184 (77.3%) | 59143 (2.6%) | 138437 (6.1%) | 17482 (0.8%) |
| MOD1MOD2BNT3 vs BNT1BNT2BNT3 | | | | | | | | | | |
| Studied schedule | 936 | 888 (94.9%) | 20 (2.1%) | 0 | 28 (3.0%) | 23 (2.5%) | 673 (71.9%) | 21 (2.2%) | 51 (5.4%) | 7 (0.7%) |

| | Total individuals | Priority groups | | | | Comorbidities | | | | |
|---|---|---|---|---|---|---|---|---|---|---|
| | | Age priority | At risk | Close contact | Healthcare workers | AIC | Cancer | CPD | CVD/DM | CKD |
| Comparison schedule | 2461534 | 2203538 (89.5%) | 101925 (4.1%) | 31 (0.0%) | 156040 (6.3%) | 90977 (3.7%) | 1921985 (78.1%) | 69171 (2.8%) | 171355 (7.0%) | 22824 (0.9%) |
| BNT1MOD2MOD3 vs BNT1BNT2BNT3 | | | | | | | | | | |
| Studied schedule | 51 | 46 (90.2%) | <5 | 0 | <5 | 0 | 30 (58.8%) | <5 | <5 | 0 |
| Comparison schedule | 2316309 | 2116090 (91.4%) | 73234 (3.2%) | 21 (0.0%) | 126964 (5.5%) | 80678 (3.5%) | 1800619 (77.7%) | 59572 (2.6%) | 150759 (6.5%) | 19004 (0.8%) |
| MOD1BNT2BNT3 vs BNT1BNT2BNT3 | | | | | | | | | | |
| Studied schedule | 90 | NE | NE | NE | NE | 5 (5.6%) | 69 (76.7%) | <5 | 9 (10.0%) | <5 |
| Comparison schedule | 2488348 | 2228591 (89.6%) | 102376 (4.1%) | 31 (0.0%) | 157350 (6.3%) | 91430 (3.7%) | 1938093 (77.9%) | 69536 (2.8%) | 171691 (6.9%) | 22887 (0.9%) |
| BNT1MOD2BNT3 vs BNT1BNT2BNT3 | | | | | | | | | | |
| Studied schedule | 24 | 18 (75.0%) | <5 | 0 | <5 | 0 | 17 (70.8%) | <5 | <5 | <5 |
| Comparison schedule | 2290559 | 2131310 (93.0%) | 51349 (2.2%) | 12 (0.0%) | 107888 (4.7%) | 75284 (3.3%) | 1759729 (76.8%) | 53341 (2.3%) | 135782 (5.9%) | 16501 (0.7%) |
| MOD1BNT2MOD3 vs BNT1BNT2BNT3 | | | | | | | | | | |
| Studied schedule | 14 | NE | NE | NE | NE | 0 | 12 (85.7%) | <5 | <5 | 0 |
| Comparison schedule | 2370020 | 2123352 (89.6%) | 94491 (4.0%) | 31 (0.0%) | 152146 (6.4%) | 89160 (3.8%) | 1862413 (78.6%) | 67418 (2.8%) | 166596 (7.0%) | 21964 (0.9%) |
| MOD1MOD2MOD3 vs BNT1BNT2BNT3 | | | | | | | | | | |
| Studied schedule | 332127 | 316696 (95.4%) | 3396 (1.0%) | 0 | 12035 (3.6%) | 8892 (2.7%) | 239644 (72.2%) | 6320 (1.9%) | 15364 (4.6%) | 1921 (0.6%) |
| Comparison schedule | 2287611 | 2111360 (92.3%) | 61458 (2.7%) | 15 (0.0%) | 114778 (5.0%) | 77547 (3.4%) | 1770398 (77.4%) | 56148 (2.5%) | 143192 (6.3%) | 17753 (0.8%) |

AID denotes autoimmunity related conditions, CPD chronic pulmonary disease, CVD/DM cardiovascular conditions and diabetes, CKD moderate to severe renal disease (chronic kidney disease), NE not estimable due national regulations on data privacy protection, and * that actual counts could not be presented (only percentage) due to national regulations on data privacy protection.

**Supplementary Table 8. Distribution of vaccine priority groups and comorbidities by comparison in Finland.**

| | | Priority groups | | | Comorbidities | | | | |
|---|---|---|---|---|---|---|---|---|---|
| | Total individuals | Others | Vulnerable | Healthcare workers | AIC | Cancer | CPD | CVD/DM | CKD |
| **Heterologous booster- vs primary schedules** | | | | | | | | | |
| AZD1AZD2BNT3 vs AZD1AZD2 | | | | | | | | | |
| Studied schedule | 46085 | 41375 (89.8%) | 64 (0.1%) | 4646 (10.1%) | 1911 (4.1%) | 4282 (9.3%) | 883 (1.9%) | 12482 (27.1%) | 458 (1.0%) |
| Comparison schedule | 46003 | 42375 (92.1%) | 89 (0.2%) | 3539 (7.7%) | 1579 (3.4%) | 4145 (9.0%) | 1116 (2.4%) | 12502 (27.2%) | 510 (1.1%) |
| AZD1AZD2MOD3 vs AZD1AZD2 | | | | | | | | | |
| Studied schedule | 29718 | 27005 (90.9%) | 36 (0.1%) | 2677 (9.0%) | 1038 (3.5%) | 2698 (9.1%) | 516 (1.7%) | 7970 (26.8%) | 284 (1.0%) |
| Comparison schedule | 29698 | 27452 (92.4%) | 66 (0.2%) | 2180 (7.3%) | 1026 (3.5%) | 2656 (8.9%) | 797 (2.7%) | 8252 (27.8%) | 353 (1.2%) |
| AZD1BNT2BNT3 vs AZD1BNT2 | | | | | | | | | |
| Studied schedule | 28175 | 22801 (80.9%) | 59 (0.2%) | 5315 (18.9%) | 1511 (5.4%) | 2312 (8.2%) | 625 (2.2%) | 10308 (36.6%) | 507 (1.8%) |
| Comparison schedule | 27969 | 22229 (79.5%) | 70 (0.3%) | 5670 (20.3%) | 1211 (4.3%) | 2152 (7.7%) | 680 (2.4%) | 9708 (34.7%) | 497 (1.8%) |
| AZD1MOD2MOD3 vs AZD1MOD2 | | | | | | | | | |
| Studied schedule | 6806 | 5371 (78.9%) | 9 (0.1%) | 1426 (21.0%) | 418 (6.1%) | 538 (7.9%) | 139 (2.0%) | 2426 (35.6%) | 119 (1.7%) |
| Comparison schedule | 6796 | 5374 (79.1%) | 14 (0.2%) | 1408 (20.7%) | 323 (4.8%) | 437 (6.4%) | 156 (2.3%) | 2374 (34.9%) | 112 (1.6%) |
| BNT1BNT2MOD3 vs BNT1BNT2 | | | | | | | | | |
| Studied schedule | 348639 | 310830 (89.2%) | 888 (0.3%) | 36921 (10.6%) | 8730 (2.5%) | 14453 (4.1%) | 2331 (0.7%) | 41241 (11.8%) | 1675 (0.5%) |
| Comparison schedule | 348641 | 316271 (90.7%) | 1686 (0.5%) | 30684 (8.8%) | 8810 (2.5%) | 14139 (4.1%) | 3107 (0.9%) | 42783 (12.3%) | 2002 (0.6%) |
| MOD1MOD2BNT3 vs MOD1MOD2 | | | | | | | | | |
| Studied schedule | 50415 | 44803 (88.9%) | 274 (0.5%) | 5338 (10.6%) | 1450 (2.9%) | 2357 (4.7%) | 477 (0.9%) | 7252 (14.4%) | 395 (0.8%) |
| Comparison schedule | 50384 | 45717 (90.7%) | 372 (0.7%) | 4295 (8.5%) | 1341 (2.7%) | 2245 (4.5%) | 590 (1.2%) | 7313 (14.5%) | 436 (0.9%) |
| BNT1MOD2MOD3 vs BNT1MOD2 | | | | | | | | | |
| Studied schedule | 7393 | 6636 (89.8%) | 11 (0.1%) | 746 (10.1%) | 106 (1.4%) | 164 (2.2%) | 18 (0.2%) | 377 (5.1%) | 24 (0.3%) |
| Comparison schedule | 7391 | 6643 (89.9%) | 28 (0.4%) | 720 (9.7%) | 112 (1.5%) | 148 (2.0%) | 26 (0.4%) | 416 (5.6%) | 26 (0.4%) |
| MOD1BNT2BNT3 vs MOD1BNT2 | | | | | | | | | |
| Studied schedule | 3087 | 2756 (89.3%) | 6 (0.2%) | 325 (10.5%) | 88 (2.9%) | 117 (3.8%) | 19 (0.6%) | 359 (11.6%) | 22 (0.7%) |
| Comparison schedule | 3084 | 2766 (89.7%) | 17 (0.6%) | 301 (9.8%) | 74 (2.4%) | 95 (3.1%) | 30 (1.0%) | 338 (11.0%) | 14 (0.5%) |
| BNT1MOD2BNT3 vs BNT1MOD2 | | | | | | | | | |
| Studied schedule | 8565 | 7762 (90.6%) | 19 (0.2%) | 784 (9.2%) | 124 (1.4%) | 132 (1.5%) | 19 (0.2%) | 339 (4.0%) | 13 (0.2%) |
| Comparison schedule | 8562 | 7771 (90.8%) | 27 (0.3%) | 764 (8.9%) | 105 (1.2%) | 140 (1.6%) | 23 (0.3%) | 401 (4.7%) | 26 (0.3%) |
| MOD1BNT2MOD3 vs MOD1BNT2 | | | | | | | | | |

| | Total individuals | Priority groups | | | Comorbidities | | | | |
|---|---|---|---|---|---|---|---|---|---|
| | | Others | Vulnerable | Healthcare workers | AIC | Cancer | CPD | CVD/DM | CKD |
| Studied schedule | 1755 | *(85.7%) | <5 | *(14.1%) | 40 (2.3%) | 56 (3.2%) | 15 (0.9%) | 174 (9.9%) | 5 (0.3%) |
| Comparison schedule | 1755 | 1560 (88.9%) | 10 (0.6%) | 185 (10.5%) | 42 (2.4%) | 50 (2.8%) | 13 (0.7%) | 162 (9.2%) | 6 (0.3%) |
| BNT1BNT2BNT3 vs BNT1BNT2 | | | | | | | | | |
| Studied schedule | 680361 | 593135 (87.2%) | 3444 (0.5%) | 83782 (12.3%) | 20891 (3.1%) | 32427 (4.8%) | 4837 (0.7%) | 86071 (12.7%) | 3968 (0.6%) |
| Comparison schedule | 679394 | 608706 (89.6%) | 4659 (0.7%) | 66029 (9.7%) | 18158 (2.7%) | 29366 (4.3%) | 6021 (0.9%) | 88308 (13.0%) | 4223 (0.6%) |
| MOD1MOD2MOD3 vs MOD1MOD2 | | | | | | | | | |
| Studied schedule | 81314 | 71669 (88.1%) | 449 (0.6%) | 9196 (11.3%) | 2432 (3.0%) | 4168 (5.1%) | 882 (1.1%) | 11755 (14.5%) | 691 (0.8%) |
| Comparison schedule | 81233 | 73630 (90.6%) | 522 (0.6%) | 7081 (8.7%) | 2226 (2.7%) | 3782 (4.7%) | 958 (1.2%) | 12113 (14.9%) | 699 (0.9%) |
| **Heterologous- vs homologous booster schedules** | | | | | | | | | |
| AZD1AZD2BNT3 vs BNT1BNT2BNT3 | | | | | | | | | |
| Studied schedule | 113620 | 102607 (90.3%) | 151 (0.1%) | 10862 (9.6%) | 4787 (4.2%) | 11602 (10.2%) | 3133 (2.8%) | 39538 (34.8%) | 1387 (1.2%) |
| Comparison schedule | 343376 | 307028 (89.4%) | 3487 (1.0%) | 32861 (9.6%) | 13946 (4.1%) | 40122 (11.7%) | 7845 (2.3%) | 103044 (30.0%) | 3940 (1.1%) |
| AZD1AZD2MOD3 vs MOD1MOD2MOD3 | | | | | | | | | |
| Studied schedule | 47168 | 43045 (91.3%) | 40 (0.1%) | 4083 (8.7%) | 1619 (3.4%) | 4139 (8.8%) | 852 (1.8%) | 12341 (26.2%) | 418 (0.9%) |
| Comparison schedule | 37383 | 34236 (91.6%) | 339 (0.9%) | 2808 (7.5%) | 1631 (4.4%) | 4197 (11.2%) | 1168 (3.1%) | 11741 (31.4%) | 594 (1.6%) |
| AZD1BNT2BNT3 vs BNT1BNT2BNT3 | | | | | | | | | |
| Studied schedule | 101326 | 82376 (81.3%) | 146 (0.1%) | 18804 (18.6%) | 5710 (5.6%) | 9008 (8.9%) | 2871 (2.8%) | 45343 (44.7%) | 1962 (1.9%) |
| Comparison schedule | 800650 | 670840 (83.8%) | 2393 (0.3%) | 127417 (15.9%) | 26970 (3.4%) | 35205 (4.4%) | 5225 (0.7%) | 94679 (11.8%) | 3536 (0.4%) |
| AZD1MOD2MOD3 vs MOD1MOD2MOD3 | | | | | | | | | |
| Studied schedule | 18738 | 15232 (81.3%) | 20 (0.1%) | 3486 (18.6%) | 1143 (6.1%) | 1548 (8.3%) | 494 (2.6%) | 8054 (43.0%) | 313 (1.7%) |
| Comparison schedule | 104355 | 92089 (88.2%) | 280 (0.3%) | 11986 (11.5%) | 3221 (3.1%) | 4717 (4.5%) | 1033 (1.0%) | 13539 (13.0%) | 637 (0.6%) |
| BNT1BNT2MOD3 vs BNT1BNT2BNT3 | | | | | | | | | |
| Studied schedule | 443176 | 395471 (89.2%) | 618 (0.1%) | 47087 (10.6%) | 11396 (2.6%) | 18402 (4.2%) | 3050 (0.7%) | 52572 (11.9%) | 1979 (0.4%) |
| Comparison schedule | 1203555 | 1048679 (87.1%) | 3998 (0.3%) | 150878 (12.5%) | 38914 (3.2%) | 73366 (6.1%) | 12683 (1.1%) | 194803 (16.2%) | 7774 (0.6%) |
| MOD1MOD2BNT3 vs BNT1BNT2BNT3 | | | | | | | | | |
| Studied schedule | 61418 | 54458 (88.7%) | 322 (0.5%) | 6638 (10.8%) | 1845 (3.0%) | 3241 (5.3%) | 668 (1.1%) | 9940 (16.2%) | 498 (0.8%) |
| Comparison schedule | 1367687 | 1193629 (87.3%) | 9553 (0.7%) | 164505 (12.0%) | 44595 (3.3%) | 90524 (6.6%) | 15764 (1.2%) | 242958 (17.8%) | 10720 (0.8%) |
| BNT1MOD2MOD3 vs BNT1BNT2BNT3 | | | | | | | | | |
| Studied schedule | 10277 | 9121 (88.8%) | 14 (0.1%) | 1142 (11.1%) | 144 (1.4%) | 216 (2.1%) | 33 (0.3%) | 478 (4.7%) | 25 (0.2%) |

| | Total individuals | Priority groups | | | Comorbidities | | | | |
|---|---|---|---|---|---|---|---|---|---|
| | | Others | Vulnerable | Healthcare workers | AIC | Cancer | CPD | CVD/DM | CKD |
| Comparison schedule | 940130 | 832230 (88.5%) | 941 (0.1%) | 106959 (11.4%) | 25112 (2.7%) | 37256 (4.0%) | 5717 (0.6%) | 100432 (10.7%) | 3775 (0.4%) |
| MOD1BNT2BNT3 vs BNT1BNT2BNT3 | | | | | | | | | |
| Studied schedule | 4504 | 3980 (88.4%) | 11 (0.2%) | 513 (11.4%) | 161 (3.6%) | 211 (4.7%) | 44 (1.0%) | 677 (15.0%) | 40 (0.9%) |
| Comparison schedule | 1324944 | 1159472 (87.5%) | 5926 (0.4%) | 159546 (12.0%) | 42163 (3.2%) | 81598 (6.2%) | 14193 (1.1%) | 218278 (16.5%) | 9155 (0.7%) |
| BNT1MOD2BNT3 vs BNT1BNT2BNT3 | | | | | | | | | |
| Studied schedule | 10148 | 9165 (90.3%) | 9 (0.1%) | 974 (9.6%) | 135 (1.3%) | 152 (1.5%) | 17 (0.2%) | 383 (3.8%) | 14 (0.1%) |
| Comparison schedule | 1018958 | 895450 (87.9%) | 1055 (0.1%) | 122453 (12.0%) | 28454 (2.8%) | 41659 (4.1%) | 6297 (0.6%) | 113197 (11.1%) | 4118 (0.4%) |
| MOD1BNT2MOD3 vs BNT1BNT2BNT3 | | | | | | | | | |
| Studied schedule | 2102 | 1792 (85.3%) | 6 (0.3%) | 304 (14.5%) | 48 (2.3%) | 73 (3.5%) | 18 (0.9%) | 218 (10.4%) | 7 (0.3%) |
| Comparison schedule | 961231 | 851745 (88.6%) | 1151 (0.1%) | 108335 (11.3%) | 26100 (2.7%) | 40695 (4.2%) | 6490 (0.7%) | 109474 (11.4%) | 4218 (0.4%) |
| MOD1MOD2MOD3 vs BNT1BNT2BNT3 | | | | | | | | | |
| Studied schedule | 150242 | 133798 (89.1%) | 1116 (0.7%) | 15328 (10.2%) | 4959 (3.3%) | 9629 (6.4%) | 2274 (1.5%) | 28345 (18.9%) | 1681 (1.1%) |
| Comparison schedule | 1312426 | 1142733 (87.1%) | 10313 (0.8%) | 159380 (12.1%) | 43691 (3.3%) | 91066 (6.9%) | 15804 (1.2%) | 244223 (18.6%) | 10855 (0.8%) |

AID denotes autoimmunity related conditions, CPD chronic pulmonary disease, CVD/DM cardiovascular conditions and diabetes, CKD moderate to severe renal disease (chronic kidney disease), and * that actual counts could not be presented (only percentage) due to national regulations on data privacy protection.

### Supplementary Table 9. Distribution of vaccine priority groups and comorbidities by comparison in Norway.

| | Total individuals | Priority groups | | | Comorbidities | | | | |
|---|---|---|---|---|---|---|---|---|---|
| | | Others | Healthcare | Vulnerable | AIC | Cancer | CPD | CVD/DM | CKD |
| **Heterologous booster- vs primary schedules** | | | | | | | | | |
| AZD1AZD2BNT3 vs AZD1AZD2 | | | | | | | | | |
| Studied schedule | 565 | 477 (84.4%) | 88 (15.6%) | 0 | <5 | 0 | 7 (1.2%) | 18 (3.2%) | 0 |
| Comparison schedule | 564 | 475 (84.2%) | 89 (15.8%) | 0 | <5 | 0 | <5 | 29 (5.1%) | 0 |
| AZD1AZD2MOD3 vs AZD1AZD2 | | | | | | | | | |
| Studied schedule | 214 | 196 (91.6%) | 18 (8.4%) | 0 | 0 | 0 | <5 | 8 (3.7%) | 0 |
| Comparison schedule | 213 | 204 (95.8%) | 9 (4.2%) | 0 | <5 | 0 | <5 | 16 (7.5%) | 0 |
| AZD1BNT2BNT3 vs AZD1BNT2 | | | | | | | | | |
| Studied schedule | 21069 | 2768 (13.1%) | 18297 (86.8%) | <5 | 337 (1.6%) | 88 (0.4%) | 1071 (5.1%) | 953 (4.5%) | 12 (0.1%) |
| Comparison schedule | 21032 | *(18.6%) | *(81.3%) | <5 | 280 (1.3%) | 65 (0.3%) | 1078 (5.1%) | 1088 (5.2%) | 7 (0.0%) |
| AZD1MOD2MOD3 vs AZD1MOD2 | | | | | | | | | |
| Studied schedule | 438 | 243 (55.5%) | 195 (44.5%) | 0 | 5 (1.1%) | 5 (1.1%) | 31 (7.1%) | 89 (20.3%) | 0 |
| Comparison schedule | 438 | 240 (54.8%) | 198 (45.2%) | 0 | 12 (2.7%) | 0 | 42 (9.6%) | 104 (23.7%) | <5 |
| BNT1BNT2MOD3 vs BNT1BNT2 | | | | | | | | | |
| Studied schedule | 238921 | 221311 (92.6%) | 17437 (7.3%) | 173 (0.1%) | 4736 (2.0%) | 3062 (1.3%) | 15655 (6.6%) | 24464 (10.2%) | 485 (0.2%) |
| Comparison schedule | 237807 | 218241 (91.8%) | 19412 (8.2%) | 154 (0.1%) | 5469 (2.3%) | 2709 (1.1%) | 15510 (6.5%) | 23759 (10.0%) | 639 (0.3%) |
| MOD1MOD2BNT3 vs MOD1MOD2 | | | | | | | | | |
| Studied schedule | 64393 | 57603 (89.5%) | 6757 (10.5%) | 33 (0.1%) | 784 (1.2%) | 406 (0.6%) | 3101 (4.8%) | 4009 (6.2%) | 73 (0.1%) |
| Comparison schedule | 63940 | 57583 (90.1%) | 6310 (9.9%) | 47 (0.1%) | 776 (1.2%) | 397 (0.6%) | 3058 (4.8%) | 4276 (6.7%) | 69 (0.1%) |
| BNT1MOD2MOD3 vs BNT1MOD2 | | | | | | | | | |
| Studied schedule | 71546 | *(96.4%) | *(3.6%) | <5 | 486 (0.7%) | 155 (0.2%) | 1795 (2.5%) | 1260 (1.8%) | 19 (0.0%) |
| Comparison schedule | 71035 | *(97.1%) | *(2.9%) | <5 | 538 (0.8%) | 191 (0.3%) | 1684 (2.4%) | 1727 (2.4%) | 24 (0.0%) |
| MOD1BNT2BNT3 vs MOD1BNT2 | | | | | | | | | |
| Studied schedule | 7859 | 7373 (93.8%) | 486 (6.2%) | 0 | 42 (0.5%) | 16 (0.2%) | 187 (2.4%) | 81 (1.0%) | <5 |
| Comparison schedule | 7793 | *(94.8%) | *(5.2%) | <5 | 44 (0.6%) | 12 (0.2%) | 192 (2.5%) | 89 (1.1%) | 0 |
| BNT1MOD2BNT3 vs BNT1MOD2 | | | | | | | | | |
| Studied schedule | 141657 | 133621 (94.3%) | 8012 (5.7%) | 24 (0.0%) | 914 (0.6%) | 231 (0.2%) | 3898 (2.8%) | 1938 (1.4%) | 18 (0.0%) |
| Comparison schedule | 140531 | 132121 (94.0%) | 8377 (6.0%) | 33 (0.0%) | 859 (0.6%) | 211 (0.2%) | 3819 (2.7%) | 1988 (1.4%) | 21 (0.0%) |
| MOD1BNT2MOD3 vs MOD1BNT2 | | | | | | | | | |

| | Total individuals | Priority groups | | | Comorbidities | | | | |
|---|---|---|---|---|---|---|---|---|---|
| | | **Others** | **Healthcare** | **Vulnerable** | **AIC** | **Cancer** | **CPD** | **CVD/DM** | **CKD** |
| Studied schedule | 4249 | 4082 (96.1%) | 167 (3.9%) | 0 | 24 (0.6%) | <5 | 91 (2.1%) | 70 (1.6%) | 0 |
| Comparison schedule | 4205 | 4049 (96.3%) | 156 (3.7%) | 0 | 30 (0.7%) | <5 | 73 (1.7%) | 88 (2.1%) | 0 |
| BNT1BNT2BNT3 vs BNT1BNT2 | | | | | | | | | |
| Studied schedule | 482290 | 425962 (88.3%) | 54111 (11.2%) | 2217 (0.5%) | 11882 (2.5%) | 5951 (1.2%) | 32402 (6.7%) | 49294 (10.2%) | 1308 (0.3%) |
| Comparison schedule | 478944 | 429417 (89.7%) | 47141 (9.8%) | 2386 (0.5%) | 10078 (2.1%) | 5630 (1.2%) | 32642 (6.8%) | 50975 (10.6%) | 1396 (0.3%) |
| MOD1MOD2MOD3 vs MOD1MOD2 | | | | | | | | | |
| Studied schedule | 63974 | 59805 (93.5%) | 4144 (6.5%) | 25 (0.0%) | 951 (1.5%) | 560 (0.9%) | 3506 (5.5%) | 4700 (7.3%) | 80 (0.1%) |
| Comparison schedule | 63591 | 59597 (93.7%) | 3966 (6.2%) | 28 (0.0%) | 905 (1.4%) | 453 (0.7%) | 3210 (5.0%) | 5105 (8.0%) | 91 (0.1%) |
| **Heterologous- vs homologous booster schedules** | | | | | | | | | |
| AZD1AZD2BNT3 vs BNT1BNT2BNT3 | | | | | | | | | |
| Studied schedule | 740 | NE | NE | NE | <5 | <5 | 14 (1.9%) | 31 (4.2%) | 0 |
| Comparison schedule | 1309526 | 1161481 (88.7%) | 145965 (11.1%) | 2080 (0.2%) | 33741 (2.6%) | 28686 (2.2%) | 106881 (8.2%) | 204775 (15.6%) | 5120 (0.4%) |
| AZD1AZD2MOD3 vs MOD1MOD2MOD3 | | | | | | | | | |
| Studied schedule | 246 | NE | NE | NE | 0 | <5 | 6 (2.4%) | 13 (5.3%) | 0 |
| Comparison schedule | 144180 | 136872 (94.9%) | 7271 (5.0%) | 37 (0.0%) | 2485 (1.7%) | 1831 (1.3%) | 8372 (5.8%) | 14148 (9.8%) | 267 (0.2%) |
| AZD1BNT2BNT3 vs BNT1BNT2BNT3 | | | | | | | | | |
| Studied schedule | 96725 | 19943 (20.6%) | 76775 (79.4%) | 7 (0.0%) | 1618 (1.7%) | 656 (0.7%) | 5819 (6.0%) | 7484 (7.7%) | 67 (0.1%) |
| Comparison schedule | 752266 | 638535 (84.9%) | 113395 (15.1%) | 336 (0.0%) | 19541 (2.6%) | 11234 (1.5%) | 54225 (7.2%) | 84993 (11.3%) | 1802 (0.2%) |
| AZD1MOD2MOD3 vs MOD1MOD2MOD3 | | | | | | | | | |
| Studied schedule | 1077 | 637 (59.1%) | 440 (40.9%) | 0 | 32 (3.0%) | 17 (1.6%) | 135 (12.5%) | 238 (22.1%) | <5 |
| Comparison schedule | 114926 | 108578 (94.5%) | 6328 (5.5%) | 20 (0.0%) | 1767 (1.5%) | 839 (0.7%) | 5505 (4.8%) | 7312 (6.4%) | 118 (0.1%) |
| BNT1BNT2MOD3 vs BNT1BNT2BNT3 | | | | | | | | | |
| Studied schedule | 320119 | 298405 (93.2%) | 21589 (6.7%) | 125 (0.0%) | 6203 (1.9%) | 4311 (1.3%) | 20009 (6.3%) | 32280 (10.1%) | 687 (0.2%) |
| Comparison schedule | 1248035 | 1126321 (90.2%) | 118616 (9.5%) | 3098 (0.2%) | 34964 (2.8%) | 32543 (2.6%) | 109061 (8.7%) | 226562 (18.2%) | 6034 (0.5%) |
| MOD1MOD2BNT3 vs BNT1BNT2BNT3 | | | | | | | | | |
| Studied schedule | 120544 | 100174 (83.1%) | 20243 (16.8%) | 127 (0.1%) | 1867 (1.5%) | 1812 (1.5%) | 7389 (6.1%) | 12926 (10.7%) | 299 (0.2%) |
| Comparison schedule | 1497941 | 1343351 (89.7%) | 147942 (9.9%) | 6648 (0.4%) | 41267 (2.8%) | 39873 (2.7%) | 131267 (8.8%) | 280210 (18.7%) | 8446 (0.6%) |
| BNT1MOD2MOD3 vs BNT1BNT2BNT3 | | | | | | | | | |
| Studied schedule | 97079 | 94416 (97.3%) | 2658 (2.7%) | 5 (0.0%) | 670 (0.7%) | 190 (0.2%) | 2209 (2.3%) | 1528 (1.6%) | 23 (0.0%) |
| Comparison schedule | 488451 | 457202 (93.6%) | 31126 (6.4%) | 123 (0.0%) | 8319 (1.7%) | 3410 (0.7%) | 24717 (5.1%) | 34429 (7.0%) | 551 (0.1%) |

| | Total individuals | Priority groups | | | Comorbidities | | | | |
|---|---|---|---|---|---|---|---|---|---|
| | | Others | Healthcare | Vulnerable | AIC | Cancer | CPD | CVD/DM | CKD |
| MOD1BNT2BNT3 vs BNT1BNT2BNT3 | | | | | | | | | |
| Studied schedule | 12620 | 11859 (94.0%) | 761 (6.0%) | 0 | 61 (0.5%) | 18 (0.1%) | 320 (2.5%) | 92 (0.7%) | <5 |
| Comparison schedule | 588157 | 537528 (91.4%) | 50512 (8.6%) | 117 (0.0%) | 9302 (1.6%) | 3421 (0.6%) | 30027 (5.1%) | 34902 (5.9%) | 570 (0.1%) |
| BNT1MOD2BNT3 vs BNT1BNT2BNT3 | | | | | | | | | |
| Studied schedule | 194343 | 183680 (94.5%) | 10647 (5.5%) | 16 (0.0%) | 1088 (0.6%) | 252 (0.1%) | 5144 (2.6%) | 2104 (1.1%) | 20 (0.0%) |
| Comparison schedule | 513894 | 471166 (91.7%) | 42655 (8.3%) | 73 (0.0%) | 6900 (1.3%) | 2271 (0.4%) | 23399 (4.6%) | 22798 (4.4%) | 357 (0.1%) |
| MOD1BNT2MOD3 vs BNT1BNT2BNT3 | | | | | | | | | |
| Studied schedule | 7458 | 7283 (97.7%) | 175 (2.3%) | 0 | 41 (0.5%) | 9 (0.1%) | 144 (1.9%) | 98 (1.3%) | 0 |
| Comparison schedule | 532352 | 494435 (92.9%) | 37788 (7.1%) | 129 (0.0%) | 8490 (1.6%) | 3395 (0.6%) | 25977 (4.9%) | 34277 (6.4%) | 529 (0.1%) |
| MOD1MOD2MOD3 vs BNT1BNT2BNT3 | | | | | | | | | |
| Studied schedule | 141956 | 135174 (95.2%) | 6741 (4.7%) | 41 (0.0%) | 2240 (1.6%) | 1774 (1.2%) | 8235 (5.8%) | 14241 (10.0%) | 260 (0.2%) |
| Comparison schedule | 1218904 | 1099384 (90.2%) | 117603 (9.6%) | 1917 (0.2%) | 31423 (2.6%) | 28737 (2.4%) | 103097 (8.5%) | 210272 (17.3%) | 5235 (0.4%) |

AID denotes autoimmunity related conditions, CPD chronic pulmonary disease, CVD/DM cardiovascular conditions and diabetes, CKD moderate to severe renal disease (chronic kidney disease), NE not estimable due national regulations on data privacy protection, and * that actual counts could not be presented (only percentage) due to national regulations on data privacy protection.

**Supplementary Table 10. Distribution of vaccine priority groups and comorbidities by comparison in Sweden.**

| | Total individuals | Priority groups | | | Comorbidities | | | | |
|---|---|---|---|---|---|---|---|---|---|
| | | Others | Healthcare personnel | Vulnerable | AIC | Cancer | CPD | CVD/DM | CKD |
| **Heterologous booster- vs primary schedules** | | | | | | | | | |
| AZD1AZD2BNT3 vs AZD1AZD2 | | | | | | | | | |
| Studied schedule | 63120 | 50703 (80.3%) | 12397 (19.6%) | 20 (0.0%) | 3363 (5.3%) | 7746 (12.3%) | 2817 (4.5%) | 13667 (21.7%) | 962 (1.5%) |
| Comparison schedule | 62243 | 51504 (82.7%) | 10711 (17.2%) | 28 (0.0%) | 3049 (4.9%) | 6167 (9.9%) | 3258 (5.2%) | 14641 (23.5%) | 1347 (2.2%) |
| AZD1AZD2MOD3 vs AZD1AZD2 | | | | | | | | | |
| Studied schedule | 50634 | 42726 (84.4%) | 7898 (15.6%) | 10 (0.0%) | 2715 (5.4%) | 6157 (12.2%) | 2259 (4.5%) | 11661 (23.0%) | 886 (1.7%) |
| Comparison schedule | 50169 | 42801 (85.3%) | 7345 (14.6%) | 23 (0.0%) | 2594 (5.2%) | 5368 (10.7%) | 2727 (5.4%) | 12747 (25.4%) | 1166 (2.3%) |
| AZD1BNT2BNT3 vs AZD1BNT2 | | | | | | | | | |
| Studied schedule | 26365 | 12064 (45.8%) | 14243 (54.0%) | 58 (0.2%) | 996 (3.8%) | 888 (3.4%) | 815 (3.1%) | 1823 (6.9%) | 159 (0.6%) |
| Comparison schedule | 26083 | 12324 (47.2%) | 13697 (52.5%) | 62 (0.2%) | 871 (3.3%) | 766 (2.9%) | 688 (2.6%) | 1663 (6.4%) | 156 (0.6%) |
| AZD1MOD2MOD3 vs AZD1MOD2 | | | | | | | | | |
| Studied schedule | 2450 | 1129 (46.1%) | 1321 (53.9%) | 0 | 78 (3.2%) | 80 (3.3%) | 81 (3.3%) | 178 (7.3%) | 21 (0.9%) |
| Comparison schedule | 2428 | 1149 (47.3%) | 1279 (52.7%) | 0 | 100 (4.1%) | 77 (3.2%) | 65 (2.7%) | 189 (7.8%) | 25 (1.0%) |
| BNT1BNT2MOD3 vs BNT1BNT2 | | | | | | | | | |
| Studied schedule | 504359 | 476510 (94.5%) | 26625 (5.3%) | 1224 (0.2%) | 20200 (4.0%) | 24367 (4.8%) | 13623 (2.7%) | 51575 (10.2%) | 4534 (0.9%) |
| Comparison schedule | 501571 | 471388 (94.0%) | 28512 (5.7%) | 1671 (0.3%) | 19915 (4.0%) | 21372 (4.3%) | 14047 (2.8%) | 56089 (11.2%) | 5091 (1.0%) |
| MOD1MOD2BNT3 vs MOD1MOD2 | | | | | | | | | |
| Studied schedule | 105725 | 98567 (93.2%) | 7080 (6.7%) | 78 (0.1%) | 3960 (3.7%) | 4430 (4.2%) | 3425 (3.2%) | 9826 (9.3%) | 968 (0.9%) |
| Comparison schedule | 105134 | 97967 (93.2%) | 7079 (6.7%) | 88 (0.1%) | 3776 (3.6%) | 3956 (3.8%) | 3698 (3.5%) | 11593 (11.0%) | 1251 (1.2%) |
| BNT1MOD2MOD3 vs BNT1MOD2 | | | | | | | | | |
| Studied schedule | 609 | 595 (97.7%) | 14 (2.3%) | 0 | 20 (3.3%) | 11 (1.8%) | 12 (2.0%) | 31 (5.1%) | 6 (1.0%) |
| Comparison schedule | 608 | 590 (97.0%) | 18 (3.0%) | 0 | 16 (2.6%) | 15 (2.5%) | 8 (1.3%) | 34 (5.6%) | <5 |
| MOD1BNT2BNT3 vs MOD1BNT2 | | | | | | | | | |
| Studied schedule | 1586 | 1511 (95.3%) | 75 (4.7%) | 0 | 36 (2.3%) | 22 (1.4%) | 54 (3.4%) | 54 (3.4%) | 7 (0.4%) |
| Comparison schedule | 1583 | 1498 (94.6%) | 85 (5.4%) | 0 | 37 (2.3%) | 21 (1.3%) | 42 (2.7%) | 57 (3.6%) | 8 (0.5%) |
| BNT1MOD2BNT3 vs BNT1MOD2 | | | | | | | | | |
| Studied schedule | 2243 | *(97.1%) | *(2.8%) | <5 | 52 (2.3%) | 16 (0.7%) | 47 (2.1%) | 58 (2.6%) | 7 (0.3%) |
| Comparison schedule | 2240 | *(97.4%) | *(2.6%) | <5 | 42 (1.9%) | 22 (1.0%) | 38 (1.7%) | 52 (2.3%) | 7 (0.3%) |
| MOD1BNT2MOD3 vs MOD1BNT2 | | | | | | | | | |

| | Total individuals | Priority groups | | | Comorbidities | | | | |
|---|---|---|---|---|---|---|---|---|---|
| | | Others | Healthcare personnel | Vulnerable | AIC | Cancer | CPD | CVD/DM | CKD |
| Studied schedule | 327 | 306 (93.6%) | 21 (6.4%) | 0 | 11 (3.4%) | 13 (4.0%) | <5 | 22 (6.7%) | <5 |
| Comparison schedule | 327 | NE | NE | NE | 7 (2.1%) | 13 (4.0%) | 9 (2.8%) | 28 (8.6%) | <5 |
| BNT1BNT2BNT3 vs BNT1BNT2 | | | | | | | | | |
| Studied schedule | 791382 | 732823 (92.6%) | 56257 (7.1%) | 2302 (0.3%) | 33770 (4.3%) | 37846 (4.8%) | 25624 (3.2%) | 83790 (10.6%) | 7132 (0.9%) |
| Comparison schedule | 784699 | 725726 (92.5%) | 57205 (7.3%) | 1768 (0.2%) | 31997 (4.1%) | 33243 (4.2%) | 25589 (3.3%) | 88450 (11.3%) | 8153 (1.0%) |
| MOD1MOD2MOD3 vs MOD1MOD2 | | | | | | | | | |
| Studied schedule | 93241 | 87382 (93.7%) | 5825 (6.2%) | 34 (0.0%) | 3937 (4.2%) | 4180 (4.5%) | 2701 (2.9%) | 8997 (9.6%) | 882 (0.9%) |
| Comparison schedule | 92640 | 86774 (93.7%) | 5793 (6.3%) | 73 (0.1%) | 3879 (4.2%) | 4283 (4.6%) | 2907 (3.1%) | 12063 (13.0%) | 1171 (1.3%) |
| **Heterologous- vs homologous booster schedules** | | | | | | | | | |
| AZD1AZD2BNT3 vs BNT1BNT2BNT3 | | | | | | | | | |
| Studied schedule | 355948 | 317235 (89.1%) | 38643 (10.9%) | 70 (0.0%) | 20083 (5.6%) | 51177 (14.4%) | 17223 (4.8%) | 90726 (25.5%) | 6221 (1.7%) |
| Comparison schedule | 1209728 | 1085857 (89.8%) | 122555 (10.1%) | 1316 (0.1%) | 67799 (5.6%) | 123235 (10.2%) | 53861 (4.5%) | 249938 (20.7%) | 18270 (1.5%) |
| AZD1AZD2MOD3 vs MOD1MOD2MOD3 | | | | | | | | | |
| Studied schedule | 120874 | 106891 (88.4%) | 13958 (11.5%) | 25 (0.0%) | 6698 (5.5%) | 15571 (12.9%) | 5452 (4.5%) | 29475 (24.4%) | 2095 (1.7%) |
| Comparison schedule | 118024 | 109856 (93.1%) | 8114 (6.9%) | 54 (0.0%) | 6006 (5.1%) | 9893 (8.4%) | 4352 (3.7%) | 19539 (16.6%) | 1604 (1.4%) |
| AZD1BNT2BNT3 vs BNT1BNT2BNT3 | | | | | | | | | |
| Studied schedule | 57041 | 23519 (41.2%) | 33345 (58.5%) | 177 (0.3%) | 2262 (4.0%) | 2149 (3.8%) | 1591 (2.8%) | 4409 (7.7%) | 326 (0.6%) |
| Comparison schedule | 1318686 | 1175780 (89.2%) | 141662 (10.7%) | 1244 (0.1%) | 63957 (4.9%) | 83772 (6.4%) | 44839 (3.4%) | 184034 (14.0%) | 12443 (0.9%) |
| AZD1MOD2MOD3 vs MOD1MOD2MOD3 | | | | | | | | | |
| Studied schedule | 4656 | 1933 (41.5%) | 2718 (58.4%) | 5 (0.1%) | 188 (4.0%) | 174 (3.7%) | 126 (2.7%) | 342 (7.3%) | 31 (0.7%) |
| Comparison schedule | 128714 | 118932 (92.4%) | 9723 (7.6%) | 59 (0.0%) | 5762 (4.5%) | 6808 (5.3%) | 3674 (2.9%) | 14430 (11.2%) | 1168 (0.9%) |
| BNT1BNT2MOD3 vs BNT1BNT2BNT3 | | | | | | | | | |
| Studied schedule | 756029 | 714409 (94.5%) | 39787 (5.3%) | 1833 (0.2%) | 31288 (4.1%) | 42163 (5.6%) | 21918 (2.9%) | 88227 (11.7%) | 7320 (1.0%) |
| Comparison schedule | 1720151 | 1563848 (90.9%) | 149750 (8.7%) | 6553 (0.4%) | 91882 (5.3%) | 176206 (10.2%) | 77265 (4.5%) | 350251 (20.4%) | 29997 (1.7%) |
| MOD1MOD2BNT3 vs BNT1BNT2BNT3 | | | | | | | | | |
| Studied schedule | 183788 | 170057 (92.5%) | 13540 (7.4%) | 191 (0.1%) | 8371 (4.6%) | 15073 (8.2%) | 8016 (4.4%) | 30755 (16.7%) | 3210 (1.7%) |
| Comparison schedule | 1975964 | 1794308 (90.8%) | 170994 (8.7%) | 10662 (0.5%) | 99596 (5.0%) | 187791 (9.5%) | 88017 (4.5%) | 375929 (19.0%) | 33725 (1.7%) |
| BNT1MOD2MOD3 vs BNT1BNT2BNT3 | | | | | | | | | |
| Studied schedule | 806 | 782 (97.0%) | 24 (3.0%) | 0 | 28 (3.5%) | 19 (2.4%) | 16 (2.0%) | 58 (7.2%) | 5 (0.6%) |

| | Total individuals | Priority groups | | | Comorbidities | | | | |
|---|---|---|---|---|---|---|---|---|---|
| | | **Others** | **Healthcare personnel** | **Vulnerable** | **AIC** | **Cancer** | **CPD** | **CVD/DM** | **CKD** |
| Comparison schedule | 1627528 | 1472656 (90.5%) | 151714 (9.3%) | 3158 (0.2%) | 85631 (5.3%) | 149470 (9.2%) | 67835 (4.2%) | 300977 (18.5%) | 23574 (1.4%) |
| MOD1BNT2BNT3 vs BNT1BNT2BNT3 | | | | | | | | | |
| Studied schedule | 2238 | *(93.5%) | *(6.4%) | <5 | 70 (3.1%) | 76 (3.4%) | 86 (3.8%) | 148 (6.6%) | 19 (0.8%) |
| Comparison schedule | 1977303 | 1797743 (90.9%) | 171665 (8.7%) | 7895 (0.4%) | 98467 (5.0%) | 179804 (9.1%) | 86413 (4.4%) | 361254 (18.3%) | 31562 (1.6%) |
| BNT1MOD2BNT3 vs BNT1BNT2BNT3 | | | | | | | | | |
| Studied schedule | 2884 | *(96.6%) | *(3.3%) | <5 | 77 (2.7%) | 24 (0.8%) | 57 (2.0%) | 86 (3.0%) | 9 (0.3%) |
| Comparison schedule | 1267739 | 1155261 (91.1%) | 111398 (8.8%) | 1080 (0.1%) | 54226 (4.3%) | 58129 (4.6%) | 39072 (3.1%) | 138057 (10.9%) | 9619 (0.8%) |
| MOD1BNT2MOD3 vs BNT1BNT2BNT3 | | | | | | | | | |
| Studied schedule | 433 | 410 (94.7%) | 23 (5.3%) | 0 | 17 (3.9%) | 26 (6.0%) | 7 (1.6%) | 35 (8.1%) | <5 |
| Comparison schedule | 1709913 | 1551855 (90.8%) | 152355 (8.9%) | 5703 (0.3%) | 90923 (5.3%) | 168826 (9.9%) | 75099 (4.4%) | 336278 (19.7%) | 28060 (1.6%) |
| MOD1MOD2MOD3 vs BNT1BNT2BNT3 | | | | | | | | | |
| Studied schedule | 182263 | 171145 (93.9%) | 11020 (6.0%) | 98 (0.1%) | 8854 (4.9%) | 16402 (9.0%) | 7112 (3.9%) | 31027 (17.0%) | 2980 (1.6%) |
| Comparison schedule | 1782303 | 1615930 (90.7%) | 150588 (8.4%) | 15785 (0.9%) | 96523 (5.4%) | 191863 (10.8%) | 83252 (4.7%) | 377479 (21.2%) | 34389 (1.9%) |

AID denotes autoimmunity related conditions, CPD chronic pulmonary disease, CVD/DM cardiovascular conditions and diabetes, CKD moderate to severe renal disease (chronic kidney disease), NE not estimable due national regulations on data privacy protection, and * that actual counts could not be presented (only percentage) due to national regulations on data privacy protection.

**Supplementary Figure 1. Density plots of the distribution of age and index date for the matched analyses comparing heterologous booster schedules and primary schedules in Denmark.**

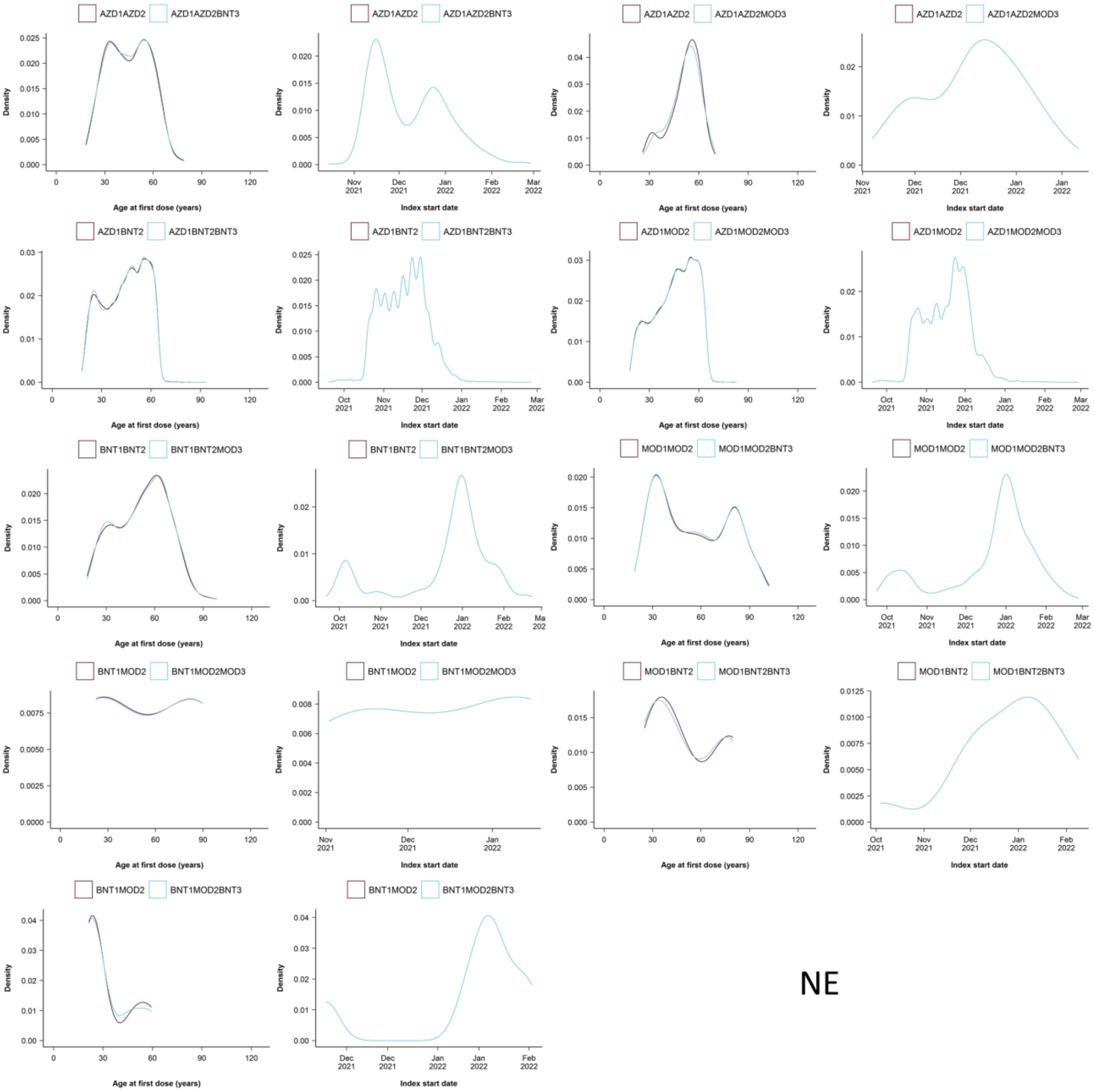

NE denotes not estimated (for MOD1BNT2MOD3 vs MOD1BNT2). Note that only those individuals who had follow-up time during the country-specific periods of omicron variant predominance were included in the final matched comparisons.

**Supplementary Figure 2. Density plots of the distribution of age and index date for the matched analyses comparing heterologous booster schedules and primary schedules in Finland.**

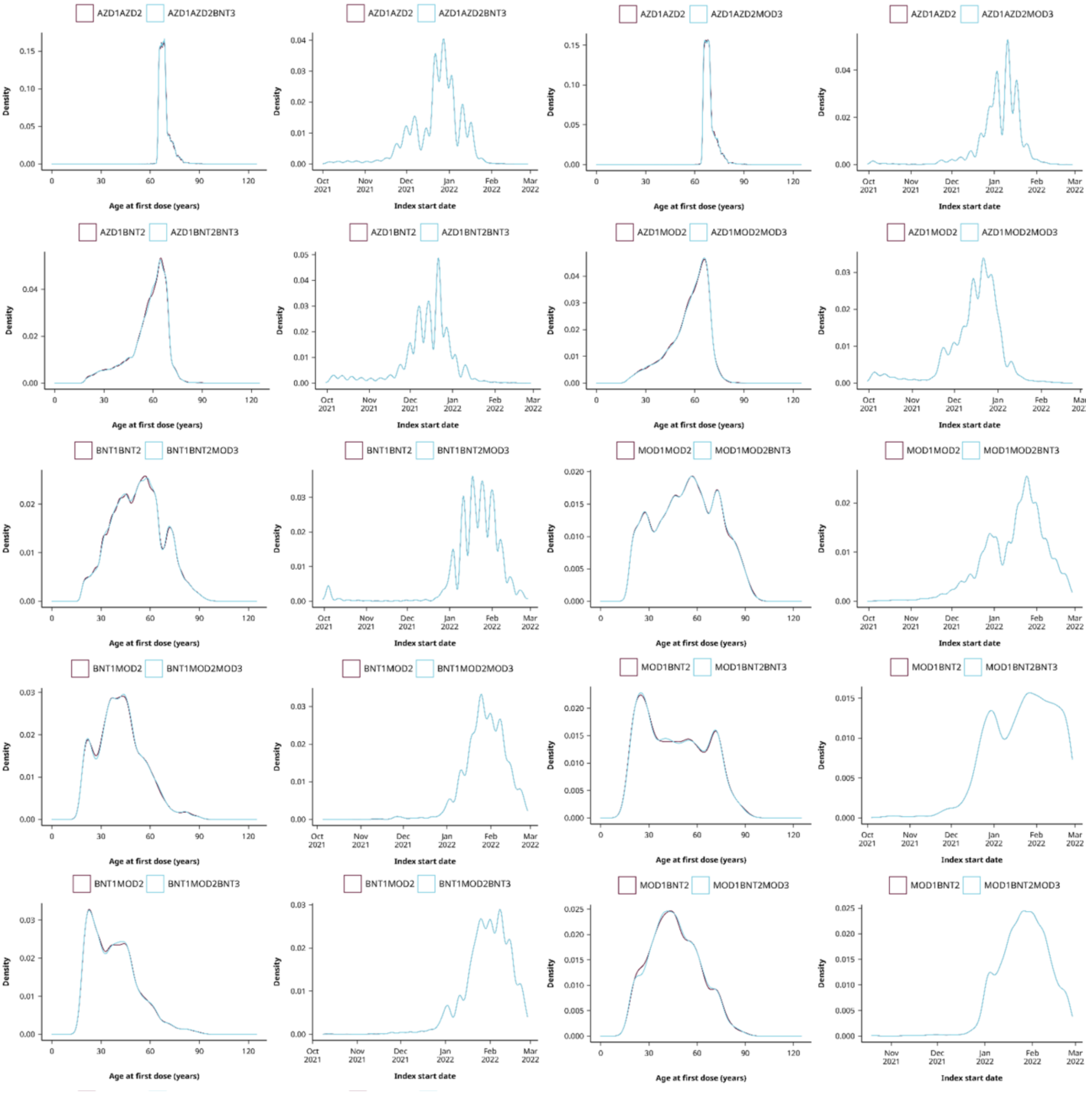

Note that only those individuals who had follow-up time during the country-specific periods of omicron variant predominance were included in the final matched comparisons.

**Supplementary Figure 3. Density plots of the distribution of age and index date for the matched analyses comparing heterologous booster schedules and primary schedules in Norway.**

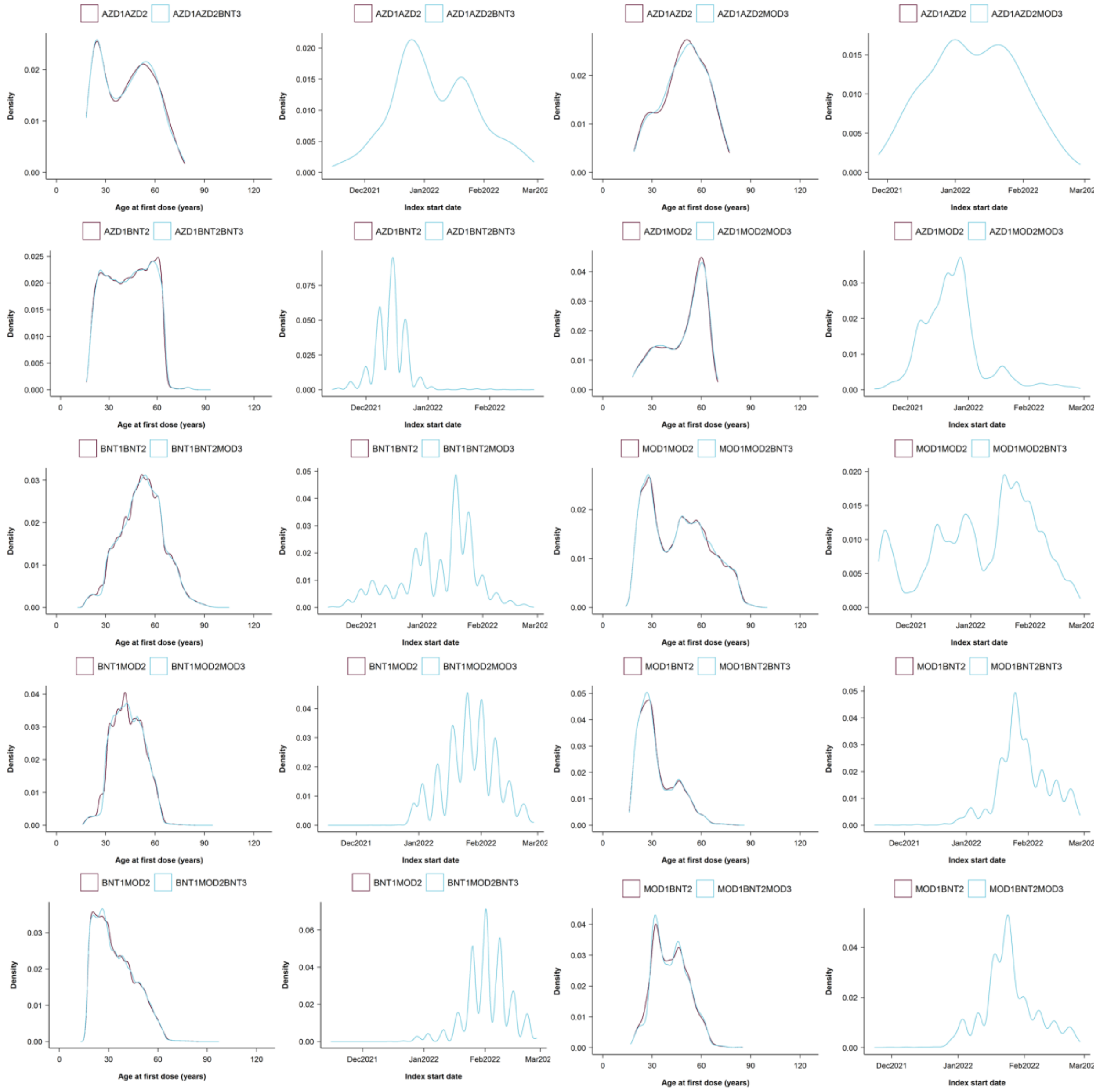

Note that only those individuals who had follow-up time during the country-specific periods of omicron variant predominance were included in the final matched comparisons.

**Supplementary Figure 4. Density plots of the distribution of age and index date for the matched analyses comparing heterologous booster schedules and primary schedules in Sweden.**

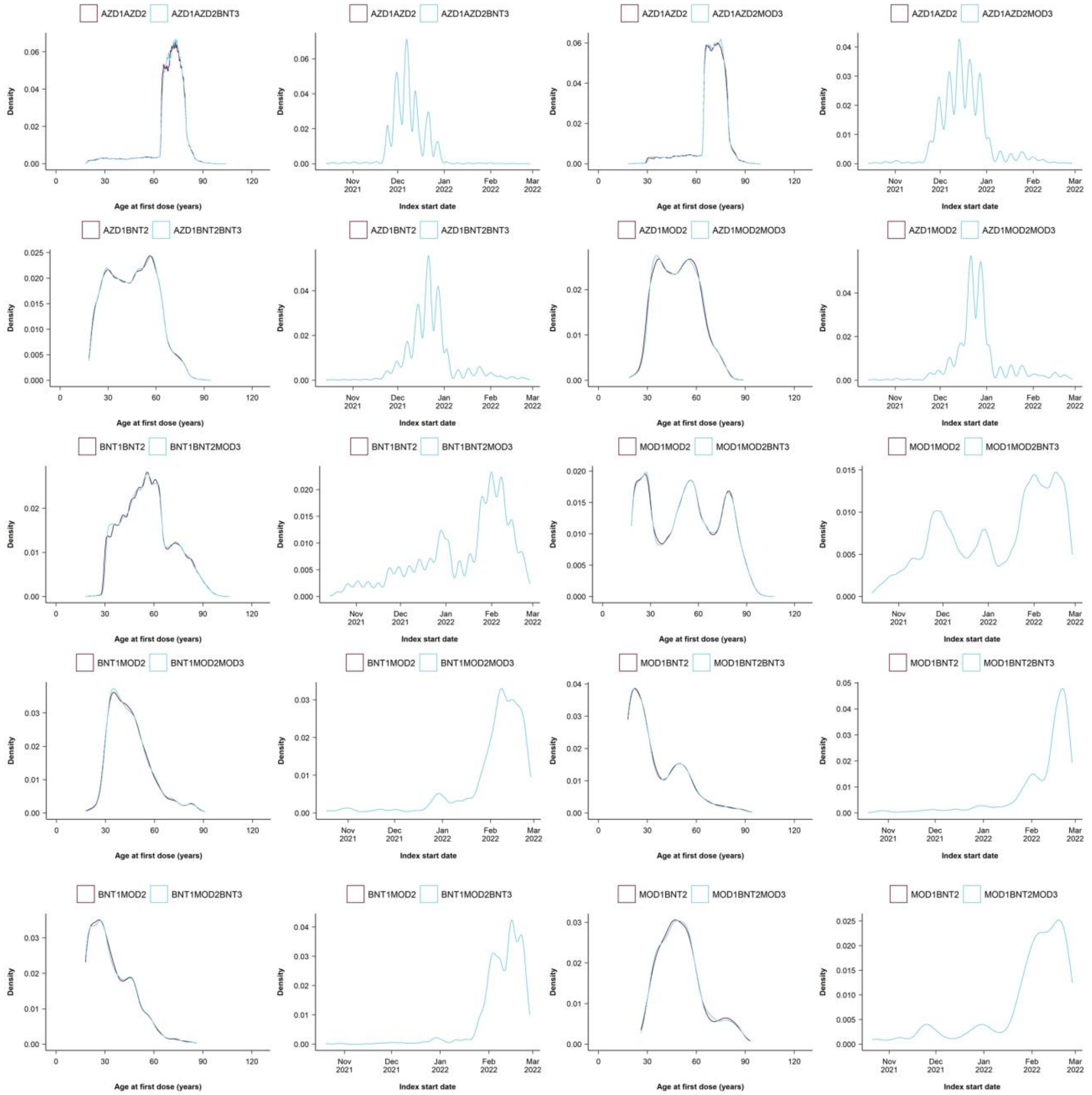

Note that only those individuals who had follow-up time during the country-specific periods of omicron variant predominance were included in the final matched comparisons.

**Supplementary Figure 5. Density plots of the distribution of age and index date for the weighted analyses comparing heterologous and homologous booster schedules in Denmark.**

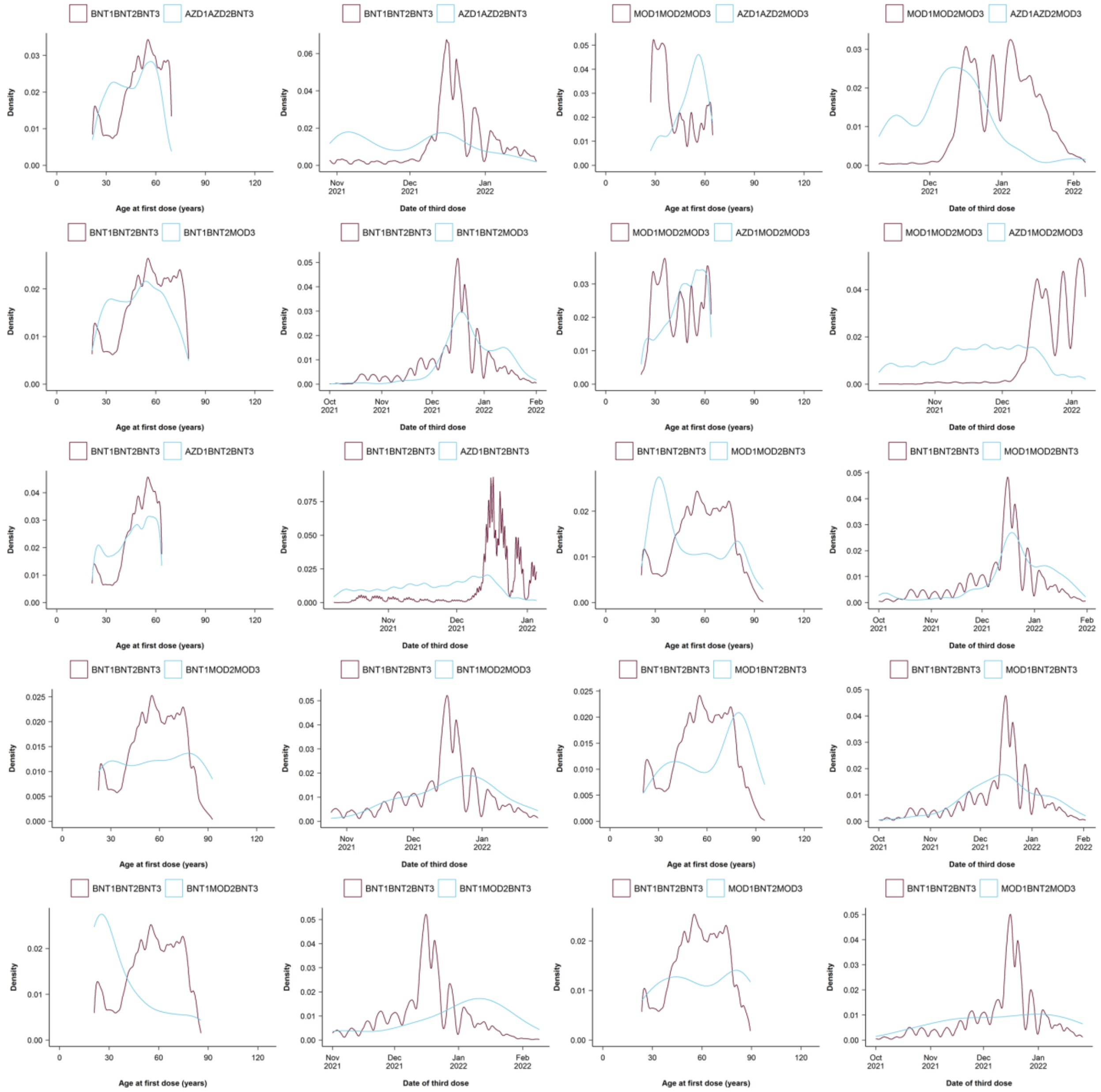

**Supplementary Figure 6. Density plots of the distribution of age and index date for the weighted analyses comparing heterologous and homologous booster schedules in Finland.**

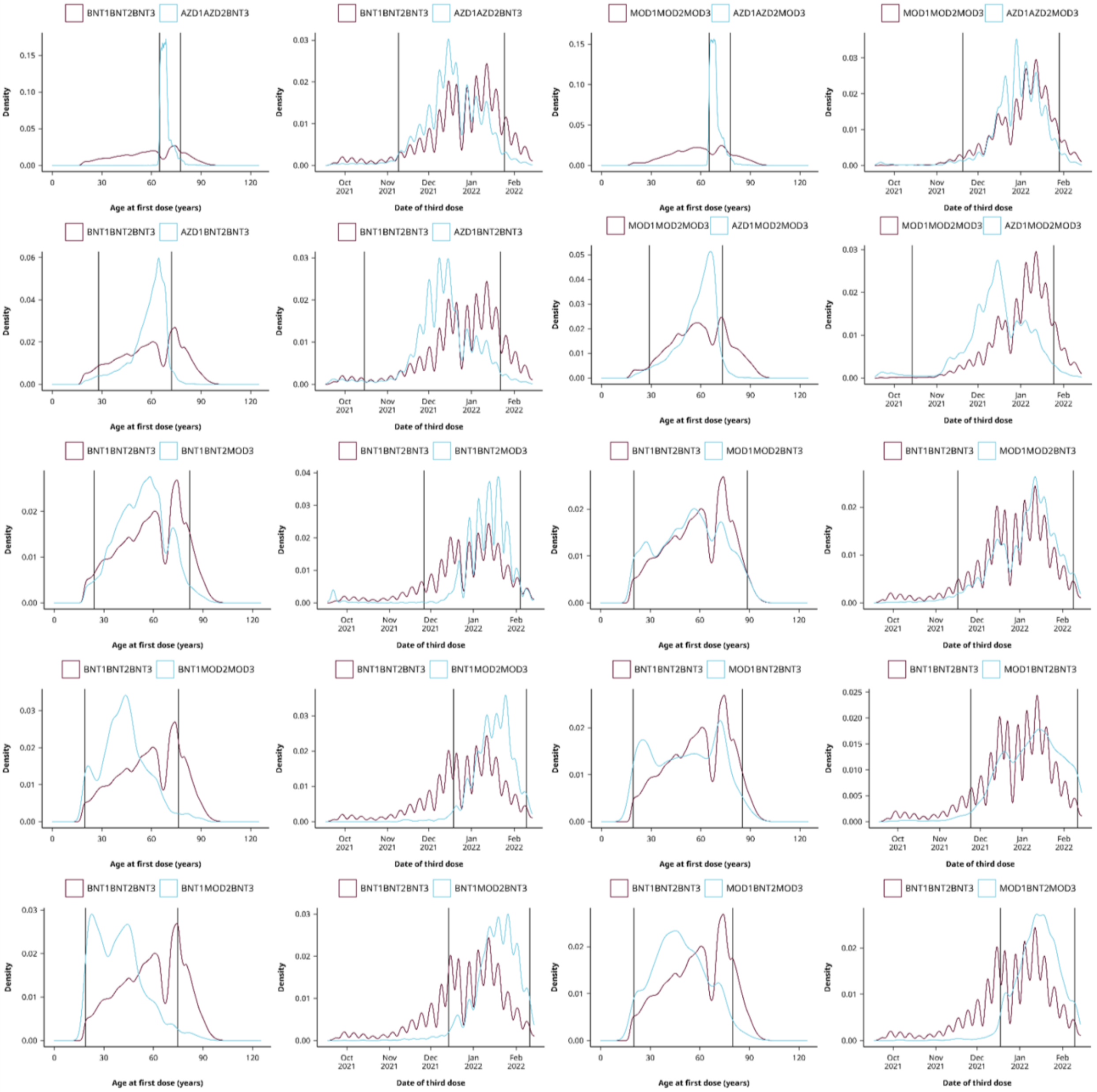

**Supplementary Figure 7. Density plots of the distribution of age and index date for the weighted analyses comparing heterologous and homologous booster schedules in Norway.**

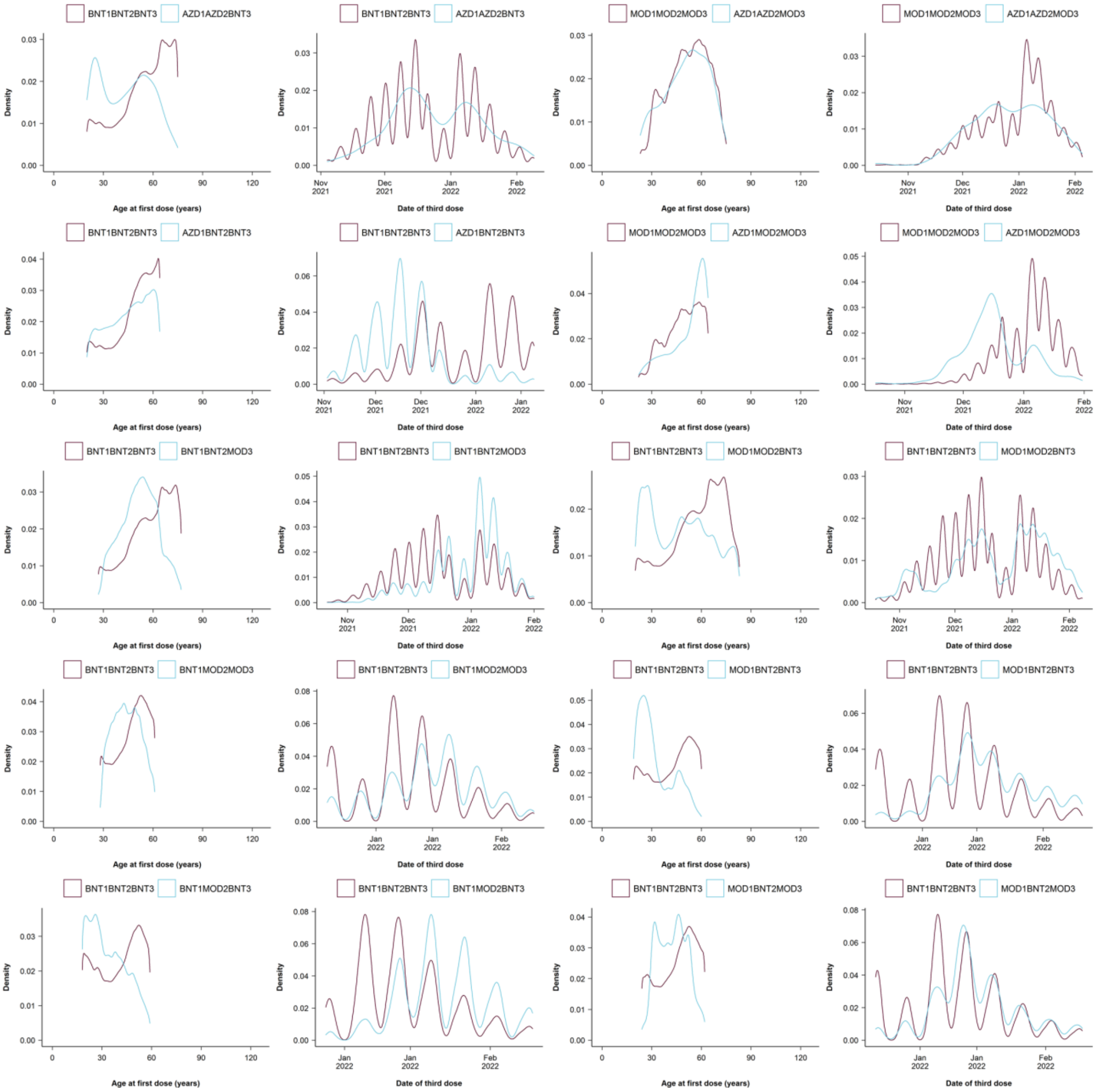

**Supplementary Figure 8. Density plots of the distribution of age and index date for the weighted analyses comparing heterologous and homologous booster schedules in Sweden.**

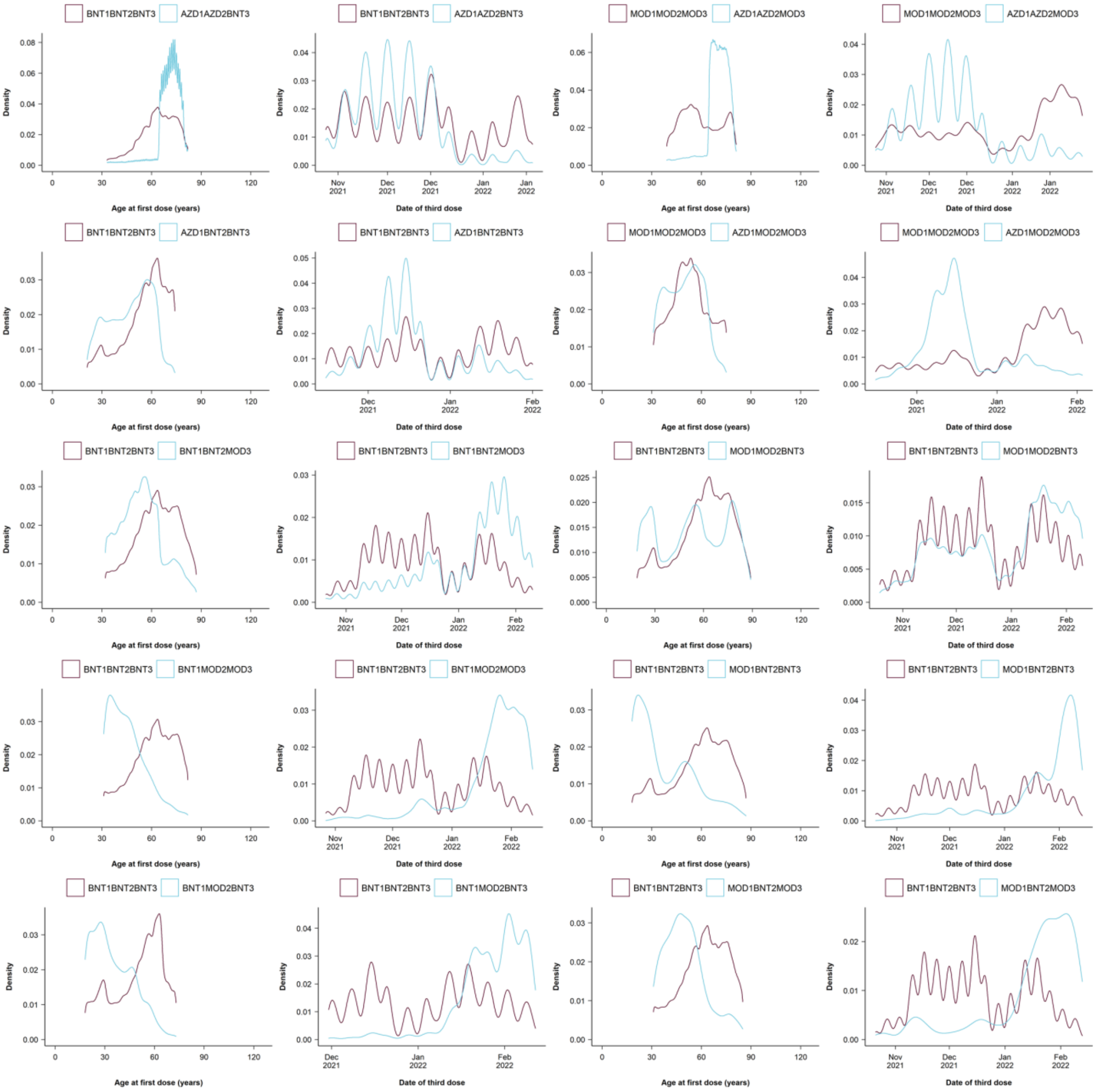

**Supplementary Figure 9. Density plots of the distribution of age and index date for the matched analyses comparing homologous booster schedules and primary schedules.**

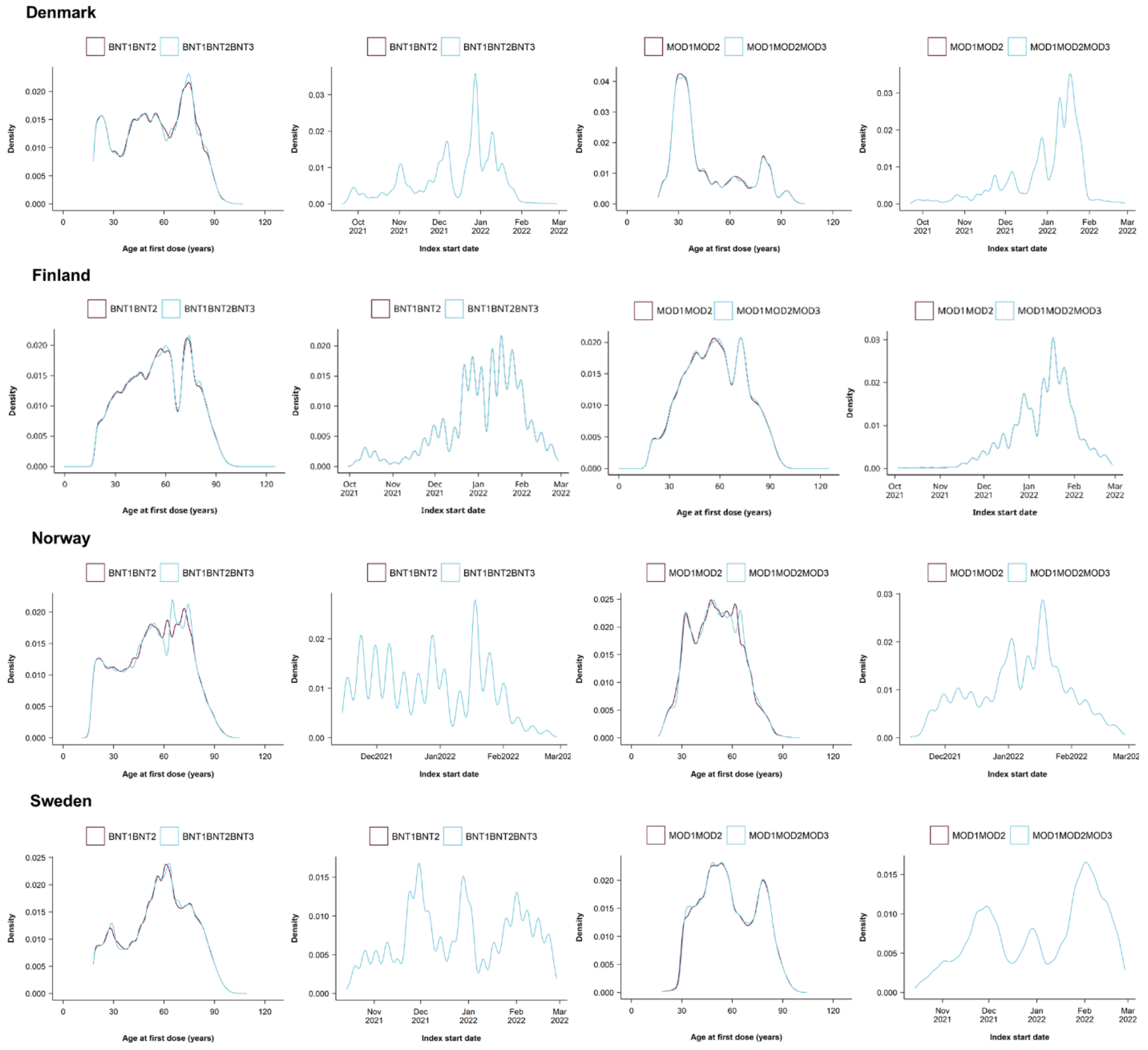

Note that only those individuals who had follow-up time during the country-specific periods of omicron variant predominance were included in the final matched comparisons.

**Supplementary Figure 10. Density plots of the distribution of age and index date for the weighted analyses comparing homologous MOD and BNT booster schedules.**

### Denmark

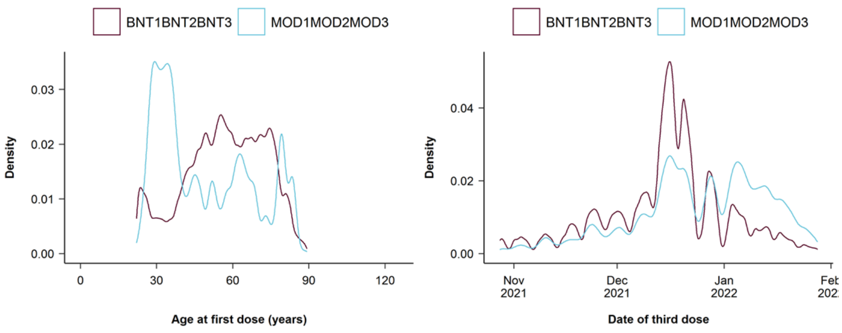

### Finland

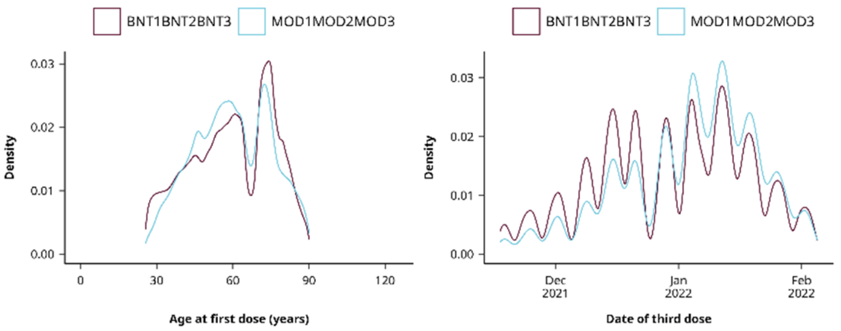

### Norway

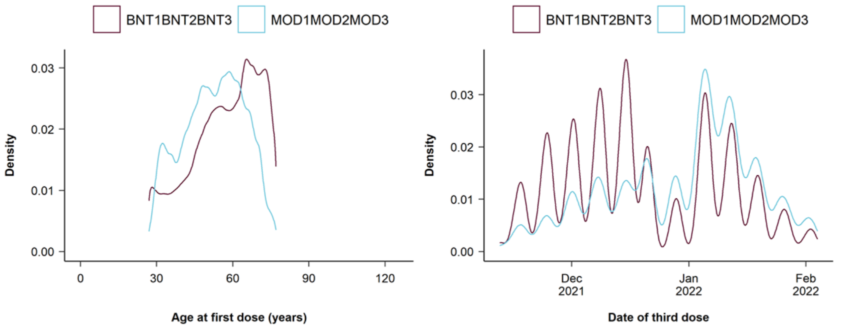

### Sweden

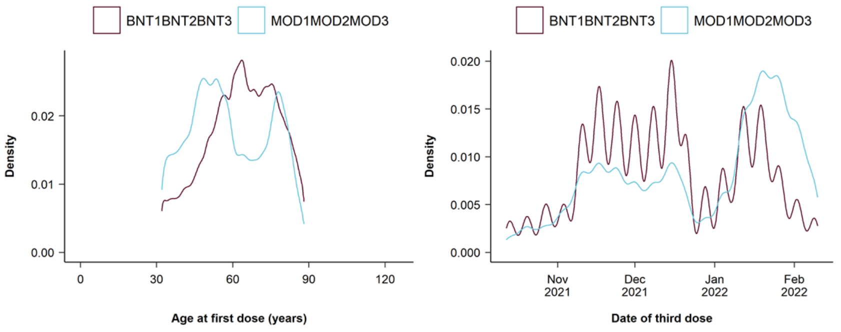

**Supplementary Figure 11. Cumulative incidence curves of documented SARS-CoV-2 infection comparing heterologous booster schedules that include the AZD1222 vaccine with matched primary schedules in each country.**

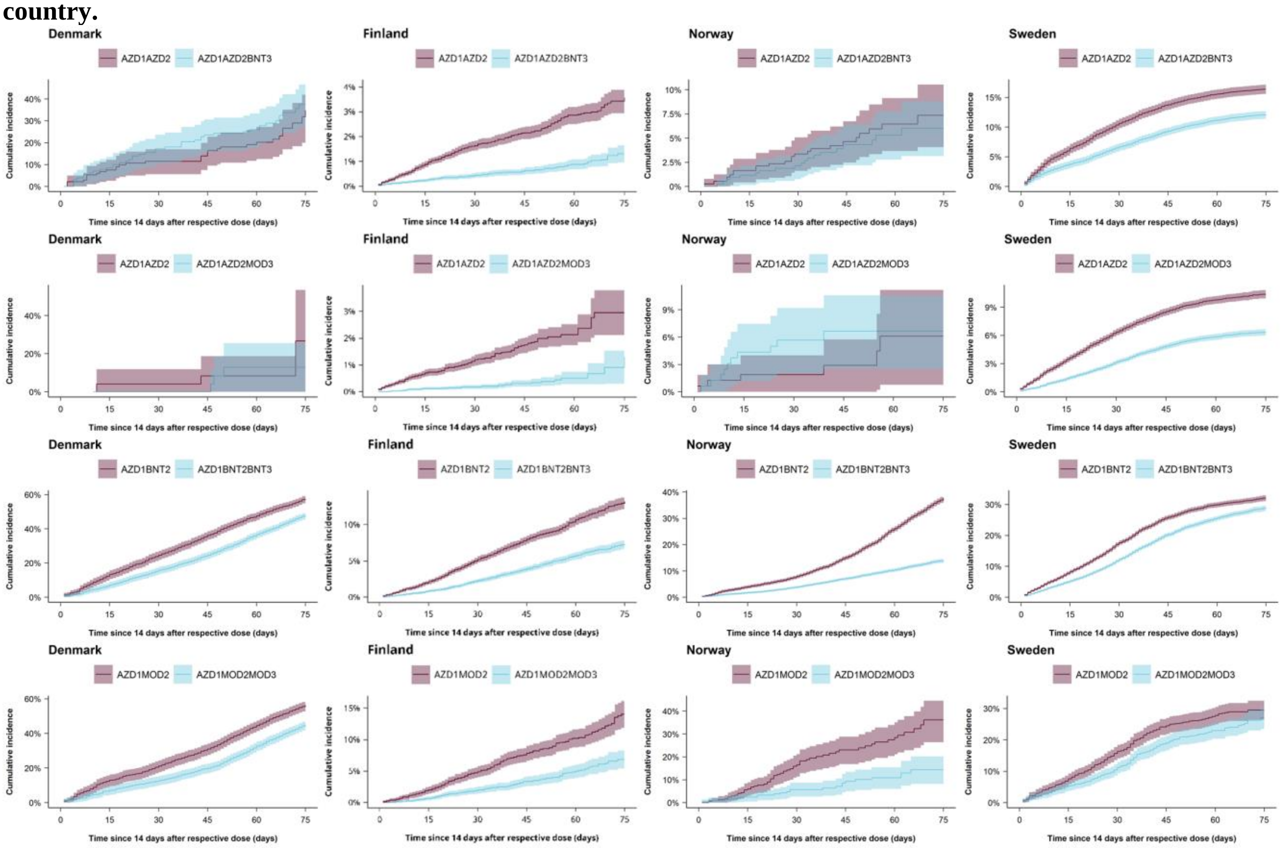

Columns are countries (Denmark, Finland, Norway, and Sweden) and rows are schedule comparisons.

**Supplementary Figure 12. Cumulative incidence curves of documented SARS-CoV-2 infection comparing heterologous booster schedules that include mRNA vaccines only with matched primary schedules in each country.**

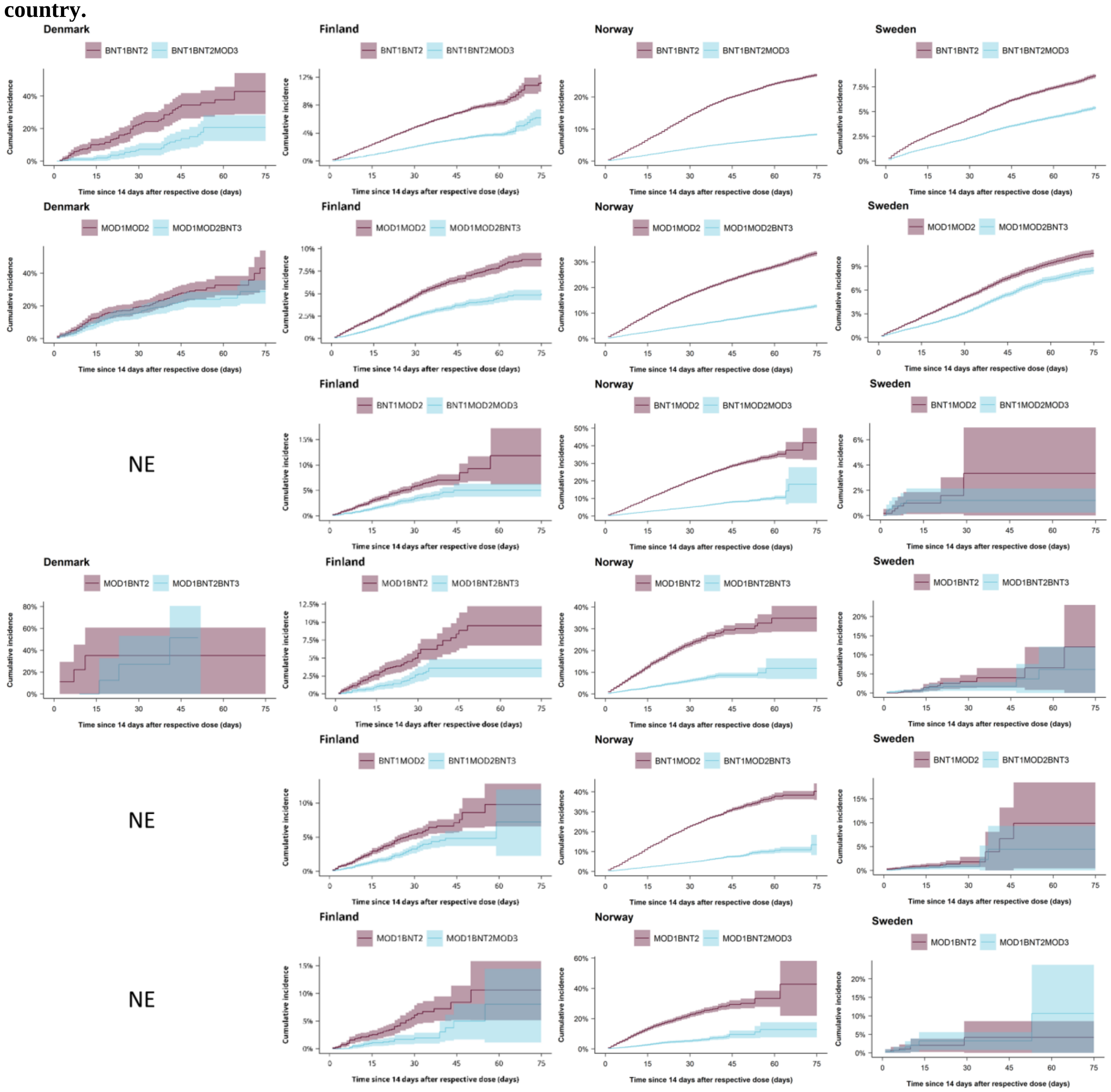

Columns are countries (Denmark, Finland, Norway, and Sweden) and rows are schedule comparisons. NE denotes not estimated.

**Supplementary Figure 13. Cumulative incidence curves of COVID-19 hospitalisation comparing heterologous booster schedules with matched primary schedules in each country.**

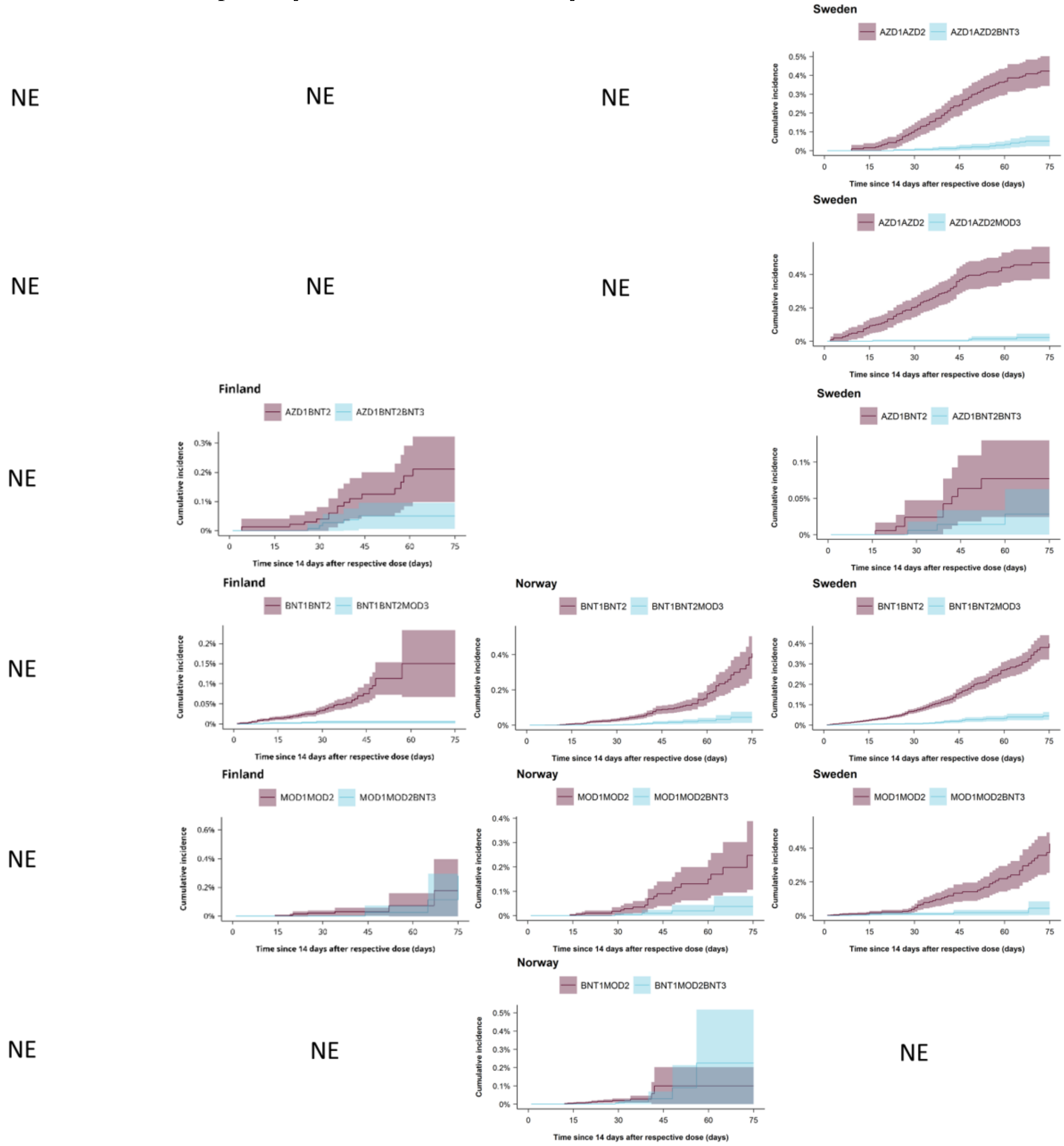

Columns are countries (Denmark, Finland, Norway, and Sweden) and rows are schedule comparisons. NE denotes not estimated.

**Supplementary Figure 14. Cumulative incidence curves of COVID-19 death comparing heterologous booster schedule of BNT1BNT2MOD3 with matched primary schedule of BNT1BNT2 in Sweden.**

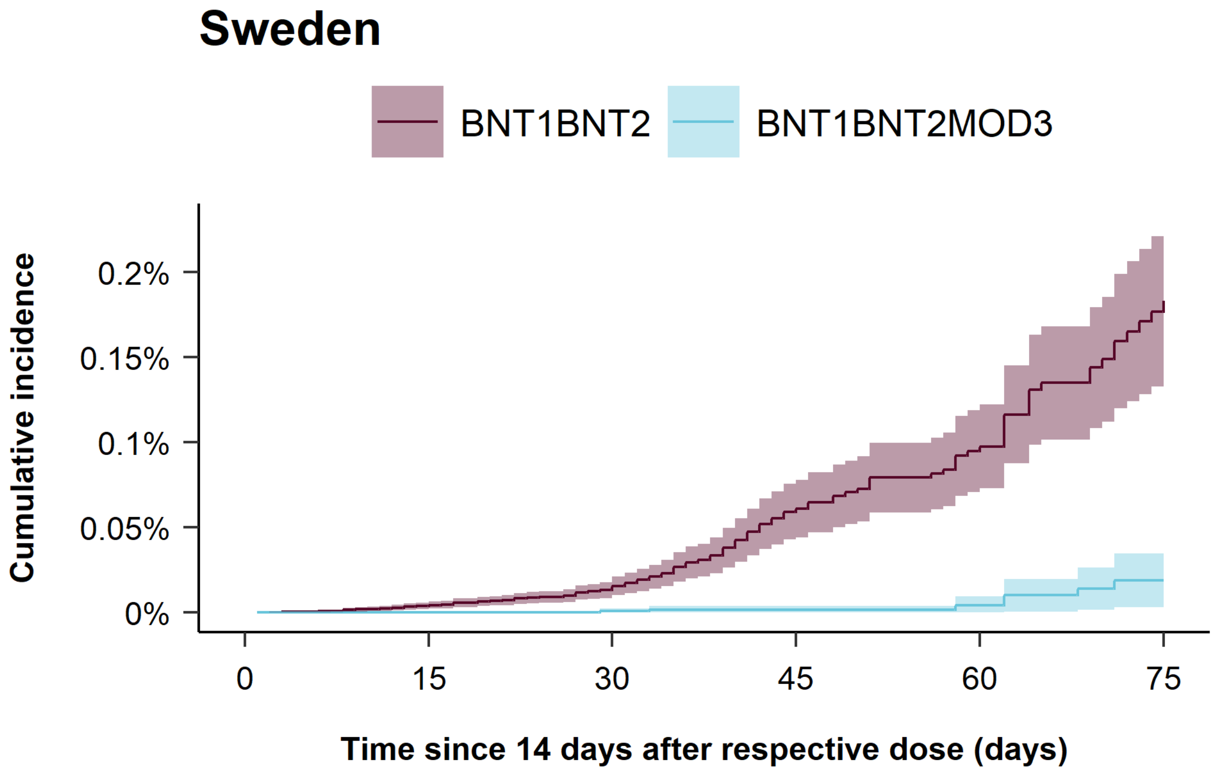

For the severe outcomes of COVID-19 related intensive care unit admission and death, due to low cell counts for the heterologous booster schedules vs primary schedules comparisons in each country and due to national regulations of data protection, only cumulative incidence curves for COVID-19 related death for BNT1BNT2MOD3 vs BNT1BNT2 in Sweden could be presented.

**Supplementary Table 11. Association between documented SARS-CoV-2 infection and heterologous booster schedules as compared with primary schedules.**

| | Studied schedule | Comparison schedule | Measures of association | |
|---|---|---|---|---|
| | Events/PYRS | Events/PYRS | RD (95% CI) per 100 individuals | CVE (95% CI) |
| AZD1AZD2BNT3 vs AZD1AZD2 | | | | |
| Denmark | 41 / 19.3 | 30 / 19.0 | 2.7 (-21.3 to 26.8) | -7.9% (-80.2% to 64.4%) |
| Finland | 144 / 2157.3 | 408 / 2089.1 | -2.2 (-2.8 to -1.6) | 62.8% (52.9% to 72.7%) |
| Norway | 21 / 56.9 | 21 / 56.8 | 0.5 (-3.8 to 4.9) | -8.6% (-79.8% to 62.6%) |
| Sweden | 2906 / 4788.7 | 3676 / 4546.6 | -4.3 (-5.5 to -3.1) | 26.3% (20.2% to 32.5%) |
| AZD1AZD2MOD3 vs AZD1AZD2 | | | | |
| Denmark | <5 / 3.7 | <5 / 3.7 | -13.8 (-61.8 to 34.3) | 51.6% (-49.9% to 100%) |
| Finland | 47 / 1478.1 | 234 / 1447.7 | -1.9 (-2.4 to -1.3) | 77.7% (67.9% to 87.4%) |
| Norway | 10 / 21.1 | 6 / 21.3 | 2.5 (-2.6 to 7.7) | -66.4% (-240.1% to 100%) |
| Sweden | 1426 / 3939.0 | 2258 / 3765.1 | -4.0 (-4.6 to -3.4) | 38.8% (34.2% to 43.3%) |
| AZD1BNT2BNT3 vs AZD1BNT2 | | | | |
| Denmark | 1477 / 428.2 | 1346 / 318.1 | -9.5 (-15.7 to -3.3) | 16.5% (6.9% to 26.1%) |
| Finland | 577 / 1607.9 | 1030 / 1490.9 | -5.7 (-6.9 to -4.4) | 42.7% (35.8% to 49.6%) |
| Norway | 1339 / 1846.2 | 3234 / 1698.7 | -22.4 (-24.4 to -20.4) | 60.9% (57.8% to 64.1%) |
| Sweden | 3529 / 1989.2 | 3976 / 1831.0 | -3.1 (-5.0 to -1.2) | 9.7% (4.1% to 15.3%) |
| AZD1MOD2MOD3 vs AZD1MOD2 | | | | |
| Denmark | 820 / 261.7 | 840 / 206.2 | -11.2 (-18.9 to -3.4) | 19.9% (7.8% to 31.9%) |
| Finland | 111 / 361.8 | 226 / 342.5 | -7.5 (-10.1 to -4.9) | 55.1% (42.9% to 67.2%) |
| Norway | 24 / 31.1 | 58 / 27.4 | -17.8 (-33.5 to -2.0) | 50.1% (19.1% to 81.0%) |
| Sweden | 272 / 174.0 | 323 / 160.5 | -2.4 (-8.9 to 4.1) | 8.1% (-13.3% to 29.4%) |
| BNT1BNT2MOD3 vs BNT1BNT2 | | | | |
| Denmark | 26 / 24.4 | 63 / 21.6 | -22.2 (-46.3 to 1.8) | 51.8% (18.0% to 85.6%) |
| Finland | 4664 / 19337.7 | 11625 / 18861.6 | -3.6 (-5.3 to -2.0) | 36.2% (21.9% to 50.4%) |
| Norway | 8136 / 16344.0 | 27130 / 15140.9 | -18.5 (-19.2 to -17.8) | 67.8% (66.3% to 69.2%) |
| Sweden | 8376 / 27888.2 | 14613 / 26971.1 | -3.2 (-3.5 to -2.9) | 37.4% (34.7% to 40.2%) |
| MOD1MOD2BNT3 vs MOD1MOD2 | | | | |
| Denmark | 88 / 45.2 | 106 / 40.4 | -14.3 (-37.5 to 9.0) | 33.2% (-6.8% to 73.2%) |
| Finland | 808 / 2763.7 | 1584 / 2699.2 | -4.1 (-5.3 to -2.8) | 44.2% (34.5% to 53.8%) |
| Norway | 2842 / 4287.8 | 8747 / 3875.5 | -21.3 (-22.9 to -19.7) | 62.3% (59.5% to 65.0%) |
| Sweden | 2286 / 5475.4 | 3263 / 5299.2 | -2.2 (-2.9 to -1.5) | 20.2% (14.3% to 26.1%) |
| BNT1MOD2MOD3 vs BNT1MOD2 | | | | |
| Denmark | <5 / 0.2 | <5 / 0.1 | NE | NE |
| Finland | 130 / 387.2 | 265 / 379.4 | -7.1 (-24.8 to 10.6) | 46.0% (-29.9% to 100%) |
| Norway | 2627 / 4015.3 | 9989 / 3639.2 | -32.3 (-56.1 to -8.6) | 69.7% (47.4% to 92.0%) |
| Sweden | 6 / 25.9 | 7 / 25.4 | -2.1 (-6.1 to 1.8) | 64.3% (14.1% to 100%) |
| MOD1BNT2BNT3 vs MOD1BNT2 | | | | |
| Denmark | <5 / 0.8 | <5 / 0.7 | NE | NE |
| Finland | 49 / 155.4 | 108 / 150.4 | 1.5 (-8.8 to 11.7) | -17.1% (-136.0% to 100%) |
| Norway | 403 / 456.3 | 1286 / 406.1 | -24.9 (-33.5 to -16.3) | 66.7% (56.0% to 77.4%) |
| Sweden | 14 / 56.6 | 18 / 55.7 | -5.9 (-20.8 to 8.9) | 49.1% (-29.0% to 100%) |
| BNT1MOD2BNT3 vs BNT1MOD2 | | | | |
| Denmark | NE | NE | NE | NE |
| Finland | 160 / 432.7 | 304 / 425.2 | -4.6 (-7.6 to -1.6) | 47.8% (27.7% to 68.0%) |

| | Studied schedule | Comparison schedule | Measures of association | |
|---|---|---|---|---|
| | Events/PYRS | Events/PYRS | RD (95% CI) per 100 individuals | CVE (95% CI) |
| Norway | 4517 / 7674.3 | 21389 / 6958.4 | -29.1 (-41.5 to -16.7) | 65.1% (48.7% to 81.5%) |
| Sweden | 13 / 88.0 | 23 / 87.1 | -5.5 (-16.8 to 5.8) | 55.4% (-14.4% to 100%) |
| MOD1BNT2MOD3 vs MOD1BNT2 | | | | |
| Denmark | NE | NE | NE | NE |
| Finland | 26 / 96.8 | 73 / 93.8 | -3.2 (-10.9 to 4.4) | 32.1% (-37.0% to 100%) |
| Norway | 185 / 251.7 | 687 / 220.0 | -25.7 (-35.9 to -15.4) | 69.8% (56.1% to 83.5%) |
| Sweden | 8 / 14.6 | 6 / 14.6 | 6.5 (-10.1 to 23.1) | NE |

CI denotes confidence interval, CVE comparative vaccine effectiveness, NE not estimated, PYRS person-years, and RD risk difference. Cell counts lower than 5 could not be reported due to national regulations on data privacy protection.

**Supplementary Table 12. Association between hospitalisation for COVID-19 and heterologous booster schedules as compared with primary schedules.**

| | Studied schedule | Comparison schedule | Measures of association | |
|---|---|---|---|---|
| | Events/PYRS | Events/PYRS | RD (95% CI) per 100 000 individuals | CVE (95% CI) |
| AZD1AZD2BNT3 vs AZD1AZD2 | | | | |
| Denmark | NE | NE | NE | NE |
| Finland | <5 / 2217.4 | 32 / 2112.1 | -266.3 (-431.9 to -100.6) | 95.5% (88.5% to 100%) |
| Norway | NE | NE | NE | NE |
| Sweden | 14 / 4952.3 | 119 / 4700.2 | -371.3 (-455.4 to -287.2) | 87.8% (80.9% to 94.7%) |
| AZD1AZD2MOD3 vs AZD1AZD2 | | | | |
| Denmark | NE | NE | NE | NE |
| Finland | NE | NE | NE | NE |
| Norway | NE | NE | NE | NE |
| Sweden | <5 / 4036.7 | 102 / 3863.5 | -447.5 (-546.7 to -348.4) | 95.2% (90.2% to 100%) |
| AZD1BNT2BNT3 vs AZD1BNT2 | | | | |
| Denmark | <5 / 506.8 | 0 / 373.2 | NE | NE |
| Finland | <5 / 1670.3 | 15 / 1537.9 | -199.5 (-341.5 to -57.5) | 86.6% (65.1% to 100%) |
| Norway | NE | NE | NE | NE |
| Sweden | <5 / 2163.3 | 9 / 2013.8 | -48.8 (-111.7 to 14.2) | 63.2% (12.1% to 100%) |
| AZD1MOD2MOD3 vs AZD1MOD2 | | | | |
| Denmark | <5 / 302.3 | <5 / 240.7 | -27.5 (-205.0 to 150.1) | 36.0% (-141.4% to 100%) |
| Finland | NE | NE | NE | NE |
| Norway | NE | NE | NE | NE |
| Sweden | NE | NE | NE | NE |
| BNT1BNT2MOD3 vs BNT1BNT2 | | | | |
| Denmark | NE | NE | NE | NE |
| Finland | 11 / 20589.8 | 93 / 19442.1 | -29.8 (-136.6 to 77.1) | 34.6% (-85.8% to 100%) |
| Norway | 11 / 17659.3 | 95 / 16407.6 | -341.1 (-460.4 to -221.9) | 94.6% (90.0% to 99.3%) |
| Sweden | 42 / 30369.9 | 306 / 28545.2 | -354.6 (-421.2 to -288.1) | 88.7% (83.6% to 93.8%) |
| MOD1MOD2BNT3 vs MOD1MOD2 | | | | |
| Denmark | NE | NE | NE | NE |
| Finland | 0 / 2888.9 | 10 / 2767.9 | NE | NE |
| Norway | <5 / 4816.0 | 21 / 4336.6 | -223.2 (-388.1 to -58.3) | 88.5% (70.8% to 100%) |
| Sweden | 9 / 5926.1 | 56 / 5610.8 | -382.1 (-521.9 to -242.2) | 89.6% (79.7% to 99.6%) |
| BNT1MOD2MOD3 vs BNT1MOD2 | | | | |
| Denmark | NE | NE | NE | NE |
| Finland | NE | NE | NE | NE |
| Norway | <5 / 4585.9 | 11 / 4152.2 | -46.7 (-80.5 to -12.9) | 95.5% (86.1% to 100%) |
| Sweden | NE | NE | NE | NE |
| MOD1BNT2BNT3 vs MOD1BNT2 | | | | |
| Denmark | NE | NE | NE | NE |
| Finland | NE | NE | NE | NE |
| Norway | NE | NE | NE | NE |
| Sweden | NE | NE | NE | NE |
| BNT1MOD2BNT3 vs BNT1MOD2 | | | | |
| Denmark | NE | NE | NE | NE |

| | Studied schedule | Comparison schedule | Measures of association | |
|---|---|---|---|---|
| | Events/PYRS | Events/PYRS | RD (95% CI) per 100 000 individuals | CVE (95% CI) |
| Finland | NE | NE | NE | NE |
| Norway | 8 / 8777.6 | 18 / 8028.6 | 128.8 (-138.1 to 395.7) | NE |
| Sweden | NE | NE | NE | NE |
| MOD1BNT2MOD3 vs MOD1BNT2 | | | | |
| Denmark | NE | NE | NE | NE |
| Finland | NE | NE | NE | NE |
| Norway | NE | NE | NE | NE |
| Sweden | NE | NE | NE | NE |

CI denotes confidence interval, CVE comparative vaccine effectiveness, NE not estimated, PYRS person-years, and RD risk difference. Cell counts lower than 5 could not be reported due to national regulations on data privacy protection.

**Supplementary Table 13. Association between COVID-19 intensive care unit admission and COVID-19 death and heterologous booster schedules as compared with primary schedules.**

| | Studied schedule | Comparison schedule | Measures of association | |
|---|---|---|---|---|
| | Events/PYRS | Events/PYRS | RD (95% CI) per 100 000 individuals | CVE (95% CI) |
| AZD1AZD2BNT3 vs AZD1AZD2 | | | | |
| ICU admission | | | | |
| Denmark | NE | NE | NE | NE |
| Finland | 0 / 2219.4 | 8 / 2112.9 | NE | NE |
| Norway | NE | NE | NE | NE |
| Sweden | NE | NE | NE | NE |
| Death | | | | |
| Denmark | NE | NE | NE | NE |
| Finland | 0 / 2275.2 | 14 / 2156.7 | NE | NE |
| Norway | NE | NE | NE | NE |
| Sweden | NE | NE | NE | NE |
| AZD1AZD2MOD3 vs AZD1AZD2 | | | | |
| ICU admission | | | | |
| Denmark | NE | NE | NE | NE |
| Finland | NE | NE | NE | NE |
| Norway | NE | NE | NE | NE |
| Sweden | NE | NE | NE | NE |
| Death | | | | |
| Denmark | NE | NE | NE | NE |
| Finland | <5 / 1584.7 | 9 / 1499.6 | -60.7 (-133.0 to 11.6) | 73.7% (18.8% to 100%) |
| Norway | NE | NE | NE | NE |
| Sweden | NE | NE | NE | NE |
| AZD1BNT2BNT3 vs AZD1BNT2 | | | | |
| ICU admission | | | | |
| Denmark | NE | NE | NE | NE |
| Finland | <5 / 1670.6 | <5 / 1538.2 | -34.8 (-136.6 to 66.9) | 52.8% (-42.8% to 100%) |
| Norway | NE | NE | NE | NE |
| Sweden | NE | NE | NE | NE |
| Death | | | | |
| Denmark | NE | NE | NE | NE |
| Finland | <5 / 1725.0 | <5 / 1599.3 | 6.6 (-85.4 to 98.5) | -11.9% (-186.4% to 100%) |
| Norway | NE | NE | NE | NE |
| Sweden | NE | NE | NE | NE |
| AZD1MOD2MOD3 vs AZD1MOD2 | | | | |
| ICU admission | | | | |
| Denmark | NE | NE | NE | NE |
| Finland | NE | NE | NE | NE |
| Norway | NE | NE | NE | NE |
| Sweden | NE | NE | NE | NE |
| Death | | | | |
| Denmark | NE | NE | NE | NE |
| Finland | NE | NE | NE | NE |
| Norway | NE | NE | NE | NE |

| | Studied schedule | Comparison schedule | Measures of association | |
|---|---|---|---|---|
| | Events/PYRS | Events/PYRS | RD (95% CI) per 100 000 individuals | CVE (95% CI) |
| Sweden | NE | NE | NE | NE |
| BNT1BNT2MOD3 vs BNT1BNT2 | | | | |
| ICU admission | | | | |
| Denmark | NE | NE | NE | NE |
| Finland | <5 / 20594.5 | 6 / 19445.2 | -2.1 (-8.3 to 4.2) | 41.4% (-47.7% to 100%) |
| Norway | NE | NE | NE | NE |
| Sweden | NE | NE | NE | NE |
| Death | | | | |
| Denmark | NE | NE | NE | NE |
| Finland | <5 / 22423.8 | 71 / 20654.6 | -260.5 (-513.7 to -7.3) | 99.3% (98.2% to 100%) |
| Norway | NE | NE | NE | NE |
| Sweden | NE | NE | NE | NE |
| MOD1MOD2BNT3 vs MOD1MOD2 | | | | |
| ICU admission | | | | |
| Denmark | NE | NE | NE | NE |
| Finland | NE | NE | NE | NE |
| Norway | NE | NE | NE | NE |
| Sweden | NE | NE | NE | NE |
| Death | | | | |
| Denmark | NE | NE | NE | NE |
| Finland | 0 / 3047.1 | 7 / 2884.3 | NE | NE |
| Norway | NE | NE | NE | NE |
| Sweden | NE | NE | NE | NE |
| BNT1MOD2MOD3 vs BNT1MOD2 | | | | |
| ICU admission | | | | |
| Denmark | NE | NE | NE | NE |
| Finland | NE | NE | NE | NE |
| Norway | <5 / 4585.9 | <5 / 4152.2 | -8.3 (-24.4 to 7.8) | 75.5% (16.8% to 100%) |
| Sweden | NE | NE | NE | NE |
| Death | | | | |
| Denmark | NE | NE | NE | NE |
| Finland | NE | NE | NE | NE |
| Norway | NE | NE | NE | NE |
| Sweden | NE | NE | NE | NE |
| MOD1BNT2BNT3 vs MOD1BNT2 | | | | |
| ICU admission | | | | |
| Denmark | NE | NE | NE | NE |
| Finland | NE | NE | NE | NE |
| Norway | NE | NE | NE | NE |
| Sweden | NE | NE | NE | NE |
| Death | | | | |
| Denmark | NE | NE | NE | NE |
| Finland | NE | NE | NE | NE |
| Norway | NE | NE | NE | NE |

| | Studied schedule | Comparison schedule | Measures of association | |
|---|---|---|---|---|
| | Events/PYRS | Events/PYRS | RD (95% CI) per 100 000 individuals | CVE (95% CI) |
| Sweden | NE | NE | NE | NE |
| BNT1MOD2BNT3 vs BNT1MOD2 | | | | |
| ICU admission | | | | |
| Denmark | NE | NE | NE | NE |
| Finland | NE | NE | NE | NE |
| Norway | NE | NE | NE | NE |
| Sweden | NE | NE | NE | NE |
| Death | | | | |
| Denmark | NE | NE | NE | NE |
| Finland | NE | NE | NE | NE |
| Norway | NE | NE | NE | NE |
| Sweden | NE | NE | NE | NE |
| MOD1BNT2MOD3 vs MOD1BNT2 | | | | |
| ICU admission | | | | |
| Denmark | NE | NE | NE | NE |
| Finland | NE | NE | NE | NE |
| Norway | NE | NE | NE | NE |
| Sweden | NE | NE | NE | NE |
| Death | | | | |
| Denmark | NE | NE | NE | NE |
| Finland | NE | NE | NE | NE |
| Norway | NE | NE | NE | NE |
| Sweden | NE | NE | NE | NE |

CI denotes confidence interval, CVE comparative vaccine effectiveness, ICU intensive care unit, NE not estimated, PYRS person-years, and RD risk difference. Cell counts lower than 5 could not be reported due to national regulations on data privacy protection.

**Supplementary Figure 15. Cumulative incidence curves of COVID-19 outcomes comparing homologous booster schedules with matched primary schedules in each country.**

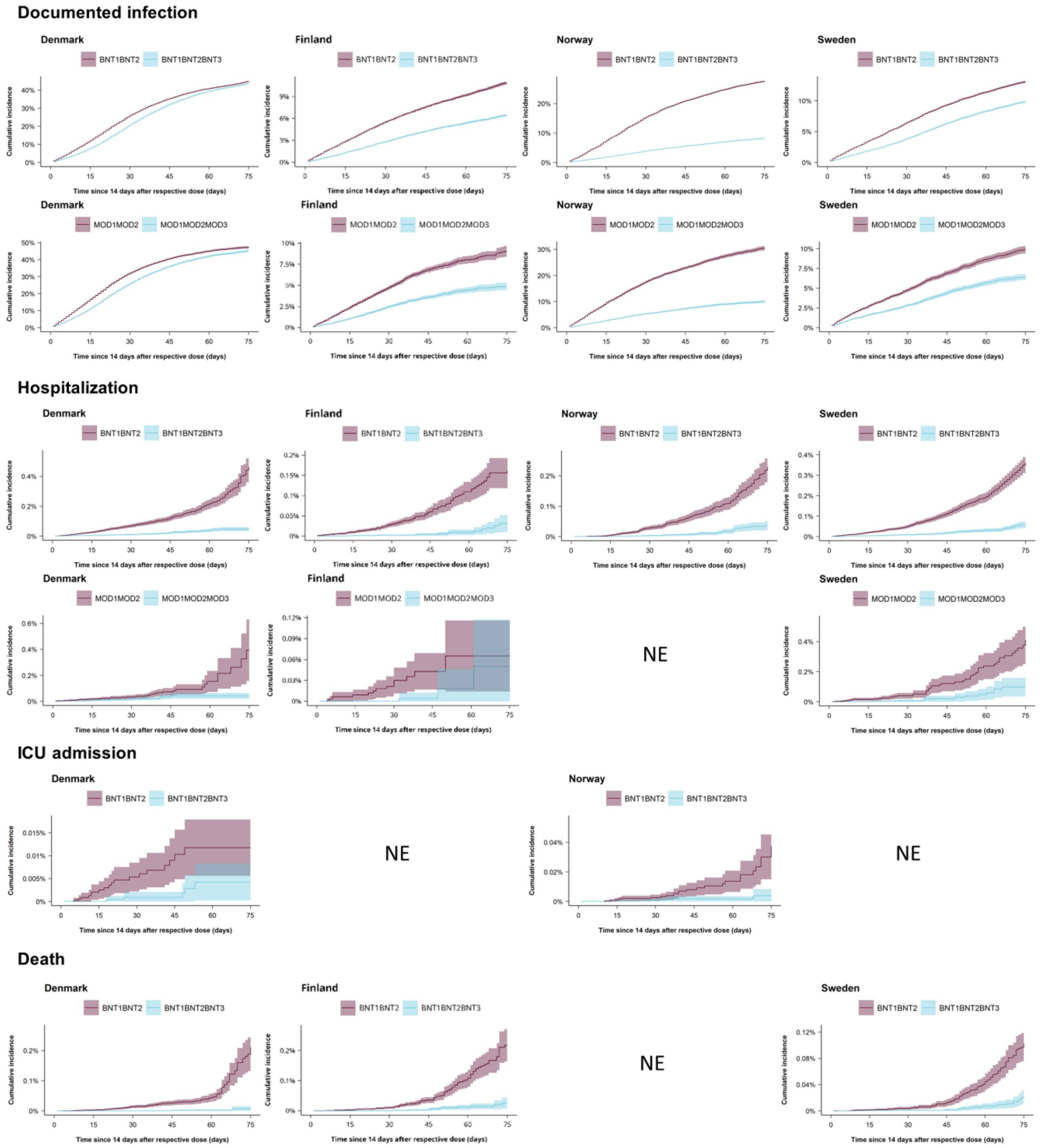

Columns are countries (Denmark, Finland, Norway, and Sweden) and rows are schedule comparisons and outcomes. ICU denotes intensive care unit and NE not estimated.

**Supplementary Table 14. Association between COVID-19 outcomes and homologous booster schedules as compared with primary schedules.**

| | Studied schedule | Comparison schedule | Measures of association | |
|---|---|---|---|---|
| | Events/PYRS | Events/PYRS | RD (95% CI) per 100 or per 100 000 individuals[a] | CVE (95% CI) |
| BNT1BNT2BNT3 vs BNT1BNT2 | | | | |
| Documented infection | | | | |
| Denmark | 72537 / 26153.7 | 75846 / 23206.0 | -1.3 (-2.1 to -0.4) | 2.8% (1.0% to 4.6%) |
| Finland | 12822 / 36465.5 | 23564 / 35174.5 | -4.2 (-4.5 to -3.9) | 38.6% (36.2% to 40.9%) |
| Norway | 14775 / 32378.1 | 54322 / 29777.0 | -19.6 (-20.0 to -19.2) | 70.2% (69.5% to 71.0%) |
| Sweden | 21284 / 42933.3 | 31553 / 41033.9 | -3.2 (-3.5 to -3.0) | 24.5% (22.8% to 26.2%) |
| Hospitalisation | | | | |
| Denmark | 65 / 33657.6 | 303 / 27311.4 | -411.5 (-498.2 to -324.8) | 88.7% (84.2% to 93.2%) |
| Finland | 28 / 39011.2 | 161 / 36350.7 | -129.1 (-174.5 to -83.8) | 77.9% (65.2% to 90.6%) |
| Norway | 33 / 34490.0 | 148 / 32053.2 | -160.1 (-202.5 to -117.8) | 80.1% (70.8% to 89.3%) |
| Sweden | 81 / 46488.5 | 449 / 43626.3 | -302.1 (-345.1 to -259.2) | 83.7% (79.0% to 88.4%) |
| ICU admission | | | | |
| Denmark | 5 / 33687.1 | 18 / 27313.9 | -7.5 (-14.9 to -0.2) | 64.1% (25.2% to 100%) |
| Finland | NE | NE | NE | NE |
| Norway | <5 / 34493.1 | 19 / 32043.3 | -29.8 (-46.4 to -13.1) | 92.2% (81.6% to 100%) |
| Sweden | NE | NE | NE | NE |
| Death | | | | |
| Denmark | 6 / 41366.0 | 94 / 33499.5 | -203.5 (-265.6 to -141.4) | 96.8% (92.6% to 100%) |
| Finland | 18 / 41796.5 | 124 / 38503.1 | -190.3 (-251.9 to -128.7) | 80.9% (69.5% to 92.4%) |
| Norway | NE | NE | NE | NE |
| Sweden | NE | NE | NE | NE |
| MOD1MOD2MOD3 vs MOD1MOD2 | | | | |
| Documented infection | | | | |
| Denmark | 19826 / 5854.0 | 21869 / 5175.7 | -2.2 (-4.2 to -0.3) | 4.7% (0.6% to 8.7%) |
| Finland | 1309 / 4463.7 | 2550 / 4341.7 | -4.3 (-5.1 to -3.5) | 46.6% (40.4% to 52.9%) |
| Norway | 2611 / 4175.2 | 8122 / 3760.3 | -20.1 (-21.5 to -18.8) | 65.9% (63.3% to 68.4%) |
| Sweden | 1818 / 5045.6 | 2901 / 4869.6 | -3.4 (-4.1 to -2.8) | 34.7% (29.3% to 40.0%) |
| Hospitalisation | | | | |
| Denmark | 18 / 7710.9 | 41 / 6376.6 | -352.5 (-590.8 to -114.2) | 89.3% (80.8% to 97.9%) |
| Finland | <5 / 4690.1 | 15 / 4457.3 | -60.3 (-124.6 to 4.0) | 76.5% (36.5% to 100%) |
| Norway | <5 / 4657.0 | 28 / 4161.0 | -300.8 (-467.0 to -134.5) | 94.4% (85.5% to 100%) |
| Sweden | 12 / 5477.3 | 52 / 5154.8 | -313.2 (-460.9 to -165.4) | 76.3% (59.5% to 93.1%) |
| ICU admission | | | | |
| Denmark | <5 / 7724.9 | 0 / 6374.8 | NE | NE |
| Finland | NE | NE | NE | NE |
| Norway | <5 / 4663.0 | 5 / 4167.3 | -55.3 (-127.9 to 17.3) | 94.0% (80.2% to 100%) |
| Sweden | NE | NE | NE | NE |
| Death | | | | |
| Denmark | <5 / 9638.2 | 7 / 7990.2 | -131.0 (-268.8 to 6.9) | 99.0% (96.9% to 100%) |
| Finland | <5 / 4901.6 | 16 / 4641.7 | -144.6 (-321.5 to 32.3) | 87.8% (60.7% to 100%) |
| Norway | NE | NE | NE | NE |
| Sweden | NE | NE | NE | NE |

CI denotes confidence interval, CVE comparative vaccine effectiveness, ICU intensive care unit, NE not estimated, PYRS person-years, and RD risk difference. Cell counts lower than 5 could not be reported due to national regulations on data privacy protection. [a]Risk differences for documented SARS-CoV-2 infection are reported per 100 individuals while risk differences for the severe COVID-19 outcomes of hospitalization, intensive care unit admission, and death are reported per 100 000 individuals.

**Supplementary Figure 16. Cumulative incidence curves of documented SARS-CoV-2 infection comparing heterologous that include the AZD1222 vaccine and homologous booster schedules in each country.**

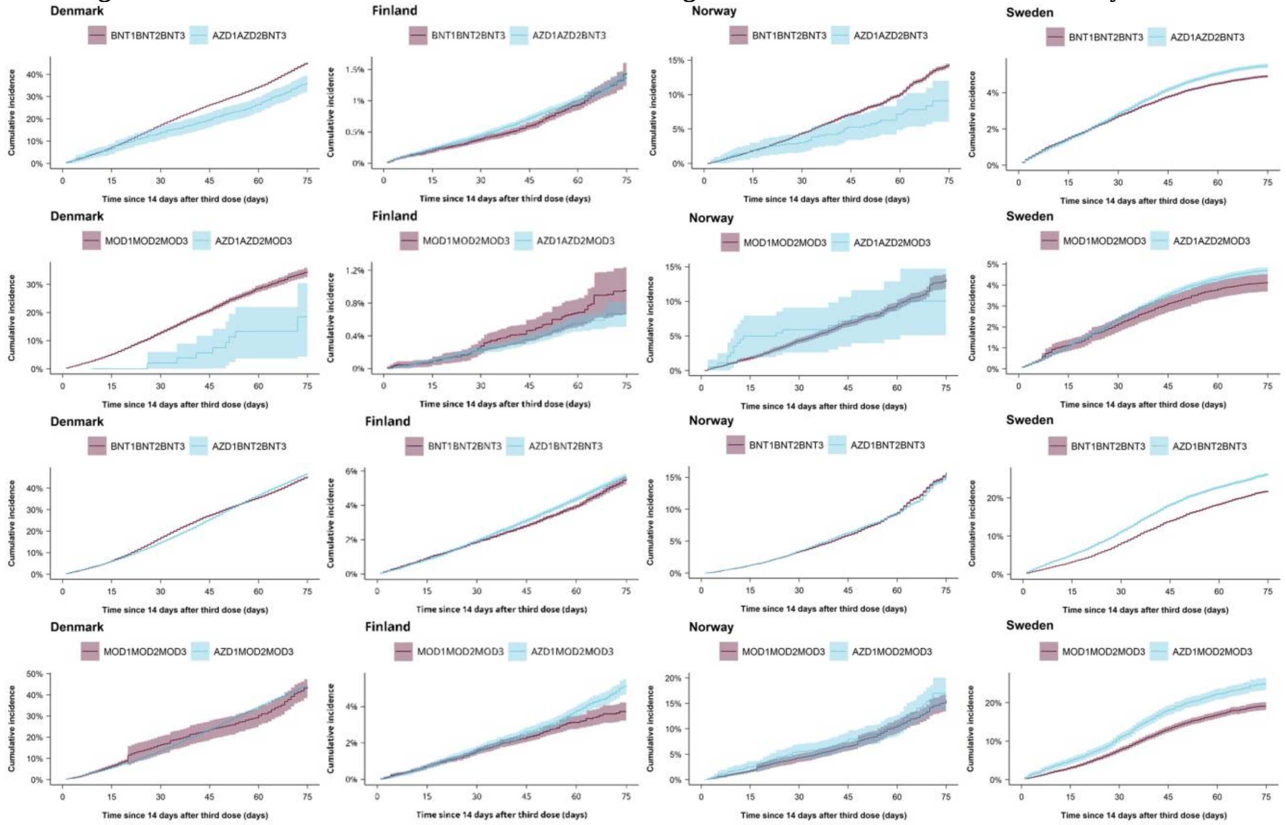

Columns are countries (Denmark, Finland, Norway, and Sweden) and rows are schedule comparisons.

**Supplementary Figure 17. Cumulative incidence curves of documented SARS-CoV-2 infection comparing heterologous that include mRNA vaccines only and homologous booster schedules in each country.**

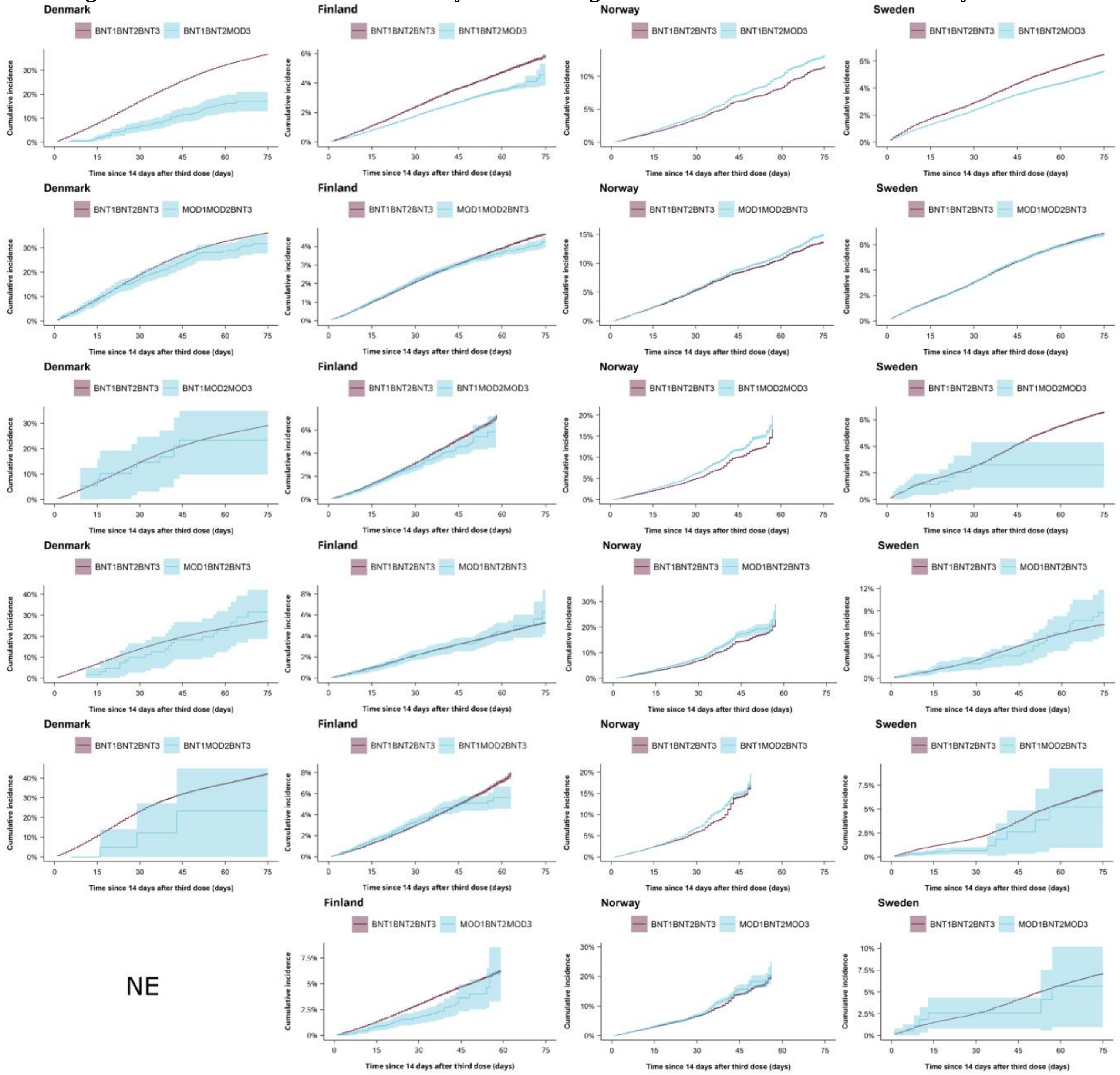

Columns are countries (Denmark, Finland, Norway, and Sweden) and rows are schedule comparisons. NE denotes not estimated.

**Supplementary Figure 18. Cumulative incidence curves of COVID-19 hospitalisation comparing heterologous and homologous booster schedules in each country.**

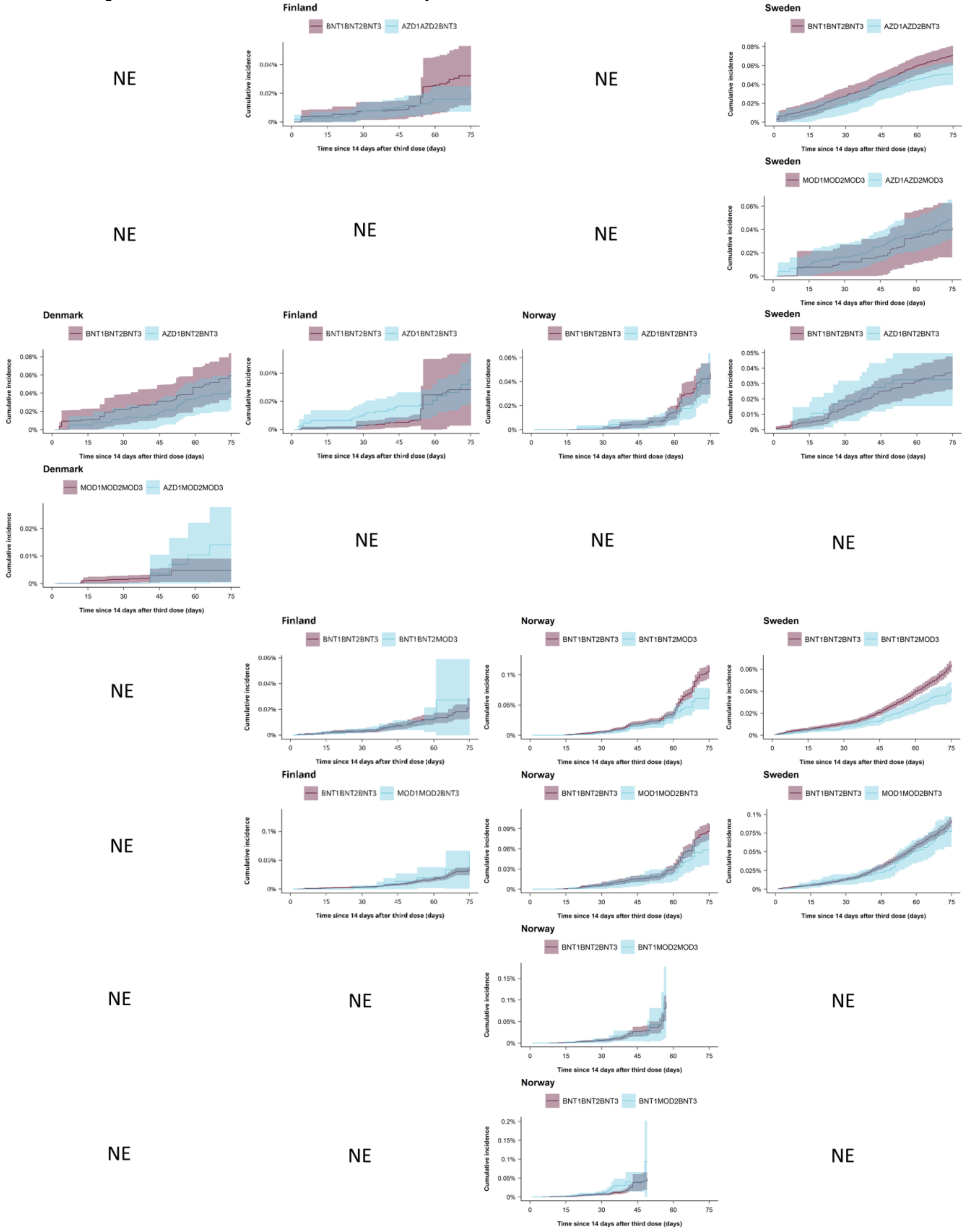

Columns are countries (Denmark, Finland, Norway, and Sweden) and rows are schedule comparisons. NE denotes not estimated.

**Supplementary Figure 19. Cumulative incidence curves of COVID-19 intensive care unit admission and COVID-19 death comparing heterologous and homologous booster schedules in each country.**

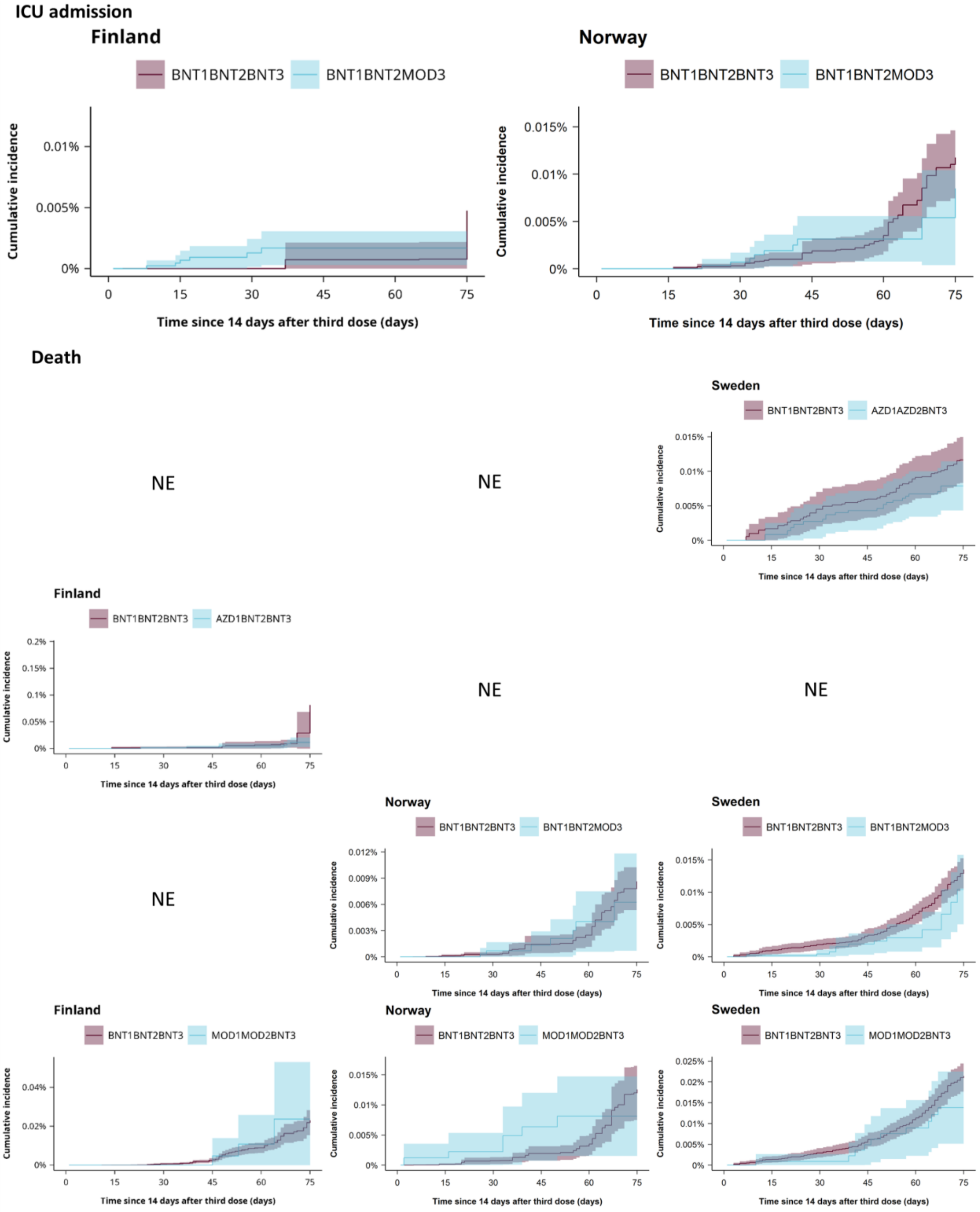

Columns are countries and rows are schedule comparisons and outcomes. ICU denotes intensive care unit and denotes not estimated. Cell counts were too low to compute cumulative incidence curves of ICU admission and death for any comparison in Denmark as well as ICU admission in Sweden.

**Supplementary Table 15. Association between documented SARS-CoV-2 infection and heterologous booster schedules as compared with homologous booster schedules.**

| | Studied schedule | Comparison schedule | Measures of association | |
|---|---|---|---|---|
| | Events/PYRS | Events/PYRS | RD (95% CI) per 100 individuals | CVE (95% CI) |
| AZD1AZD2BNT3 vs BNT1BNT2BNT3 | | | | |
| Denmark | 229 / 106.7 | 517302 / 210481.9 | -9.1 (-12.9 to -5.3) | 20.0% (11.7% to 28.4%) |
| Finland | 986 / 15457.7 | 2356 / 47969.2 | -0.1 (-0.3 to 0.1) | 4.1% (-9.9% to 18.2%) |
| Norway | 41 / 92.0 | 68585 / 166278.5 | -5.4 (-8.4 to -2.4) | 37.2% (16.6% to 57.8%) |
| Sweden | 12843 / 48515.0 | 58756 / 152425.3 | 0.6 (0.4 to 0.7) | -11.8% (-14.8% to -8.9%) |
| AZD1AZD2MOD3 vs MOD1MOD2MOD3 | | | | |
| Denmark | 8 / 8.3 | 75144 / 24196.7 | -16.1 (-29.2 to -3.0) | 46.7% (9.0% to 84.4%) |
| Finland | 176 / 5665.3 | 187 / 4729.5 | -0.3 (-0.6 to 0.0) | 29.5% (3.1% to 56.0%) |
| Norway | 17 / 29.3 | 10959 / 16424.1 | -3.1 (-8.1 to 1.8) | 23.8% (-13.2% to 60.7%) |
| Sweden | 3915 / 16218.7 | 3707 / 12528.7 | 0.6 (0.1 to 1.0) | -13.8% (-25.9% to -1.7%) |
| AZD1BNT2BNT3 vs BNT1BNT2BNT3 | | | | |
| Denmark | 23844 / 7275.4 | 414234 / 156233.3 | 1.5 (0.9 to 2.1) | -3.3% (-4.6% to -2.0%) |
| Finland | 4048 / 14038.0 | 30942 / 90440.2 | 0.2 (-0.1 to 0.5) | -3.5% (-9.0% to 2.0%) |
| Norway | 10967 / 14713.4 | 54798 / 94786.2 | -0.4 (-0.8 to 0.0) | 2.4% (-0.1% to 4.8%) |
| Sweden | 10752 / 6894.9 | 63323 / 145842.6 | 4.4 (3.8 to 4.9) | -20.0% (-22.6% to -17.4%) |
| AZD1MOD2MOD3 vs MOD1MOD2MOD3 | | | | |
| Denmark | 13053 / 4220.7 | 54891 / 18868.7 | 0.9 (-3.6 to 5.4) | -2.1% (-12.6% to 8.4%) |
| Finland | 625 / 2534.1 | 2871 / 11136.5 | 1.4 (0.8 to 2.1) | -38.7% (-60.2% to -17.2%) |
| Norway | 119 / 149.9 | 10078 / 12448.1 | 1.4 (-2.2 to 4.9) | -8.8% (-32.0% to 14.4%) |
| Sweden | 846 / 563.9 | 4155 / 11813.5 | 5.8 (3.8 to 7.7) | -30.2% (-41.3% to -19.1%) |
| BNT1BNT2MOD3 vs BNT1BNT2BNT3 | | | | |
| Denmark | 67 / 68.1 | 580653 / 285991.3 | -19.8 (-23.9 to -15.8) | 53.8% (42.8% to 64.8%) |
| Finland | 9045 / 41087.3 | 35133 / 136227.2 | -1.4 (-2.1 to -0.7) | 23.7% (12.1% to 35.2%) |
| Norway | 22755 / 37011.0 | 60448 / 164700.4 | 1.7 (1.5 to 1.9) | -14.6% (-16.7% to -12.6%) |
| Sweden | 17858 / 63874.3 | 66697 / 186663.1 | -1.3 (-1.4 to -1.1) | 19.3% (17.8% to 20.8%) |
| MOD1MOD2BNT3 vs BNT1BNT2BNT3 | | | | |
| Denmark | 215 / 103.1 | 586998 / 304529.9 | -4.6 (-8.4 to -0.7) | 12.6% (1.9% to 23.2%) |
| Finland | 1531 / 6288.5 | 37728 / 154405.4 | -0.4 (-0.7 to -0.1) | 8.7% (2.4% to 15.1%) |
| Norway | 10124 / 13306.3 | 70657 / 190310.5 | 1.3 (0.9 to 1.6) | -9.3% (-11.9% to -6.8%) |
| Sweden | 5744 / 16309.7 | 74728 / 202021.2 | -0.1 (-0.3 to 0.0) | 2.0% (-0.6% to 4.6%) |
| BNT1MOD2MOD3 vs BNT1BNT2BNT3 | | | | |
| Denmark | 10 / 6.4 | 573968 / 297012.1 | -5.8 (-18.2 to 6.6) | 19.9% (-22.6% to 62.4%) |
| Finland | 284 / 793.9 | 28741 / 89021.6 | -1.4 (-2.8 to 0.0) | 19.9% (0.9% to 38.9%) |
| Norway | 6995 / 7980.9 | 30755 / 47764.3 | 3.2 (2.2 to 4.3) | -19.3% (-25.6% to -13.0%) |
| Sweden | 12 / 48.2 | 66067 / 177793.9 | -4.0 (-5.7 to -2.3) | 60.4% (34.4% to 86.3%) |
| MOD1BNT2BNT3 vs BNT1BNT2BNT3 | | | | |
| Denmark | 20 / 11.1 | 595378 / 306951.1 | 3.9 (-7.8 to 15.6) | -14.1% (-56.5% to 28.3%) |
| Finland | 119 / 441.6 | 37126 / 146707.4 | 1.1 (-1.1 to 3.2) | -19.9% (-61.0% to 21.2%) |
| Norway | 1064 / 930.9 | 38236 / 55002.8 | 6.2 (0.3 to 12.1) | -27.0% (-52.8% to -1.2%) |
| Sweden | 41 / 119.7 | 74260 / 200193.9 | 1.5 (-1.6 to 4.7) | -21.3% (-64.7% to 22.2%) |
| BNT1MOD2BNT3 vs BNT1BNT2BNT3 | | | | |

| | Studied schedule | Comparison schedule | Measures of association | |
|---|---|---|---|---|
| | Events/PYRS | Events/PYRS | RD (95% CI) per 100 individuals | CVE (95% CI) |
| Denmark | <5 / 2.3 | 584116 / 294224.6 | -19.0 (-44.5 to 6.5) | 44.9% (-15.5% to 100%) |
| Finland | 305 / 751.1 | 31999 / 101003.0 | -2.3 (-3.5 to -1.2) | 29.0% (15.4% to 42.7%) |
| Norway | 10660 / 12518.7 | 32303 / 43862.6 | 1.4 (-0.1 to 2.9) | -7.8% (-16.0% to 0.4%) |
| Sweden | 19 / 142.3 | 52427 / 123389.4 | -1.8 (-6.0 to 2.3) | 26.0% (-33.1% to 85.2%) |
| MOD1BNT2MOD3 vs BNT1BNT2BNT3 | | | | |
| Denmark | 0 / 1.6 | 564709 / 296644.2 | | |
| Finland | 47 / 178.8 | 28770 / 91758.2 | -0.3 (-3.0 to 2.3) | 5.2% (-36.7% to 47.1%) |
| Norway | 649 / 612.5 | 32573 / 50075.2 | 2.9 (-2.4 to 8.1) | -12.9% (-36.6% to 10.7%) |
| Sweden | 10 / 27.2 | 67166 / 184851.6 | -1.4 (-6.0 to 3.1) | 20.1% (-44.2% to 84.4%) |

CI denotes confidence interval, CVE comparative vaccine effectiveness, NE not estimated, PYRS person-years, and RD risk difference. Cell counts lower than 5 could not be reported due to national regulations on data privacy protection.

**Supplementary Table 16. Association between hospitalisation for COVID-19 and heterologous booster schedules as compared with homologous booster schedules.**

| | Studied schedule | Comparison schedule | Measures of association | |
|---|---|---|---|---|
| | Events/PYRS | Events/PYRS | RD per 100 000 individuals | CVE (95% CI) |
| AZD1AZD2BNT3 vs BNT1BNT2BNT3 | | | | |
| Denmark | 0 / 114.4 | 506 / 227582.4 | NE | NE |
| Finland | 15 / 15491.4 | 53 / 48048.9 | -10.0 (-34.7 to 14.6) | 30.8% (-29.3% to 90.9%) |
| Norway | <5 / 92.4 | 492 / 162942.5 | 337.7 (-470.7 to 1146.1) | NE |
| Sweden | 129 / 49118.2 | 459 / 155561.6 | -19.4 (-35.1 to -3.6) | 27.5% (7.6% to 47.4%) |
| AZD1AZD2MOD3 vs MOD1MOD2MOD3 | | | | |
| Denmark | 0 / 8.7 | 44 / 27090.6 | NE | NE |
| Finland | <5 / 5671.0 | <5 / 4735.3 | -15.2 (-65.3 to 34.8) | 59.9% (-35.8% to 100%) |
| Norway | 0 / 29.8 | 31 / 16740.9 | NE | NE |
| Sweden | 38 / 16375.9 | 28 / 12723.0 | 7.2 (-21.9 to 36.2) | -17.3% (-95.7% to 61.1%) |
| AZD1BNT2BNT3 vs BNT1BNT2BNT3 | | | | |
| Denmark | 18 / 8111.6 | 366 / 173397.9 | -17.5 (-49.2 to 14.2) | 29.4% (-15.4% to 74.2%) |
| Finland | 21 / 14175.5 | 41 / 91387.9 | 5.1 (-25.7 to 35.9) | -16.7% (-130.9% to 97.4%) |
| Norway | 24 / 15017.0 | 255 / 96254.4 | -3.2 (-27.0 to 20.7) | 6.8% (-42.9% to 56.4%) |
| Sweden | 14 / 7315.4 | 281 / 148817.3 | -4.3 (-24.6 to 16.0) | 11.7% (-41.8% to 65.1%) |
| AZD1MOD2MOD3 vs MOD1MOD2MOD3 | | | | |
| Denmark | <5 / 4662.1 | 29 / 21889.2 | 9.2 (-5.2 to 23.6) | NE |
| Finland | 0 / 2554.0 | <5 / 11218.9 | NE | NE |
| Norway | 0 / 153.1 | 18 / 12744.1 | NE | NE |
| Sweden | <5 / 596.5 | 19 / 12040.3 | -8.8 (-70.6 to 53.0) | 26.4% (-141.5% to 100%) |
| BNT1BNT2MOD3 vs BNT1BNT2BNT3 | | | | |
| Denmark | 0 / 70.4 | 936 / 306929.8 | NE | NE |
| Finland | 23 / 41338.8 | 98 / 137285.6 | 9.8 (-28.2 to 47.9) | -47.6% (-236.3% to 100%) |
| Norway | 61 / 37625.3 | 533 / 166054.7 | -39.0 (-64.4 to -13.7) | 34.3% (13.5% to 55.1%) |
| Sweden | 95 / 64907.4 | 709 / 189673.1 | -21.8 (-32.7 to -10.9) | 34.0% (18.1% to 49.8%) |
| MOD1MOD2BNT3 vs BNT1BNT2BNT3 | | | | |
| Denmark | 0 / 111.2 | 1211 / 325568.4 | NE | NE |
| Finland | 6 / 6331.2 | 147 / 155538.0 | 27.6 (-40.6 to 95.8) | -81.0% (-284.2% to 100%) |
| Norway | 28 / 13595.4 | 645 / 192142.1 | -37.1 (-65.4 to -8.9) | 38.9% (12.7% to 65.1%) |
| Sweden | 58 / 16586.0 | 785 / 205396.6 | -11.1 (-33.4 to 11.3) | 12.0% (-12.0% to 36.1%) |
| BNT1MOD2MOD3 vs BNT1BNT2BNT3 | | | | |
| Denmark | <5 / 6.8 | 1169 / 321102.9 | 2339.6 (-2498.9 to 7178.0) | NE |
| Finland | 0 / 801.5 | 38 / 89852.8 | NE | NE |
| Norway | 12 / 8157.9 | 91 / 47676.2 | -0.7 (-89.8 to 88.4) | 0.7% (-92.8% to 94.2%) |
| Sweden | 0 / 48.9 | 534 / 179785.9 | NE | NE |
| MOD1BNT2BNT3 vs BNT1BNT2BNT3 | | | | |
| Denmark | 0 / 11.7 | 1220 / 328308.9 | NE | NE |
| Finland | 0 / 445.0 | 117 / 147816.9 | NE | NE |
| Norway | 0 / 964.1 | 99 / 56075.8 | NE | NE |
| Sweden | 0 / 121.9 | 731 / 203560.9 | NE | NE |
| BNT1MOD2BNT3 vs BNT1BNT2BNT3 | | | | |
| Denmark | 0 / 2.5 | 1062 / 315280.9 | NE | NE |

| | Studied schedule | Comparison schedule | Measures of association | |
|---|---|---|---|---|
| | Events/PYRS | Events/PYRS | RD per 100 000 individuals | CVE (95% CI) |
| Finland | 0 / 759.4 | 37 / 101939.4 | NE | NE |
| Norway | 21 / 12837.2 | 69 / 44793.7 | 45.0 (-66.3 to 156.4) | -92.4% (-336.5% to 100%) |
| Sweden | 0 / 143.7 | 161 / 123056.9 | NE | NE |
| MOD1BNT2MOD3 vs BNT1BNT2BNT3 | | | | |
| Denmark | 0 / 1.6 | 1157 / 316793.1 | NE | NE |
| Finland | 0 / 180.0 | 41 / 92590.3 | NE | NE |
| Norway | <5 / 632.9 | 100 / 50967.4 | -70.7 (-146.6 to 5.2) | 74.5% (22.1% to 100%) |
| Sweden | 0 / 27.7 | 655 / 187889.6 | NE | NE |

CI denotes confidence interval, CVE comparative vaccine effectiveness, ICU intensive care unit, NE not estimated, PYRS person-years, and RD risk difference. Cell counts lower than 5 could not be reported due to national regulations on data privacy protection.

**Supplementary Table 17. Association between COVID-19 intensive care unit admission and death and heterologous booster schedules as compared with homologous booster schedules.**

| | Studied schedule | Comparison schedule | Measures of association | |
|---|---|---|---|---|
| | Events/PYRS | Events/PYRS | RD (95% CI) per 100 000 individuals | CVE (95% CI) |
| AZD1AZD2BNT3 vs BNT1BNT2BNT3 | | | | |
| ICU admission | | | | |
| Denmark | 0 / 114.4 | 39 / 227596.7 | NE | NE |
| Finland | <5 / 15491.8 | <5 / 48050.2 | 0.5 (-3.4 to 4.4) | -44.1% (-416.2% to 100%) |
| Norway | 0 / 92.4 | 55 / 162943.1 | NE | NE |
| Sweden | 0 / 49123.0 | 0 / 155578.0 | NE | NE |
| Death | | | | |
| Denmark | 0 / 120.2 | 46 / 241932.7 | NE | NE |
| Finland | 0 / 15519.4 | 24 / 48115.7 | NE | NE |
| Norway | 0 / 93.3 | 53 / 164409.4 | NE | NE |
| Sweden | 21 / 49637.8 | 74 / 158389.7 | -3.8 (-8.6 to 1.1) | 32.3% (-3.8% to 68.4%) |
| AZD1AZD2MOD3 vs MOD1MOD2MOD3 | | | | |
| ICU admission | | | | |
| Denmark | 0 / 8.7 | <5 / 27091.9 | NE | NE |
| Finland | 0 / 5671.0 | 0 / 4735.3 | NE | NE |
| Norway | 0 / 29.8 | <5 / 16741.0 | NE | NE |
| Sweden | 0 / 16377.1 | 0 / 12723.9 | NE | NE |
| Death | | | | |
| Denmark | 0 / 8.9 | 6 / 29320.2 | NE | NE |
| Finland | <5 / 5675.5 | 0 / 4739.4 | NE | NE |
| Norway | 0 / 30.2 | <5 / 17033.3 | NE | NE |
| Sweden | <5 / 16535.8 | <5 / 12967.6 | 5.6 (-3.8 to 15.0) | NE |
| AZD1BNT2BNT3 vs BNT1BNT2BNT3 | | | | |
| ICU admission | | | | |
| Denmark | 0 / 8112.1 | 23 / 173408.0 | NE | NE |
| Finland | <5 / 14176.0 | <5 / 91389.0 | 5.3 (-3.1 to 13.7) | NE |
| Norway | <5 / 15017.0 | 24 / 96254.8 | -1.3 (-7.2 to 4.7) | 27.4% (-81.7% to 100%) |
| Sweden | 0 / 7316.0 | 0 / 148827.9 | NE | NE |
| Death | | | | |
| Denmark | 0 / 8712.8 | 25 / 185154.5 | NE | NE |
| Finland | 7 / 14283.9 | 13 / 91958.6 | -69.3 (-179.1 to 40.6) | 85.7% (63.4% to 100%) |
| Norway | <5 / 15252.7 | 15 / 97490.3 | -0.6 (-6.2 to 5.0) | 21.2% (-165.3% to 100%) |
| Sweden | <5 / 7717.6 | 32 / 152322.9 | -0.1 (-4.8 to 4.7) | 3.1% (-211.2% to 100%) |
| AZD1MOD2MOD3 vs MOD1MOD2MOD3 | | | | |
| ICU admission | | | | |
| Denmark | 0 / 4662.1 | <5 / 21890.1 | NE | NE |
| Finland | 0 / 2554.0 | 0 / 11218.9 | NE | NE |
| Norway | 0 / 153.1 | 0 / 12744.2 | NE | NE |
| Sweden | 0 / 596.5 | 0 / 12041.1 | NE | NE |
| Death | | | | |
| Denmark | 0 / 4952.2 | <5 / 23537.1 | NE | NE |
| Finland | <5 / 2568.1 | 0 / 11261.1 | NE | NE |
| Norway | 0 / 155.9 | 0 / 13021.4 | NE | NE |
| Sweden | 0 / 627.2 | 0 / 12341.2 | NE | NE |
| BNT1BNT2MOD3 vs BNT1BNT2BNT3 | | | | |

| | Studied schedule | Comparison schedule | Measures of association | |
|---|---|---|---|---|
| | **Events/PYRS** | **Events/PYRS** | **RD (95% CI) per 100 000 individuals** | **CVE (95% CI)** |
| ICU admission | | | | |
| Denmark | 0 / 70.4 | 70 / 306955.7 | NE | NE |
| Finland | 6 / 41339.1 | <5 / 137288.0 | -2.9 (-10.8 to 4.9) | 63.6% (-3.8% to 100%) |
| Norway | 7 / 37625.3 | 56 / 166055.1 | -3.5 (-12.2 to 5.2) | 30.0% (-41.2% to 100%) |
| Sweden | 0 / 64911.7 | 0 / 189697.0 | NE | NE |
| Death | | | | |
| Denmark | 0 / 72.2 | 127 / 322585.7 | NE | NE |
| Finland | 0 / 41447.1 | 36 / 137913.0 | NE | NE |
| Norway | 5 / 38171.3 | 64 / 167319.7 | -3.7 (-9.5 to 2.2) | 43.0% (-20.5% to 100%) |
| Sweden | 17 / 66396.5 | 148 / 193099.6 | -3.1 (-9.0 to 2.8) | 23.0% (-19.0% to 65.1%) |
| MOD1MOD2BNT3 vs BNT1BNT2BNT3 | | | | |
| ICU admission | | | | |
| Denmark | 0 / 111.2 | 82 / 325602.1 | NE | NE |
| Finland | 0 / 6331.3 | <5 / 155541.6 | NE | NE |
| Norway | <5 / 13595.5 | 66 / 192142.6 | -0.9 (-12.4 to 10.5) | 9.6% (-107.2% to 100%) |
| Sweden | 0 / 16588.0 | 0 / 205422.8 | NE | NE |
| Death | | | | |
| Denmark | 0 / 117.7 | 228 / 341336.0 | NE | NE |
| Finland | <5 / 6351.0 | 90 / 156214.9 | 0.4 (-29.7 to 30.5) | -1.8% (-131.6% to 100%) |
| Norway | 5 / 13848.8 | 100 / 193644.9 | -5.5 (-13.1 to 2.2) | 44.0% (-9.8% to 97.9%) |
| Sweden | 10 / 16922.6 | 173 / 209288.8 | -7.6 (-16.9 to 1.7) | 35.4% (-6.3% to 77.2%) |
| BNT1MOD2MOD3 vs BNT1BNT2BNT3 | | | | |
| ICU admission | | | | |
| Denmark | 0 / 6.8 | 79 / 321135.3 | NE | NE |
| Finland | 0 / 801.5 | <5 / 89853.7 | NE | NE |
| Norway | <5 / 8157.9 | 5 / 47676.5 | -4.4 (-13.3 to 4.5) | 76.3% (18.0% to 100%) |
| Sweden | 0 / 48.9 | 0 / 179804.6 | NE | NE |
| Death | | | | |
| Denmark | 0 / 6.9 | 216 / 336846.8 | NE | NE |
| Finland | 0 / 804.0 | 7 / 90293.6 | NE | NE |
| Norway | 0 / 8337.4 | <5 / 48394.0 | NE | NE |
| Sweden | 0 / 50.2 | 91 / 183222.1 | NE | NE |
| MOD1BNT2BNT3 vs BNT1BNT2BNT3 | | | | |
| ICU admission | | | | |
| Denmark | 0 / 11.7 | 83 / 328342.9 | NE | NE |
| Finland | 0 / 445.0 | <5 / 147819.8 | NE | NE |
| Norway | 0 / 964.1 | 7 / 56076.1 | NE | NE |
| Sweden | 0 / 121.9 | 0 / 203585.6 | NE | NE |
| Death | | | | |
| Denmark | 0 / 12.0 | 228 / 344332.5 | NE | NE |
| Finland | 0 / 446.9 | 58 / 148474.0 | NE | NE |
| Norway | 0 / 995.0 | 5 / 57016.5 | NE | NE |
| Sweden | 0 / 124.7 | 147 / 206641.2 | NE | NE |
| BNT1MOD2BNT3 vs BNT1BNT2BNT3 | | | | |
| ICU admission | | | | |
| Denmark | 0 / 2.5 | 73 / 315310.5 | NE | NE |
| Finland | 0 / 759.4 | <5 / 101940.3 | NE | NE |

| | Studied schedule | Comparison schedule | Measures of association | |
|---|---|---|---|---|
| | Events/PYRS | Events/PYRS | RD (95% CI) per 100 000 individuals | CVE (95% CI) |
| Norway | <5 / 12837.3 | 5 / 44794.0 | -10.9 (-30.9 to 9.2) | 91.3% (69.0% to 100%) |
| Sweden | 0 / 143.7 | 0 / 123063.7 | NE | NE |
| Death | | | | |
| Denmark | 0 / 2.7 | 168 / 331071.9 | NE | NE |
| Finland | 0 / 762.7 | 6 / 102457.5 | NE | NE |
| Norway | <5 / 13113.8 | <5 / 45628.5 | 0.1 (-4.7 to 4.9) | -6.6% (-248.1% to 100%) |
| Sweden | 0 / 147.7 | 20 / 123113.2 | NE | NE |
| MOD1BNT2MOD3 vs BNT1BNT2BNT3 | | | | |
| ICU admission | | | | |
| Denmark | 0 / 1.6 | 78 / 316825.1 | NE | NE |
| Finland | 0 / 180.0 | <5 / 92591.3 | NE | NE |
| Norway | 0 / 632.9 | 7 / 50967.8 | NE | NE |
| Sweden | 0 / 27.7 | 0 / 187911.9 | NE | NE |
| Death | | | | |
| Denmark | 0 / 1.6 | 195 / 331931.8 | NE | NE |
| Finland | 0 / 180.5 | 9 / 93032.0 | NE | NE |
| Norway | 0 / 653.7 | 6 / 51749.2 | NE | NE |
| Sweden | 0 / 28.2 | 119 / 188932.9 | NE | NE |

CI denotes confidence interval, CVE comparative vaccine effectiveness, ICU intensive care unit, NE not estimated, PYRS person-years, and RD risk difference. Cell counts lower than 5 could not be reported due to national regulations on data privacy protection.

**Supplementary Figure 20. Cumulative incidence curves of documented SARS-CoV-2 infection and severe COVID-19 outcomes comparing homologous MOD and BNT booster schedules in each country.**

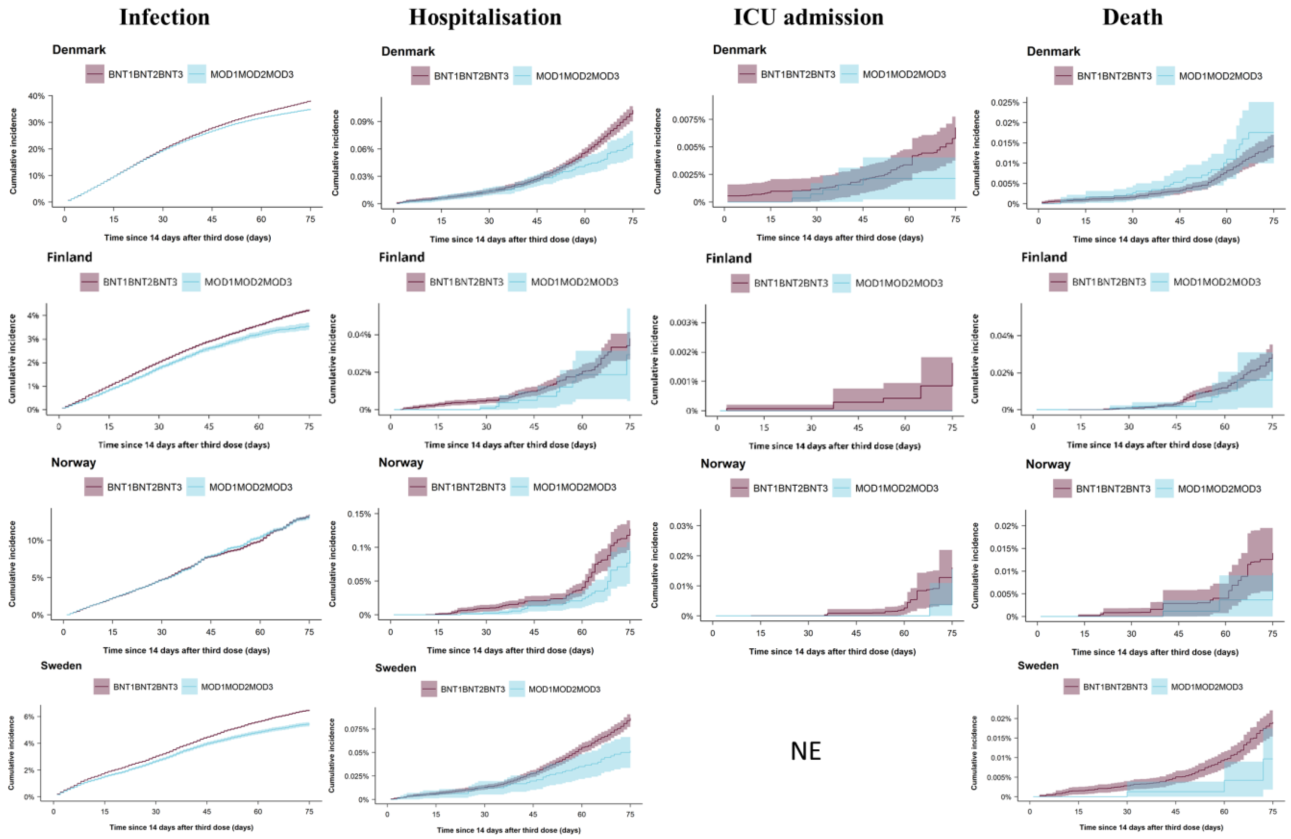

Rows are countries (Denmark, Finland, Norway, and Sweden) and columns are COVID-19 outcome of interest. NE denotes not estimated.

**Supplementary Table 18. Associated risk of COVID-19 outcomes comparing homologous BNT and MOD booster schedules.**

| | Studied schedule | Comparison schedule | Measures of association | |
|---|---|---|---|---|
| | Events/PYRS | Events/PYRS | RD (95% CI) per 100 or per 100 000 individuals[a] | CVE (95% CI) |
| MOD1MOD2MOD3 vs. BNT1BNT2BNT3 | | | | |
| Documented infection | | | | |
| Denmark | 87100 / 37241.8 | 574850 / 294734.5 | -3.1 (-3.4 to -2.9) | 8.2% (7.6% to 8.9%) |
| Finland | 3338 / 16225.1 | 36187 / 151925.9 | -0.7 (-0.8 to -0.5) | 16.0% (12.2% to 19.9%) |
| Norway | 10750 / 16259.1 | 61093 / 160717.6 | -0.1 (-0.4 to 0.2) | 0.8% (-1.7% to 3.3%) |
| Sweden | 4615 / 16175.2 | 67320 / 189016.9 | -1.0 (-1.2 to -0.9) | 16.0% (13.5% to 18.5%) |
| Hospitalisation | | | | |
| Denmark | 102 / 40560.2 | 1114 / 315452.1 | -35.6 (-53.7 to -17.6) | 34.8% (18.4% to 51.1%) |
| Finland | 11 / 16319.6 | 152 / 153020.4 | -8.6 (-34.7 to 17.5) | 22.6% (-44.8% to 90.0%) |
| Norway | 30 / 16620.2 | 535 / 163463.0 | -32.9 (-76.6 to 10.9) | 25.8% (-6.3% to 57.8%) |
| Sweden | 37 / 16416.3 | 759 / 192041.8 | -34.7 (-52.9 to -16.5) | 40.1% (20.1% to 60.1%) |
| ICU admission | | | | |
| Denmark | 5 / 40563.3 | 75 / 315487.9 | -4.6 (-7.7 to -1.5) | 68.4% (37.9% to 98.8%) |
| Finland | 0 / 16319.8 | <5 / 153024.1 | NE | NE |
| Norway | <5 / 16620.2 | 57 / 163463.5 | -0.3 (-21.6 to 20.9) | 2.1% (-129.7% to 100%) |
| Sweden | 0 / 16417.7 | 0 / 192066.8 | NE | NE |
| Death | | | | |
| Denmark | 27 / 43056.1 | 188 / 330993.5 | 3.3 (-4.8 to 11.3) | -23.0% (-81.4% to 35.4%) |
| Finland | 7 / 16367.5 | 94 / 153679.4 | -2.7 (-31.2 to 25.8) | 8.9% (-84.5% to 100%) |
| Norway | <5 / 16907.6 | 66 / 164766.7 | -4.4 (-19.2 to 10.5) | 31.4% (-67.3% to 100%) |
| Sweden | 6 / 16730.0 | 166 / 195475.0 | -9.4 (-17.8 to -0.9) | 49.2% (7.2% to 91.2%) |

CI denotes confidence interval, CVE comparative vaccine effectiveness, ICU intensive care unit, NE not estimated, PYRS person-years, and RD risk difference. Cell counts lower than 5 could not be reported due to national regulations on data privacy protection. [a]Risk differences for documented SARS-CoV-2 infection are reported per 100 individuals while risk differences for the severe COVID-19 outcomes of hospitalization, intensive care unit admission, and death are reported per 100 000 individuals.

**Supplementary Figure 21. Cumulative incidence curves of documented SARS-CoV-2 infection comparing heterologous booster- with matched primary schedules in each country according to history of previous SARS-CoV-2 infection.**

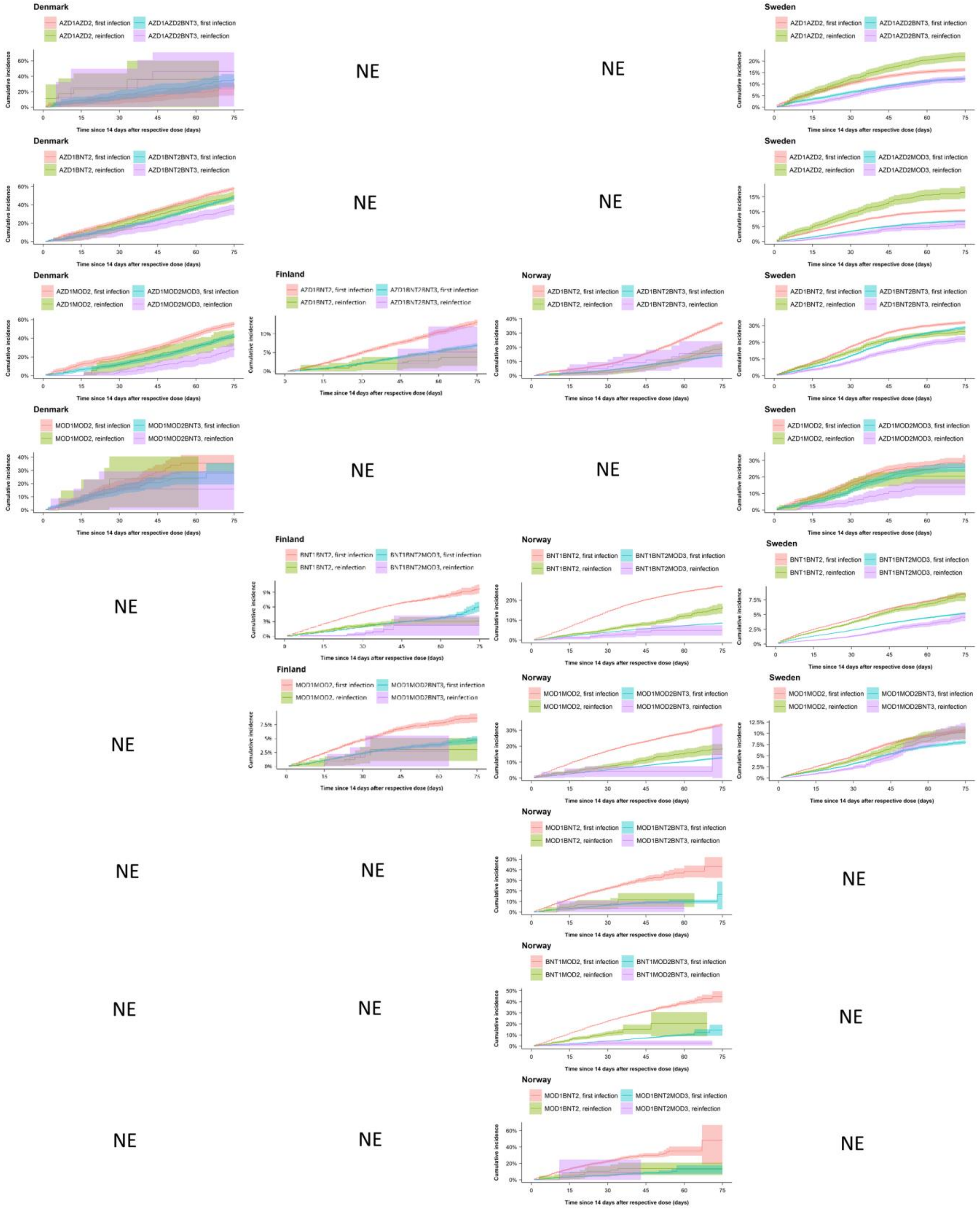

Columns are countries (Denmark, Finland, Norway, and Sweden) and rows are schedule comparisons. NE denotes not estimated. Individuals with a positive PCR test for SARS-CoV-2 within 12 weeks prior to index date were not included.

**Supplementary Figure 22. Cumulative incidence curves of documented SARS-CoV-2 infection comparing heterologous- and homologous booster schedules in each country according to history of previous SARS-CoV-2 infection.**

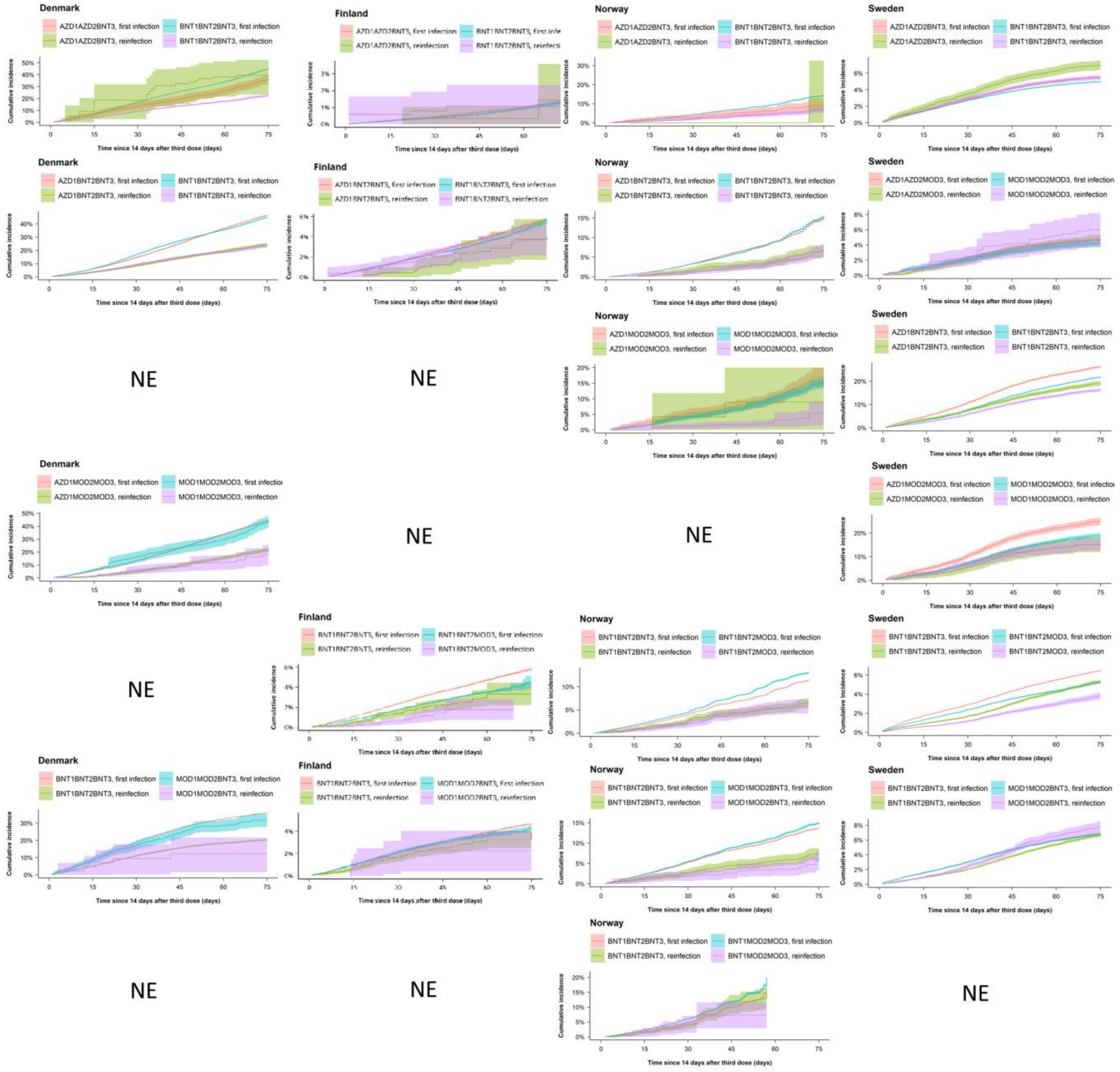

Columns are countries (Denmark, Finland, Norway, and Sweden) and rows are schedule comparisons. NE denotes not estimated. Individuals with a positive PCR test for SARS-CoV-2 within 12 weeks prior to index date were not included.

**Supplementary Figure 23. Cumulative incidence curves of documented SARS-CoV-2 infection comparing homologous booster schedules with matched primary schedules in each country according to history of previous SARS-CoV-2 infection.**

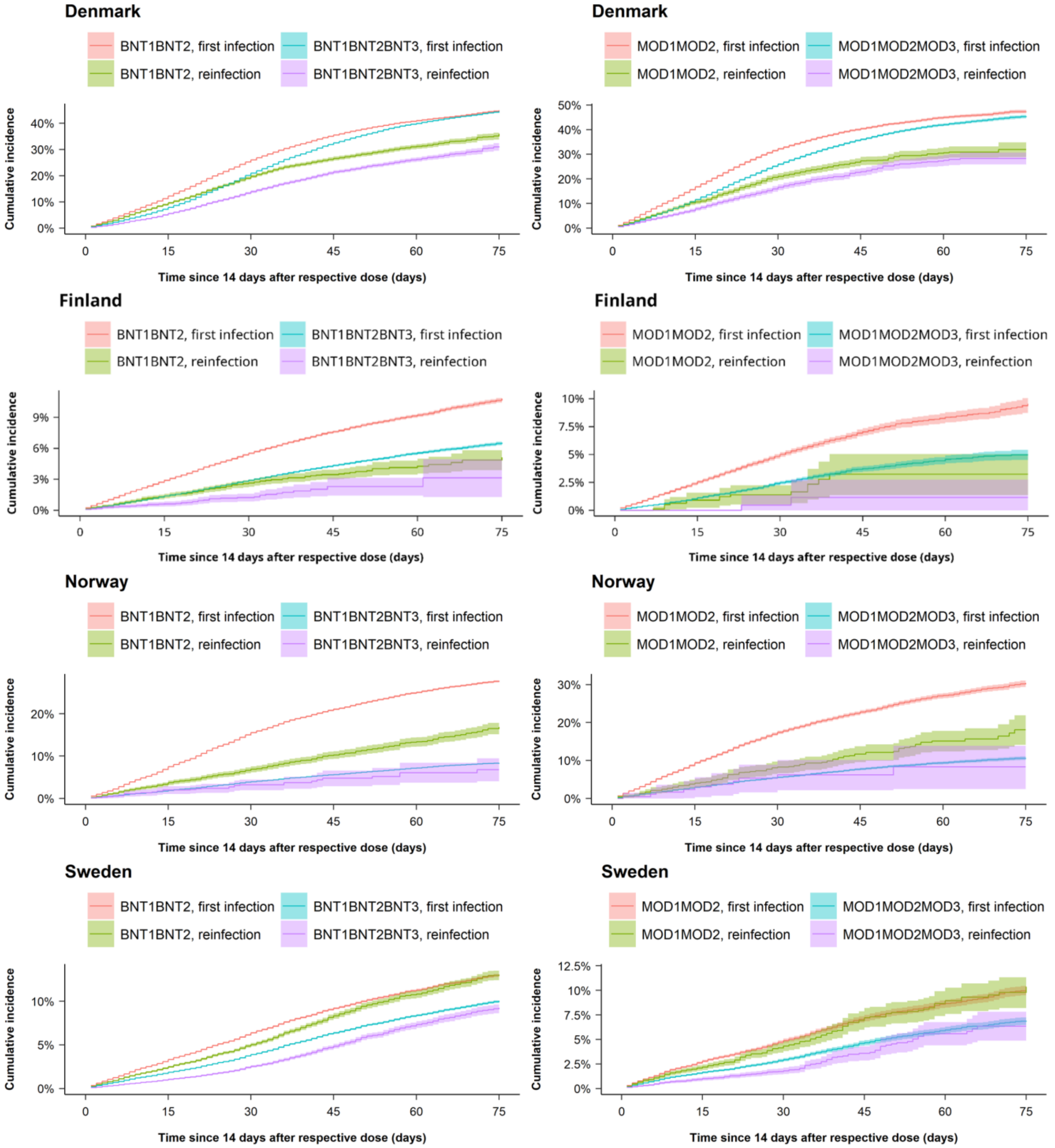

Rows are countries (Denmark, Finland, Norway, and Sweden) and columns are schedule comparisons. Individuals with a positive PCR test for SARS-CoV-2 within 12 weeks prior to index date were not included.

**Supplementary Figure 24. Cumulative incidence curves of documented SARS-CoV-2 infection comparing homologous MOD and BNT booster schedules in each country according to history of previous SARS-CoV-2 infection.**

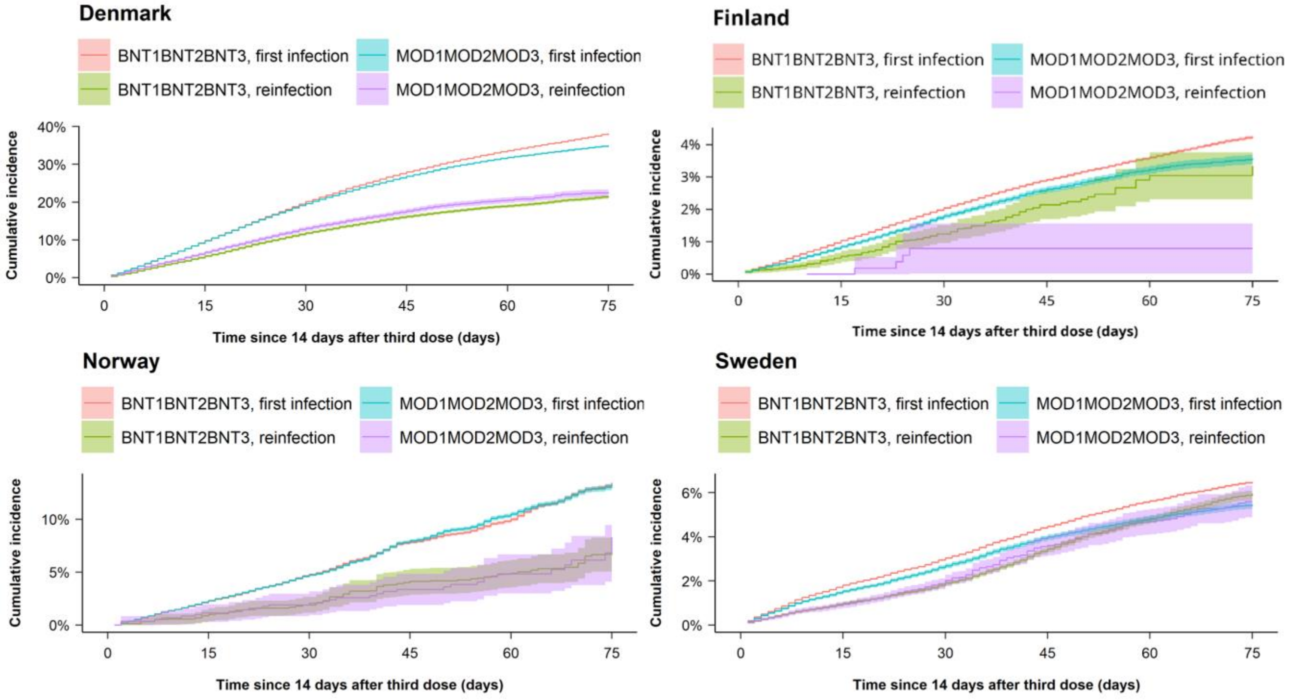

Individuals with a positive PCR test for SARS-CoV-2 within 12 weeks prior to index date were not included.

**Supplementary Table 19. Association between documented SARS-CoV-2 infection and booster schedules as compared with matched primary schedules in each country according to history of previous SARS-CoV-2 infection.**

| | Previously infected | Studied schedule | Comparison schedule | Measures of association | |
|---|---|---|---|---|---|
| | | Events/PYRS | Events/PYRS | RD per 100 individuals | CVE (95% CI) |
| AZD1AZD2BNT3 vs AZD1AZD2 | | | | | |
| Denmark | No | 46 / 22.1 | 28 / 19.8 | 7.0 (-14.9 to 28.9) | -23.5% (-106.4% to 59.4%) |
| Denmark | Yes | <5 / 1.2 | <5 / 1.2 | NE | NE |
| Finland | No | 132 / 2181.0 | 406 / 2077.6 | -2.3 (-2.9 to -1.7) | 64.6% (54.8% to 74.4%) |
| Finland | Yes | 0 / 6.1 | 0 / 23.0 | NE | NE |
| Norway | No | 17 / 59.8 | 20 / 56.8 | -1.0 (-4.5 to 2.6) | 17.0% (-39.7% to 73.7%) |
| Norway | Yes | 0 / 0.6 | <5 / 1.5 | NE | NE |
| Sweden | No | 3166 / 4953.6 | 3712 / 4557.9 | -3.9 (-5.0 to -2.8) | 24.0% (18.0% to 30.0%) |
| Sweden | Yes | 305 / 417.1 | 546 / 417.6 | -9.4 (-12.4 to -6.5) | 43.1% (33.1% to 53.0%) |
| AZD1AZD2MOD3 vs AZD1AZD2 | | | | | |
| Denmark | No | <5 / 4.7 | <5 / 4.4 | 2.8 (-17.1 to 22.7) | -25.1% (-225.5% to 100%) |
| Denmark | Yes | NE | NE | NE | NE |
| Finland | No | 47 / 1472.4 | 239 / 1424.7 | -2.5 (-3.5 to -1.4) | 81.1% (71.7% to 90.6%) |
| Finland | Yes | 0 / 3.1 | 0 / 19.4 | NE | NE |
| Norway | No | 10 / 21.5 | <5 / 21.4 | 3.9 (-1.7 to 9.4) | NE |
| Norway | Yes | NE | NE | NE | NE |
| Sweden | No | 1574 / 4043.9 | 2298 / 3781.4 | -3.7 (-4.3 to -3.0) | 35.0% (30.3% to 39.6%) |
| Sweden | Yes | 101 / 318.8 | 276 / 270.9 | -10.7 (-13.6 to -7.9) | 65.1% (55.0% to 75.2%) |
| AZD1BNT2BNT3 vs AZD1BNT2 | | | | | |
| Denmark | No | 1712 / 497.2 | 1372 / 315.9 | -9.9 (-15.9 to -3.9) | 17.2% (8.0% to 26.4%) |
| Denmark | Yes | 115 / 50.6 | 173 / 47.6 | -14.4 (-28.3 to -0.4) | 28.6% (5.4% to 51.8%) |
| Finland | No | 570 / 1661.7 | 1010 / 1484.8 | -6.3 (-7.5 to -5.1) | 47.3% (41.0% to 53.6%) |
| Finland | Yes | <5 / 15.3 | 10 / 49.3 | 1.4 (-6.5 to 9.2) | -36.3% (-253.0% to 100%) |
| Norway | No | 1800 / 2418.9 | 3249 / 1692.4 | -22.9 (-24.9 to -20.9) | 61.1% (58.2% to 64.0%) |
| Norway | Yes | 10 / 10.8 | 90 / 89.4 | -3.5 (-15.3 to 8.4) | 18.2% (-42.8% to 79.1%) |
| Sweden | No | 3689 / 2081.4 | 4007 / 1839.2 | -3.0 (-4.9 to -1.1) | 9.4% (3.8% to 15.0%) |
| Sweden | Yes | 538 / 422.4 | 714 / 412.3 | -4.3 (-7.9 to -0.7) | 16.3% (3.8% to 28.9%) |
| AZD1MOD2MOD3 vs AZD1MOD2 | | | | | |
| Denmark | No | 861 / 290.8 | 844 / 207.4 | -13.1 (-20.6 to -5.7) | 23.4% (12.2% to 34.6%) |
| Denmark | Yes | 41 / 21.1 | 51 / 17.8 | -12.8 (-31.4 to 5.7) | 30.3% (-5.3% to 65.8%) |
| Finland | No | 110 / 366.8 | 233 / 339.2 | -8.7 (-11.5 to -5.8) | 59.6% (48.4% to 70.8%) |
| Finland | Yes | 0 / 2.1 | <5 / 7.2 | NE | NE |
| Norway | No | 31 / 43.1 | 60 / 28.5 | -23.0 (-37.3 to -8.6) | 63.9% (44.1% to 83.7%) |
| Norway | Yes | 0 / 0.9 | <5 / 2.3 | NE | NE |
| Sweden | No | 317 / 190.9 | 309 / 156.2 | -4.0 (-10.3 to 2.3) | 13.4% (-6.0% to 32.8%) |
| Sweden | Yes | 30 / 35.4 | 65 / 42.5 | -6.6 (-14.7 to 1.5) | 32.1% (-1.3% to 65.6%) |
| BNT1BNT2MOD3 vs BNT1BNT2 | | | | | |
| Denmark | No | 32 / 33.4 | 58 / 22.0 | -16.6 (-33.2 to 0.0) | 47.5% (15.5% to 79.6%) |
| Denmark | Yes | 0 / 1.5 | <5 / 1.2 | NE | NE |
| Finland | No | 4375 / 18778.1 | 10951 / 17954.2 | -3.8 (-5.4 to -2.2) | 38.7% (25.4% to 52.0%) |
| Finland | Yes | 9 / 127.5 | 134 / 501.9 | -0.9 (-3.1 to 1.4) | 28.2% (-41.5% to 97.8%) |
| Norway | No | 9117 / 19000.7 | 25026 / 14011.2 | -18.5 (-19.3 to -17.8) | 68.3% (67.0% to 69.7%) |
| Norway | Yes | 19 / 65.4 | 344 / 470.5 | -12.0 (-16.5 to -7.6) | 71.2% (54.1% to 88.3%) |
| Sweden | No | 8719 / 29333.9 | 14641 / 27481.3 | -3.4 (-3.7 to -3.1) | 39.2% (36.6% to 41.8%) |
| Sweden | Yes | 719 / 4113.5 | 1851 / 3899.9 | -3.9 (-5.1 to -2.7) | 46.3% (35.6% to 56.9%) |
| MOD1MOD2BNT3 vs MOD1MOD2 | | | | | |
| Denmark | No | 88 / 46.9 | 94 / 37.6 | -7.5 (-22.7 to 7.6) | 21.3% (-17.7% to 60.3%) |
| Denmark | Yes | <5 / 3.0 | 5 / 2.2 | NE | NE |
| Finland | No | 755 / 2727.1 | 1534 / 2641.0 | -3.9 (-5.1 to -2.8) | 45.3% (35.3% to 55.2%) |
| Finland | Yes | <5 / 19.6 | 10 / 41.8 | NE | NE |
| Norway | No | 3328 / 5069.1 | 8125 / 3694.8 | -20.8 (-22.4 to -19.2) | 62.0% (59.4% to 64.5%) |
| Norway | Yes | 7 / 13.8 | 131 / 133.1 | -4.0 (-27.8 to 19.7) | 21.4% (-104.1% to 100%) |
| Sweden | No | 2340 / 5711.8 | 3324 / 5394.1 | -2.7 (-3.4 to -2.1) | 25.4% (19.9% to 30.8%) |
| Sweden | Yes | 287 / 861.5 | 431 / 841.0 | 0.4 (-2.2 to 3.0) | -4.0% (-30.1% to 22.1%) |

| | | | | | |
|---|---|---|---|---|---|
| BNT1MOD2MOD3 vs BNT1MOD2 | | | | | |
| Denmark | No | NE | NE | NE | NE |
| Denmark | Yes | NE | NE | NE | NE |
| Finland | No | 131 / 384.2 | 258 / 367.1 | -14.2 (-40.8 to 12.5) | 66.9% (22.3% to 100%) |
| Finland | Yes | NE | NE | NE | NE |
| Norway | No | 2810 / 4340.5 | 9306 / 3450.0 | -29.9 (-45.4 to -14.5) | 70.8% (58.4% to 83.3%) |
| Norway | Yes | NE | NE | NE | NE |
| Sweden | No | 7 / 25.8 | 6 / 25.6 | -2.0 (-6.1 to 2.1) | 58.3% (0.7% to 100%) |
| Sweden | Yes | <5 / 5.2 | <5 / 5.1 | NE | NE |
| MOD1BNT2BNT3 vs MOD1BNT2 | | | | | |
| Denmark | No | 5 / 1.1 | <5 / 1.3 | 26.6 (-56.5 to 100) | -92.2% (-458.0% to 100%) |
| Denmark | Yes | NE | NE | NE | NE |
| Finland | No | 51 / 153.6 | 112 / 147.6 | -3.6 (-7.0 to -0.1) | 39.4% (7.7% to 71.1%) |
| Finland | Yes | NE | NE | NE | NE |
| Norway | No | 427 / 526.4 | 1222 / 389.5 | -26.3 (-49.7 to -2.9) | 61.0% (21.1% to 100%) |
| Norway | Yes | NE | NE | NE | NE |
| Sweden | No | 10 / 60.2 | 15 / 58.3 | 2.9 (-3.1 to 8.9) | NE |
| Sweden | Yes | <5 / 13.3 | <5 / 13.5 | NE | NE |
| BNT1MOD2BNT3 vs BNT1MOD2 | | | | | |
| Denmark | No | NE | NE | NE | NE |
| Denmark | Yes | NE | NE | NE | NE |
| Finland | No | 169 / 425.6 | 283 / 409.5 | -8.4 (-21.0 to 4.1) | 54.8% (14.7% to 95.0%) |
| Finland | Yes | NE | NE | NE | NE |
| Norway | No | 4622 / 8124.7 | 20453 / 6719.6 | -30.3 (-41.3 to -19.3) | 67.8% (53.1% to 82.4%) |
| Norway | Yes | NE | NE | NE | NE |
| Sweden | No | 13 / 90.7 | 18 / 89.7 | -5.5 (-16.8 to 5.9) | 57.5% (-11.5% to 100%) |
| Sweden | Yes | <5 / 19.5 | <5 / 18.9 | 6.4 (-44.6 to 57.5) | -47.5% (-478.7% to 100%) |
| MOD1BNT2MOD3 vs MOD1BNT2 | | | | | |
| Denmark | No | NE | NE | NE | NE |
| Denmark | Yes | NE | NE | NE | NE |
| Finland | No | 18 / 95.6 | 70 / 91.8 | -7.0 (-16.9 to 2.9) | 57.5% (1.0% to 100%) |
| Finland | Yes | NE | NE | NE | NE |
| Norway | No | 228 / 309.3 | 668 / 209.8 | -35.6 (-80.7 to 9.4) | 74.0% (46.8% to 100%) |
| Norway | Yes | <5 / 1.1 | 11 / 8.1 | NE | NE |
| Sweden | No | 6 / 15.3 | <5 / 14.6 | 12.7 (-13.7 to 39.0) | NE |
| Sweden | Yes | NE | NE | NE | NE |
| BNT1BNT2BNT3 vs BNT1BNT2 | | | | | |
| Denmark | No | 96037 / 33872.9 | 76560 / 23389.1 | -0.4 (-1.2 to 0.4) | 0.8% (-1.0% to 2.6%) |
| Denmark | Yes | 4185 / 2357.5 | 4608 / 1914.3 | -4.8 (-7.8 to -1.8) | 13.4% (5.5% to 21.2%) |
| Finland | No | 12673 / 36307.5 | 22812 / 34411.7 | -4.3 (-4.6 to -4.0) | 39.5% (37.2% to 41.8%) |
| Finland | Yes | 53 / 316.6 | 274 / 888.7 | -2.0 (-4.2 to 0.2) | 38.7% (-0.9% to 78.4%) |
| Norway | No | 17285 / 37754.6 | 53369 / 29243.6 | -19.4 (-19.8 to -19.0) | 69.8% (69.2% to 70.5%) |
| Norway | Yes | 34 / 92.3 | 614 / 698.9 | -10.1 (-13.4 to -6.7) | 59.7% (41.9% to 77.5%) |
| Sweden | No | 22450 / 44969.3 | 31505 / 41626.6 | -3.0 (-3.3 to -2.7) | 23.0% (21.3% to 24.8%) |
| Sweden | Yes | 2265 / 6454.7 | 4116 / 6312.2 | -3.8 (-4.6 to -3.0) | 29.1% (23.6% to 34.6%) |
| MOD1MOD2MOD3 vs MOD1MOD2 | | | | | |
| Denmark | No | 23779 / 6992.2 | 21875 / 5198.9 | -2.0 (-3.9 to 0.0) | 4.2% (0.2% to 8.2%) |
| Denmark | Yes | 856 / 412.9 | 978 / 379.1 | -3.6 (-9.1 to 1.9) | 11.2% (-4.8% to 27.3%) |
| Finland | No | 1292 / 4456.6 | 2563 / 4293.3 | -4.6 (-5.5 to -3.7) | 47.9% (41.5% to 54.3%) |
| Finland | Yes | <5 / 29.1 | 15 / 68.2 | -2.1 (-4.6 to 0.4) | 64.4% (9.6% to 100%) |
| Norway | No | 3248 / 5031.9 | 7690 / 3614.8 | -19.8 (-21.1 to -18.4) | 65.1% (62.8% to 67.4%) |
| Norway | Yes | 10 / 17.4 | 133 / 134.2 | -9.8 (-17.6 to -1.9) | 53.9% (17.6% to 90.3%) |
| Sweden | No | 1955 / 5233.8 | 2955 / 4935.9 | -3.1 (-3.8 to -2.4) | 31.0% (25.5% to 36.6%) |
| Sweden | Yes | 202 / 765.5 | 405 / 735.1 | -4.0 (-6.5 to -1.5) | 38.7% (19.7% to 57.6%) |

CI denotes confidence interval, CVE comparative vaccine effectiveness, ICU intensive care unit, NE not estimated, PYRS person-years, and RD risk difference. Cell counts lower than 5 could not be reported due to national regulations on data privacy protection. Individuals with a positive PCR test for SARS-CoV-2 within 12 weeks prior to index date were not included.

**Supplementary Table 20. Association between documented SARS-CoV-2 infection and heterologous booster schedules as compared with homologous booster schedules in each country according to history of previous SARS-CoV-2 infection.**

| | Previously infected | Studied schedule Events/PYRS | Comparison schedule Events/PYRS | Measures of association RD per 100 individuals | CVE (95% CI) |
|---|---|---|---|---|---|
| AZD1AZD2BNT3 vs BNT1BNT2BNT3 | | | | | |
| Denmark | No | 229 / 106.7 | 517302 / 210481.9 | -9.1 (-12.9 to -5.3) | 20.1% (11.7% to 28.4%) |
| Denmark | Yes | 16 / 6.9 | 14403 / 13653.9 | 16.9 (2.4 to 31.4) | -74.3% (-138.4% to -10.3%) |
| Finland | No | 986 / 15457.7 | 2356 / 47969.2 | -0.1 (-0.3 to 0.1) | 4.1% (-9.9% to 18.1%) |
| Finland | Yes | <5 / 42.5 | 5 / 117.5 | 0.3 (-2.2 to 2.9) | -32.2% (-290.2% to 100%) |
| Norway | No | 41 / 92.0 | 68584 / 166272.9 | -5.4 (-8.4 to -2.5) | 37.5% (17.0% to 58.0%) |
| Norway | Yes | <5 / 3.3 | 217 / 916.3 | 5.3 (-17.6 to 28.3) | -74.7% (-397.1% to 100%) |
| Sweden | No | 12843 / 48515.0 | 58756 / 152423.5 | 0.5 (0.4 to 0.7) | -10.6% (-13.6% to -7.7%) |
| Sweden | Yes | 728 / 2089.5 | 3666 / 10389.5 | 1.5 (0.9 to 2.1) | -27.3% (-39.8% to -14.8%) |
| AZD1AZD2MOD3 vs MOD1MOD2MOD3 | | | | | |
| Denmark | No | 8 / 8.3 | 75144 / 24196.7 | -16.1 (-29.2 to -3.0) | 46.6% (8.9% to 84.4%) |
| Denmark | Yes | 0 / 0.3 | 2207 / 1332.7 | NE | NE |
| Finland | No | 176 / 5665.3 | 187 / 4729.5 | -0.3 (-0.6 to 0.0) | 29.5% (3.1% to 56.0%) |
| Finland | Yes | 0 / 12.1 | 0 / 10.5 | NE | NE |
| Norway | No | 17 / 29.3 | 10959 / 16423.4 | -3.2 (-8.1 to 1.8) | 23.9% (-12.9% to 60.8%) |
| Norway | Yes | 0 / 0.4 | 32 / 98.6 | NE | NE |
| Sweden | No | 3915 / 16218.7 | 3707 / 12528.6 | 0.5 (0.1 to 1.0) | -12.8% (-25.0% to -0.5%) |
| Sweden | Yes | 214 / 827.4 | 290 / 1074.1 | -1.3 (-3.6 to 1.0) | 21.2% (-9.8% to 52.2%) |
| AZD1BNT2BNT3 vs BNT1BNT2BNT3 | | | | | |
| Denmark | No | 23844 / 7275.3 | 414234 / 156233.3 | 1.5 (0.9 to 2.0) | -3.2% (-4.5% to -2.0%) |
| Denmark | Yes | 1453 / 1002.8 | 11728 / 11178.6 | 1.2 (-0.4 to 2.9) | -5.3% (-12.4% to 1.8%) |
| Finland | No | 4048 / 14038.0 | 30942 / 90440.2 | 0.2 (-0.1 to 0.5) | -3.5% (-9.1% to 2.0%) |
| Finland | Yes | 14 / 75.4 | 109 / 524.0 | -0.3 (-2.9 to 2.3) | 6.5% (-56.6% to 69.5%) |
| Norway | No | 10967 / 14713.4 | 54797 / 94782.4 | -0.4 (-0.8 to 0.0) | 2.4% (-0.1% to 4.9%) |
| Norway | Yes | 57 / 165.7 | 171 / 543.8 | -0.4 (-2.8 to 1.9) | 5.8% (-27.0% to 38.6%) |
| Sweden | No | 10752 / 6894.7 | 63322 / 145840.7 | 4.4 (3.9 to 5.0) | -20.2% (-22.8% to -17.6%) |
| Sweden | Yes | 1199 / 1118.0 | 4504 / 13415.1 | 2.8 (1.5 to 4.0) | -17.0% (-25.1% to -8.8%) |
| AZD1MOD2MOD3 vs MOD1MOD2MOD3 | | | | | |
| Denmark | No | 13053 / 4220.7 | 54891 / 18868.7 | 0.7 (-3.8 to 5.3) | -1.6% (-12.2% to 8.9%) |
| Denmark | Yes | 491 / 364.0 | 1552 / 1019.1 | 4.0 (-4.2 to 12.1) | -22.0% (-76.6% to 32.6%) |
| Finland | No | 625 / 2534.1 | 2871 / 11136.5 | 1.5 (0.8 to 2.1) | -38.9% (-60.4% to -17.3%) |
| Finland | Yes | <5 / 9.4 | <5 / 47.0 | 7.7 (-1.9 to 17.3) | NE |
| Norway | No | 119 / 149.8 | 10078 / 12447.5 | 1.4 (-2.2 to 4.9) | -8.7% (-31.9% to 14.5%) |
| Norway | Yes | <5 / 3.7 | 32 / 71.9 | 3.3 (-9.1 to 15.8) | -59.3% (-296.9% to 100%) |
| Sweden | No | 846 / 563.9 | 4155 / 11813.3 | 6.0 (4.1 to 7.9) | -31.6% (-42.8% to -20.4%) |
| Sweden | Yes | 83 / 92.3 | 363 / 1285.7 | -0.2 (-4.3 to 3.8) | 1.4% (-24.9% to 27.8%) |
| BNT1BNT2MOD3 vs BNT1BNT2BNT3 | | | | | |
| Denmark | No | 67 / 68.1 | 580653 / 285991.3 | -20.1 (-24.1 to -16.0) | 54.1% (43.2% to 65.0%) |
| Denmark | Yes | 0 / 3.6 | 15302 / 15970.6 | NE | NE |
| Finland | No | 9045 / 41087.3 | 35133 / 136227.2 | -1.4 (-2.1 to -0.7) | 23.6% (12.0% to 35.2%) |
| Finland | Yes | 20 / 210.0 | 135 / 704.4 | NE | NE |
| Norway | No | 22855 / 37121.6 | 61453 / 165992.3 | 1.7 (1.5 to 1.9) | -14.7% (-16.8% to -12.7%) |
| Norway | Yes | 61 / 208.0 | 177 / 886.3 | -1.0 (-2.8 to 0.9) | 14.7% (-11.4% to 40.8%) |
| Sweden | No | 17858 / 63874.2 | 66695 / 186660.1 | -1.2 (-1.3 to -1.1) | 18.9% (17.4% to 20.4%) |
| Sweden | Yes | 1071 / 6629.0 | 4182 / 13688.7 | -1.6 (-2.0 to -1.1) | 28.9% (21.6% to 36.1%) |
| MOD1MOD2BNT3 vs BNT1BNT2BNT3 | | | | | |
| Denmark | No | 215 / 103.1 | 586998 / 304529.9 | -4.4 (-8.3 to -0.6) | 12.2% (1.5% to 23.0%) |
| Denmark | Yes | 5 / 5.7 | 15323 / 16322.1 | -8.3 (-18.4 to 1.7) | 40.6% (-8.4% to 89.6%) |
| Finland | No | 1531 / 6288.5 | 37728 / 154405.4 | -0.4 (-0.7 to -0.1) | 8.7% (2.4% to 15.1%) |
| Finland | Yes | 6 / 30.6 | 154 / 782.0 | -1.7 (-4.0 to 0.7) | 42.7% (-8.7% to 94.2%) |
| Norway | No | 10124 / 13305.5 | 70655 / 190302.9 | 1.3 (1.0 to 1.7) | -9.5% (-12.1% to -7.0%) |
| Norway | Yes | 24 / 97.6 | 225 / 1042.5 | -1.3 (-4.3 to 1.7) | 17.0% (-21.6% to 55.7%) |
| Sweden | No | 5743 / 16308.3 | 74724 / 202017.4 | -0.2 (-0.3 to 0.0) | 2.4% (-0.2% to 5.0%) |
| Sweden | Yes | 517 / 1498.3 | 4980 / 15708.2 | 1.1 (0.3 to 1.9) | -15.8% (-28.0% to -3.7%) |

| | | | | | |
|---|---|---|---|---|---|
| BNT1MOD2MOD3 vs BNT1BNT2BNT3 | | | | | |
| Denmark | No | 10 / 6.4 | 573968 / 297012.1 | -5.5 (-17.9 to 6.9) | 19.1% (-23.8% to 62.0%) |
| Denmark | Yes | NE | 14779 / 15874.0 | NE | NE |
| Finland | No | 284 / 793.9 | 28741 / 89021.6 | -1.4 (-2.8 to 0.0) | 19.7% (0.7% to 38.8%) |
| Finland | Yes | 0 / 4.7 | 125 / 621.0 | NE | NE |
| Norway | No | 6995 / 7980.7 | 30755 / 47764.2 | 3.3 (2.2 to 4.3) | -19.4% (-25.7% to -13.1%) |
| Norway | Yes | 12 / 21.5 | 99 / 174.1 | -7.7 (-14.4 to -0.9) | 51.0% (17.3% to 84.7%) |
| Sweden | No | 12 / 48.2 | 66066 / 177791.1 | -3.9 (-5.6 to -2.2) | 60.0% (33.8% to 86.2%) |
| Sweden | Yes | <5 / 7.9 | 4219 / 13599.9 | 0.7 (-11.1 to 12.6) | -12.9% (-227.3% to 100%) |
| MOD1BNT2BNT3 vs BNT1BNT2BNT3 | | | | | |
| Denmark | No | 20 / 11.1 | 595378 / 306951.1 | 4.1 (-7.6 to 15.8) | -15.1% (-57.8% to 27.7%) |
| Denmark | Yes | 0 / 0.4 | 15753 / 16554.4 | NE | NE |
| Finland | No | 119 / 441.6 | 37126 / 146707.4 | 1.1 (-1.1 to 3.2) | -20.0% (-61.2% to 21.1%) |
| Finland | Yes | <5 / 2.4 | 151 / 777.4 | 12.0 (-9.2 to 33.3) | NE |
| Norway | No | 1064 / 930.8 | 38236 / 55002.6 | 6.2 (0.3 to 12.1) | -27.1% (-52.9% to -1.3%) |
| Norway | Yes | <5 / 4.8 | 134 / 218.4 | NE | NE |
| Sweden | No | 41 / 119.4 | 74256 / 200190.0 | 1.7 (-1.5 to 4.8) | -23.7% (-67.9% to 20.6%) |
| Sweden | Yes | <5 / 18.4 | 4971 / 15763.1 | 14.7 (-11.3 to 40.6) | NE |
| BNT1MOD2BNT3 vs BNT1BNT2BNT3 | | | | | |
| Denmark | No | <5 / 2.3 | 584116 / 294224.6 | -18.8 (-44.3 to 6.8) | 44.6% (-16.1% to 100%) |
| Denmark | Yes | NE | 15420 / 16039.5 | NE | NE |
| Finland | No | 305 / 751.1 | 31999 / 101003.0 | -2.3 (-3.4 to -1.2) | 28.8% (15.1% to 42.4%) |
| Finland | Yes | <5 / 5.9 | 135 / 662.6 | NE | NE |
| Norway | No | 10660 / 12518.7 | 32303 / 43862.4 | 1.4 (-0.1 to 2.9) | -7.8% (-16.0% to 0.4%) |
| Norway | Yes | 21 / 37.1 | 123 / 168.7 | 10.2 (-23.1 to 43.4) | -46.1% (-203.7% to 100%) |
| Sweden | No | 19 / 142.3 | 52426 / 123388.3 | -1.6 (-5.7 to 2.6) | 23.0% (-38.5% to 84.6%) |
| Sweden | Yes | <5 / 26.7 | 3999 / 13113.2 | -5.8 (-6.4 to -5.2) | 96.7% (90.3% to 100%) |
| MOD1BNT2MOD3 vs BNT1BNT2BNT3 | | | | | |
| Denmark | No | 0 / 1.6 | 564709 / 296644.2 | NE | NE |
| Denmark | Yes | 0 / 0.2 | 14232 / 15743.2 | NE | NE |
| Finland | No | 47 / 178.8 | 28770 / 91758.2 | -0.3 (-3.0 to 2.3) | 4.8% (-37.3% to 46.9%) |
| Finland | Yes | 0 / 1.4 | 127 / 626.5 | NE | NE |
| Norway | No | 649 / 612.5 | 32573 / 50075.1 | 2.9 (-2.4 to 8.1) | -13.0% (-36.7% to 10.7%) |
| Norway | Yes | <5 / 2.7 | 111 / 181.1 | NE | NE |
| Sweden | No | 10 / 27.2 | 67164 / 184848.6 | -1.2 (-5.8 to 3.3) | 17.9% (-48.3% to 84.0%) |
| Sweden | Yes | <5 / 4.2 | 4250 / 13776.7 | 2.2 (-12.3 to 16.7) | -34.5% (-259.0% to 100%) |

CI denotes confidence interval, CVE comparative vaccine effectiveness, ICU intensive care unit, NE not estimated, PYRS person-years, and RD risk difference. Cell counts lower than 5 could not be reported due to national regulations on data privacy protection. Individuals with a positive PCR test for SARS-CoV-2 within 12 weeks prior to index date were not included.

**Supplementary Table 21. Association between documented SARS-CoV-2 infection and homologous booster schedules as compared with primary schedules and MOD vs BNT in each country according to history of previous SARS-CoV-2 infection.**

| | Previously infected | Studied schedule | Comparison schedule | Measures of association | |
|---|---|---|---|---|---|
| | | Events/PYRS | Events/PYRS | RD per 100 individuals | CVE (95% CI) |
| BNT1BNT2BNT3 vs BNT1BNT2 | | | | | |
| Denmark | No | 96037 / 33872.9 | 76560 / 23389.1 | -0.4 (-1.2 to 0.4) | 0.8% (-1.0% to 2.6%) |
| Denmark | Yes | 4185 / 2357.5 | 4608 / 1914.3 | -4.8 (-7.8 to -1.8) | 13.4% (5.5% to 21.2%) |
| Finland | No | 12673 / 36307.5 | 22812 / 34411.7 | -4.3 (-4.6 to -4.0) | 39.5% (37.2% to 41.8%) |
| Finland | Yes | 53 / 316.6 | 274 / 888.7 | -2.0 (-4.2 to 0.2) | 38.7% (-0.9% to 78.4%) |
| Norway | No | 17285 / 37754.6 | 53369 / 29243.6 | -19.4 (-19.8 to -19.0) | 69.8% (69.2% to 70.5%) |
| Norway | Yes | 34 / 92.3 | 614 / 698.9 | -10.1 (-13.4 to -6.7) | 59.7% (41.9% to 77.5%) |
| Sweden | No | 22450 / 44969.3 | 31505 / 41626.6 | -3.0 (-3.3 to -2.7) | 23.0% (21.3% to 24.8%) |
| Sweden | Yes | 2265 / 6454.7 | 4116 / 6312.2 | -3.8 (-4.6 to -3.0) | 29.1% (23.6% to 34.6%) |
| MOD1MOD2MOD3 vs MOD1MOD2 | | | | | |
| Denmark | No | 23779 / 6992.2 | 21875 / 5198.9 | -2.0 (-3.9 to 0.0) | 4.2% (0.2% to 8.2%) |
| Denmark | Yes | 856 / 412.9 | 978 / 379.1 | -3.6 (-9.1 to 1.9) | 11.2% (-4.8% to 27.3%) |
| Finland | No | 1292 / 4456.6 | 2563 / 4293.3 | -4.6 (-5.5 to -3.7) | 47.9% (41.5% to 54.3%) |
| Finland | Yes | <5 / 29.1 | 15 / 68.2 | -2.1 (-4.6 to 0.4) | 64.4% (9.6% to 100%) |
| Norway | No | 3248 / 5031.9 | 7690 / 3614.8 | -19.8 (-21.1 to -18.4) | 65.1% (62.8% to 67.4%) |
| Norway | Yes | 10 / 17.4 | 133 / 134.2 | -9.8 (-17.6 to -1.9) | 53.9% (17.6% to 90.3%) |
| Sweden | No | 1955 / 5233.8 | 2955 / 4935.9 | -3.1 (-3.8 to -2.4) | 31.0% (25.5% to 36.6%) |
| Sweden | Yes | 202 / 765.5 | 405 / 735.1 | -4.0 (-6.5 to -1.5) | 38.7% (19.7% to 57.6%) |
| MOD1MOD2MOD3 vs BNT1BNT2BNT3 | | | | | |
| Denmark | No | 87100 / 37241.7 | 574850 / 294734.5 | -3.1 (-3.4 to -2.9) | 8.2% (7.6% to 8.8%) |
| Denmark | Yes | 2467 / 1679.9 | 14881 / 15843.7 | 1.0 (0.0 to 2.1) | -4.8% (-9.9% to 0.2%) |
| Finland | No | 3338 / 16225.1 | 36187 / 151925.9 | -0.7 (-0.8 to -0.5) | 16.0% (12.1% to 19.8%) |
| Finland | Yes | <5 / 58.9 | 137 / 726.6 | -2.5 (-3.7 to -1.3) | 76.2% (52.1% to 100%) |
| Norway | No | 10750 / 16258.7 | 61093 / 160714.1 | -0.1 (-0.4 to 0.2) | 0.8% (-1.8% to 3.3%) |
| Norway | Yes | 31 / 97.4 | 179 / 851.3 | 1.4 (-2.3 to 5.1) | -19.9% (-75.9% to 36.1%) |
| Sweden | No | 4615 / 16174.9 | 67317 / 189013.5 | -1.0 (-1.2 to -0.9) | 16.0% (13.5% to 18.5%) |
| Sweden | Yes | 373 / 1413.8 | 4215 / 13771.6 | -0.3 (-1.1 to 0.4) | 5.8% (-6.8% to 18.3%) |

CI denotes confidence interval, CVE comparative vaccine effectiveness, ICU intensive care unit, NE not estimated, PYRS person-years, and RD risk difference. Cell counts lower than 5 could not be reported due to national regulations on data privacy protection. Individuals with a positive PCR test for SARS-CoV-2 within 12 weeks prior to index date were not included.

**Supplementary Table 22. Associated risk of documented SARS-CoV-2 infection comparing previously and non-previously infected within booster schedules.**

| | History of previous infection | | Measures of association | |
|---|---|---|---|---|
| | Yes | No | | |
| | Events/PYRS | Events/PYRS | RD per 100 individuals | CVE (95% CI) |
| AZD1AZD2BNT3 | | | | |
| Denmark | 16 / 6.9 | 229 / 106.7 | 3.4 (-11.5 to 18.4) | -9.5% (-51.0% to 32.0%) |
| Finland | <5 / 42.5 | 986 / 15457.7 | 0.0 (-2.2 to 2.2) | -2.8% (-161.7% to 100%) |
| Norway | <5 / 3.3 | <5 / 92.0 | 3.4 (-19.7 to 26.5) | -37.6% (-293.9% to 100%) |
| Sweden | 862 / 2702.2 | 12843 / 48515.0 | 0.9 (0.4 to 1.4) | -16.0% (-25.1% to -6.8%) |
| AZD1AZD2MOD3 | | | | |
| Denmark | 0 / 0.3 | 8 / 8.3 | NE | NE |
| Finland | 0 / 12.1 | 176 / 5665.3 | NE | NE |
| Norway | 0 / 0.4 | <5 / 29.3 | NE | NE |
| Sweden | 247 / 1099.2 | 3915 / 16218.7 | -0.6 (-1.1 to 0.0) | 11.9% (0.2% to 23.6%) |
| AZD1BNT2BNT3 | | | | |
| Denmark | 1453 / 1002.8 | 23844 / 7275.3 | -22.2 (-23.4 to -21.0) | 47.4% (44.9% to 49.8%) |
| Finland | 14 / 75.4 | 4048 / 14038.0 | -2.0 (-4.0 to 0.0) | 34.6% (-0.5% to 69.7%) |
| Norway | 57 / 165.7 | <5 / 14713.4 | -8.8 (-10.5 to -7.1) | 57.3% (46.2% to 68.3%) |
| Sweden | 1473 / 1430.2 | 10752 / 6894.7 | -7.8 (-8.8 to -6.8) | 29.7% (26.0% to 33.5%) |
| AZD1MOD2MOD3 | | | | |
| Denmark | 491 / 364.0 | 13053 / 4220.7 | -22.7 (-24.5 to -20.9) | 50.8% (46.9% to 54.7%) |
| Finland | <5 / 9.4 | 625 / 2534.1 | 2.7 (-6.9 to 12.3) | -52.6% (-237.8% to 100%) |
| Norway | <5 / 3.7 | <5 / 149.8 | -8.0 (-20.3 to 4.2) | 47.3% (-23.0% to 100%) |
| Sweden | 94 / 112.0 | 846 / 563.9 | -10.0 (-13.6 to -6.4) | 40.2% (26.6% to 53.8%) |
| BNT1BNT2MOD3 | | | | |
| Denmark | 0 / 3.6 | 67 / 68.1 | NE | NE |
| Finland | 20 / 210.0 | 9045 / 41087.3 | NE | NE |
| Norway | 61 / 208.0 | <5 / 37121.6 | -7.5 (-9.0 to -6.0) | 56.6% (45.4% to 67.8%) |
| Sweden | 1344 / 8911.5 | 17858 / 63874.2 | -1.5 (-1.8 to -1.2) | 28.6% (22.6% to 34.6%) |
| MOD1MOD2BNT3 | | | | |
| Denmark | 5 / 5.7 | 215 / 103.1 | -19.5 (-30.3 to -8.8) | 61.6% (29.5% to 93.6%) |
| Finland | 6 / 30.6 | 1531 / 6288.5 | -2.0 (-3.9 to -0.2) | 47.6% (4.6% to 90.5%) |
| Norway | <5 / 97.6 | <5 / 13305.5 | -8.9 (-11.5 to -6.3) | 58.8% (41.5% to 76.0%) |
| Sweden | 624 / 1898.9 | 5743 / 16308.3 | 0.6 (-0.1 to 1.2) | -8.1% (-18.2% to 2.0%) |
| BNT1MOD2MOD3 | | | | |
| Denmark | NE | 10 / 6.4 | NE | NE |
| Finland | 0 / 4.7 | 284 / 793.9 | NE | NE |
| Norway | <5 / 21.5 | 6995 / 7980.7 | -12.7 (-17.2 to -8.2) | 63.3% (41.3% to 85.2%) |
| Sweden | <5 / 8.9 | 12 / 48.2 | 3.0 (-7.8 to 13.7) | NE |
| MOD1BNT2BNT3 | | | | |
| Denmark | 0 / 0.4 | 20 / 11.1 | NE | NE |
| Finland | <5 / 2.4 | 119 / 441.6 | 9.2 (-12.1 to 30.5) | NE |
| Norway | <5 / 4.8 | <5 / 930.8 | NE | NE |
| Sweden | 6 / 23.1 | 41 / 119.4 | 8.8 (-8.6 to 26.3) | NE |
| BNT1MOD2BNT3 | | | | |
| Denmark | NE | <5 / 2.3 | NE | NE |
| Finland | <5 / 5.9 | 305 / 751.1 | NE | NE |
| Norway | <5 / 37.1 | <5 / 12518.7 | 12.7 (-19.1 to 44.5) | -65.1% (-228.2% to 97.9%) |
| Sweden | <5 / 31.7 | 19 / 142.3 | -4.7 (-8.9 to -0.5) | 90.6% (75.0% to 100%) |
| MOD1BNT2MOD3 | | | | |
| Denmark | 0 / 0.2 | 0 / 1.6 | NE | NE |

| | | | | |
|---|---|---|---|---|
| Finland | 0 / 1.4 | 47 / 178.8 | NE | NE |
| Norway | <5 / 2.7 | 649 / 612.5 | NE | NE |
| Sweden | <5 / 5.2 | 10 / 27.2 | 1.4 (-11.3 to 14.2) | -25.2% (-257.2% to 100%) |
| MOD1MOD2MOD3 | | | | |
| Denmark | 2467 / 1679.9 | 87100 / 37241.7 | -12.4 (-13.4 to -11.4) | 35.4% (32.7% to 38.2%) |
| Finland | <5 / 58.9 | 3338 / 16225.1 | -2.8 (-3.6 to -2.0) | 77.8% (56.1% to 99.5%) |
| Norway | <5 / 97.4 | <5 / 16258.7 | -5.0 (-8.4 to -1.7) | 37.8% (12.8% to 62.8%) |
| Sweden | 451 / 1799.5 | 4615 / 16174.9 | -0.1 (-0.7 to 0.5) | 1.6% (-9.9% to 13.1%) |
| BNT1BNT2BNT3 | | | | |
| Denmark | 14881 / 15843.7 | 574850 / 294734.5 | -16.6 (-17.1 to -16.1) | 43.5% (42.1% to 44.8%) |
| Finland | 137 / 726.6 | 36187 / 151925.9 | -0.9 (-1.8 to 0.0) | 21.5% (-0.2% to 43.3%) |
| Norway | <5 / 851.3 | 61093 / 160714.1 | -6.5 (-8.2 to -4.9) | 48.5% (36.2% to 60.8%) |
| Sweden | 5759 / 18832.0 | 67317 / 189013.5 | -0.4 (-0.6 to -0.2) | 6.6% (3.6% to 9.5%) |

CI denotes confidence interval, CVE comparative vaccine effectiveness, ICU intensive care unit, NE not estimated, PYRS person-years, and RD risk difference. Cell counts lower than 5 could not be reported due to national regulations on data privacy protection. Individuals with a positive PCR test for SARS-CoV-2 within 12 weeks prior to index date were not included.

**Supplementary Table 23. Association between COVID-19 outcomes and heterologous booster schedules as compared with homologous booster schedules in each country with use of adjustment for calendar week instead of calendar month.**

| COVID-19 outcome | Studied schedule | Comparison schedule | Measures of association | |
|---|---|---|---|---|
| | Events/PYRS | Events/PYRS | RD per 100 individuals | CVE (95% CI) |
| AZD1AZD2BNT3 vs BNT1BNT2BNT3 | | | | |
| Denmark | 229 / 106.7 | 517302 / 210481.9 | -8.2 (-12.0 to -4.4) | 18.4% (9.8% to 26.9%) |
| Finland | 986 / 15457.7 | 2356 / 47969.2 | -0.1 (-0.3 to 0.1) | 4.1% (-9.9% to 18.2%) |
| Norway | 41 / 92.0 | 68585 / 166278.5 | -5.7 (-8.7 to -2.7) | 38.4% (18.2% to 58.6%) |
| Sweden | 12843 / 48515.0 | 58756 / 152425.3 | 0.6 (0.5 to 0.8) | -12.9% (-15.9% to -9.9%) |
| AZD1AZD2MOD3 vs MOD1MOD2MOD3 | | | | |
| Denmark | 8 / 8.3 | 75144 / 24196.7 | -16.1 (-29.2 to -3.0) | 46.7% (9.0% to 84.4%) |
| Finland | 176 / 5665.3 | 187 / 4729.5 | -0.3 (-0.6 to 0.0) | 29.5% (3.1% to 56.0%) |
| Norway | 17 / 29.3 | 10959 / 16424.1 | -2.8 (-7.7 to 2.1) | 21.5% (-16.4% to 59.5%) |
| Sweden | 3915 / 16218.7 | 3707 / 12528.7 | 0.6 (0.2 to 1.1) | -15.9% (-28.1% to -3.8%) |
| AZD1BNT2BNT3 vs BNT1BNT2BNT3 | | | | |
| Denmark | 23844 / 7275.4 | 414234 / 156233.3 | 1.9 (1.2 to 2.6) | -4.2% (-5.8% to -2.6%) |
| Finland | 4048 / 14038.0 | 30942 / 90440.2 | 0.2 (-0.1 to 0.5) | -3.5% (-9.0% to 2.0%) |
| Norway | 10967 / 14713.4 | 54798 / 94786.2 | -0.1 (-0.5 to 0.3) | 0.9% (-1.7% to 3.5%) |
| Sweden | 10752 / 6894.9 | 63323 / 145842.6 | 4.1 (3.6 to 4.6) | -18.5% (-21.1% to -16.0%) |
| AZD1MOD2MOD3 vs MOD1MOD2MOD3 | | | | |
| Denmark | 13053 / 4220.7 | 54891 / 18868.7 | -1.5 (-7.9 to 4.9) | 3.2% (-10.3% to 16.6%) |
| Finland | 625 / 2534.1 | 2871 / 11136.5 | 1.4 (0.8 to 2.1) | -38.7% (-60.2% to -17.2%) |
| Norway | 119 / 149.9 | 10078 / 12448.1 | 1.2 (-2.4 to 4.7) | -7.6% (-30.5% to 15.4%) |
| Sweden | 846 / 563.9 | 4155 / 11813.5 | 5.1 (3.2 to 7.1) | -26.0% (-36.8% to -15.2%) |
| BNT1BNT2MOD3 vs BNT1BNT2BNT3 | | | | |
| Denmark | 67 / 68.1 | 580653 / 285991.3 | -19.5 (-23.5 to -15.4) | 53.3% (42.2% to 64.4%) |
| Finland | 9045 / 41087.3 | 35133 / 136227.2 | -1.4 (-2.1 to -0.7) | 23.7% (12.1% to 35.2%) |
| Norway | 22755 / 37011.0 | 60448 / 164700.4 | 1.6 (1.4 to 1.8) | -13.9% (-15.9% to -11.8%) |
| Sweden | 17858 / 63874.3 | 66697 / 186663.1 | -0.8 (-0.9 to -0.7) | 12.8% (11.1% to 14.4%) |
| MOD1MOD2BNT3 vs BNT1BNT2BNT3 | | | | |
| Denmark | 215 / 103.1 | 586998 / 304529.9 | -4.7 (-8.6 to -0.8) | 12.9% (2.3% to 23.6%) |
| Finland | 1531 / 6288.5 | 37728 / 154405.4 | -0.4 (-0.7 to -0.1) | 8.7% (2.4% to 15.1%) |
| Norway | 10124 / 13306.3 | 70657 / 190310.5 | 1.2 (0.9 to 1.6) | -8.7% (-11.2% to -6.1%) |
| Sweden | 5744 / 16309.7 | 74728 / 202021.2 | -0.1 (-0.2 to 0.1) | 0.8% (-1.9% to 3.4%) |
| BNT1MOD2MOD3 vs BNT1BNT2BNT3 | | | | |
| Denmark | 10 / 6.4 | 573968 / 297012.1 | -5.8 (-18.2 to 6.6) | 20.0% (-22.5% to 62.4%) |
| Finland | 284 / 793.9 | 28741 / 89021.6 | -1.4 (-2.8 to 0.0) | 19.9% (0.9% to 38.9%) |
| Norway | 6995 / 7980.9 | 30755 / 47764.3 | 2.9 (1.9 to 4.0) | -17.1% (-23.3% to -10.9%) |
| Sweden | 12 / 48.2 | 66067 / 177793.9 | -3.4 (-5.1 to -1.7) | 56.4% (27.9% to 85.0%) |
| MOD1BNT2BNT3 vs BNT1BNT2BNT3 | | | | |
| Denmark | 20 / 11.1 | 595378 / 306951.1 | 4.1 (-7.6 to 15.8) | -15.1% (-57.8% to 27.7%) |
| Finland | 119 / 441.6 | 37126 / 146707.4 | 1.1 (-1.1 to 3.2) | -19.9% (-61.0% to 21.2%) |
| Norway | 1064 / 930.9 | 38236 / 55002.8 | 5.9 (0.0 to 11.8) | -25.4% (-50.9% to 0.1%) |
| Sweden | 41 / 119.7 | 74260 / 200193.9 | 1.7 (-1.4 to 4.9) | -24.8% (-69.5% to 19.9%) |

| COVID-19 outcome | Studied schedule<br>Events/PYRS | Comparison schedule<br>Events/PYRS | Measures of association<br>RD per 100 individuals | CVE (95% CI) |
|---|---|---|---|---|
| BNT1MOD2BNT3 vs BNT1BNT2BNT3 | | | | |
| Denmark | <5 / 2.3 | 584116 / 294224.6 | -18.3 (-43.8 to 7.3) | 44.0% (-17.4% to 100%) |
| Finland | 305 / 751.1 | 31999 / 101003.0 | -2.3 (-3.5 to -1.2) | 29.0% (15.4% to 42.7%) |
| Norway | 10660 / 12518.7 | 32303 / 43862.6 | 1.3 (-0.2 to 2.7) | -7.1% (-15.1% to 1.0%) |
| Sweden | 19 / 142.3 | 52427 / 123389.4 | -1.1 (-5.2 to 3.1) | 16.8% (-49.7% to 83.4%) |
| MOD1BNT2MOD3 vs BNT1BNT2BNT3 | | | | |
| Denmark | 0 / 1.6 | 564709 / 296644.2 | | |
| Finland | 47 / 178.8 | 28770 / 91758.2 | -0.3 (-3.0 to 2.3) | 5.2% (-36.7% to 47.1%) |
| Norway | 649 / 612.5 | 32573 / 50075.2 | 2.5 (-2.7 to 7.7) | -11.1% (-34.4% to 12.2%) |
| Sweden | 10 / 27.2 | 67166 / 184851.6 | -1.0 (-5.5 to 3.6) | 14.5% (-54.3% to 83.3%) |
| MOD1MOD2MOD3 vs. BNT1BNT2BNT3 | | | | |
| Denmark | 87100 / 37241.8 | 574850 / 294734.5 | -3.1 (-3.3 to -2.8) | 8.0% (7.4% to 8.7%) |
| Finland | 3338 / 16225.1 | 36187 / 151925.9 | -0.7 (-0.8 to -0.5) | 16.0% (12.2% to 19.9%) |
| Norway | 10750 / 16259.1 | 61093 / 160717.6 | 0.0 (-0.4 to 0.3) | 0.1% (-2.5% to 2.7%) |
| Sweden | 4615 / 16175.2 | 67320 / 189016.9 | -0.8 (-0.9 to -0.6) | 12.4% (9.8% to 15.0%) |

CI denotes confidence interval, CVE comparative vaccine effectiveness, ICU intensive care unit, NE not estimated, PYRS person-years, and RD risk difference. Cell counts lower than 5 could not be reported due to national regulations on data privacy protection.

**Supplementary Figure 25. Cumulative incidence curves of documented SARS-CoV-2 infection comparing booster and matched primary schedules in Denmark stratified by calendar periods.**

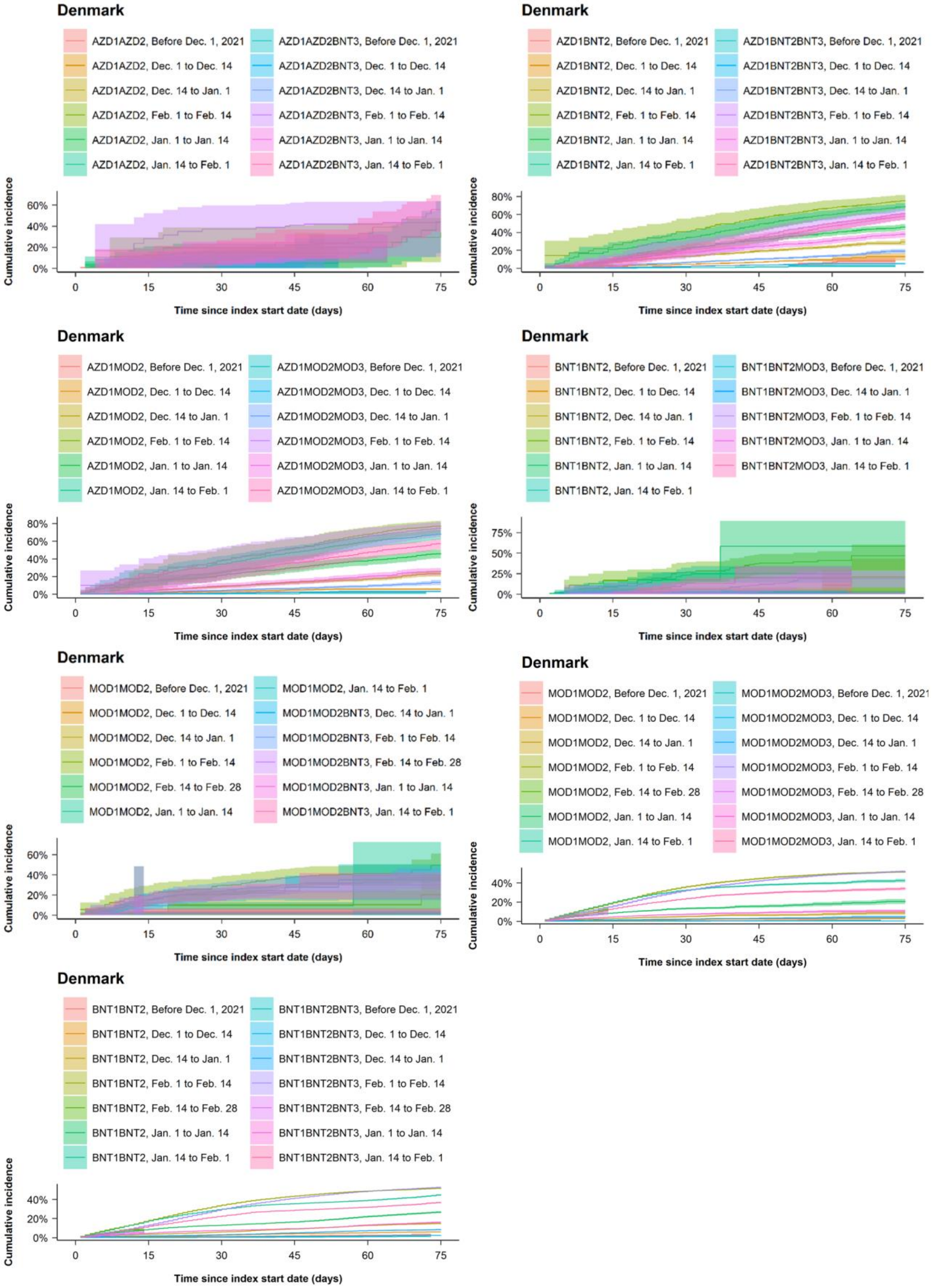

**Supplementary Table 24. Association between documented SARS-CoV-2 infection and heterologous booster schedules as compared with matched primary schedules in Denmark stratified by calendar periods.**

| | Studied schedule | Comparison schedule | Measures of association | |
|---|---|---|---|---|
| | Events/PYRS | Events/PYRS | RD per 100 individuals | CVE (95% CI) |
| AZD1AZD2BNT3 vs AZD1AZD2 | | | | |
| Dec. 1 to Dec. 14 | NE | NE | NE | NE |
| Dec. 14 to Jan. 1 | NE | NE | NE | NE |
| Jan. 1 to Jan. 14 | NE | NE | NE | NE |
| Jan. 14 to Feb. 1 | 20 / 6.1 | 9 / 5.8 | 11.5 (-46.1 to 69.2) | -26.1% (-176.4% to 100%) |
| Feb. 1 to Feb. 14 | 8 / 3.5 | 6 / 3.7 | 4.2 (-68.0 to 76.5) | -9.7% (-183.6% to 100%) |
| Feb. 14 to Feb. 28 | <5 / 3.2 | <5 / 3.5 | 15.5 (-197.0 to 100) | -28.5% (-471.4% to 100%) |
| AZD1BNT2BNT3 vs AZD1BNT2 | | | | |
| Dec. 1 to Dec. 14 | 197 / 702.7 | 410 / 684.5 | -8.0 (-12.9 to -3.0) | 61.1% (44.5% to 77.8%) |
| Dec. 14 to Jan. 1 | 388 / 418.8 | 646 / 376.2 | -10.1 (-15.6 to -4.7) | 34.5% (20.0% to 49.0%) |
| Jan. 1 to Jan. 14 | 378 / 165.8 | 421 / 131.3 | -8.1 (-16.7 to 0.5) | 17.5% (0.7% to 34.2%) |
| Jan. 14 to Feb. 1 | 613 / 123.0 | 495 / 83.5 | -10.6 (-27.7 to 6.4) | 15.3% (-6.7% to 37.4%) |
| Feb. 1 to Feb. 14 | 300 / 47.0 | 220 / 30.3 | -12.2 (-52.0 to 27.6) | 16.0% (-29.8% to 61.7%) |
| Feb. 14 to Feb. 28 | 108 / 23.4 | 90 / 14.8 | -20.1 (-71.2 to 31.1) | 28.1% (-31.0% to 87.2%) |
| AZD1MOD2MOD3 vs AZD1MOD2 | | | | |
| Dec. 1 to Dec. 14 | 62 / 381.4 | 148 / 373.4 | -2.6 (-4.2 to -1.1) | 45.8% (24.7% to 67.0%) |
| Dec. 14 to Jan. 1 | 157 / 263.8 | 295 / 245.7 | -10.1 (-15.6 to -4.5) | 43.1% (24.8% to 61.3%) |
| Jan. 1 to Jan. 14 | 150 / 102.5 | 246 / 86.2 | -19.8 (-30.5 to -9.0) | 42.6% (26.1% to 59.2%) |
| Jan. 14 to Feb. 1 | 334 / 75.2 | 315 / 54.3 | -11.5 (-33.6 to 10.7) | 16.7% (-12.4% to 45.7%) |
| Feb. 1 to Feb. 14 | 219 / 28.4 | 168 / 18.9 | -3.8 (-55.1 to 47.4) | 4.8% (-57.4% to 66.9%) |
| Feb. 14 to Feb. 28 | 75 / 11.3 | 46 / 7.1 | -2.7 (-61.3 to 55.8) | 4.2% (-82.5% to 90.9%) |
| BNT1BNT2MOD3 vs BNT1BNT2 | | | | |
| Dec. 1 to Dec. 14 | 0 / 0.6 | <5 / 1.0 | NE | NE |
| Dec. 14 to Jan. 1 | <5 / 2.4 | <5 / 2.5 | -2.4 (-7.9 to 3.1) | 71.7% (1.0% to 100%) |
| Jan. 1 to Jan. 14 | <5 / 5.3 | 14 / 5.1 | -57.8 (-197.0 to 81.4) | 98.7% (94.7% to 100%) |
| Jan. 14 to Feb. 1 | | | NE | NE |
| Feb. 1 to Feb. 14 | 6 / 5.4 | 17 / 4.7 | -21.0 (-52.4 to 10.5) | 52.8% (-4.6% to 100%) |
| Feb. 14 to Feb. 28 | 7 / 5.6 | 12 / 4.6 | NE | NE |
| MOD1MOD2BNT3 vs MOD1MOD2 | | | | |
| Dec. 1 to Dec. 14 | 0 / 2.5 | <5 / 2.4 | NE | NE |
| Dec. 14 to Jan. 1 | <5 / 4.9 | 6 / 4.6 | -19.1 (-48.0 to 9.7) | 94.4% (80.8% to 100%) |
| Jan. 1 to Jan. 14 | 7 / 9.6 | 20 / 9.0 | -35.9 (-116.1 to 44.3) | 90.9% (71.1% to 100%) |
| Jan. 14 to Feb. 1 | 36 / 13.8 | 33 / 12.4 | -5.0 (-37.5 to 27.5) | 14.2% (-69.8% to 98.2%) |
| Feb. 1 to Feb. 14 | 25 / 10.0 | 33 / 8.7 | -25.8 (-81.5 to 29.9) | 44.0% (-13.9% to 100%) |
| Feb. 14 to Feb. 28 | 19 / 9.8 | 18 / 8.4 | -4.4 (-42.0 to 33.1) | 10.8% (-74.2% to 95.8%) |
| BNT1BNT2BNT3 vs BNT1BNT2 | | | | |
| Dec. 1 to Dec. 14 | 366 / 4224.4 | 1262 / 4112.4 | -3.7 (-4.3 to -3.1) | 60.3% (53.8% to 66.9%) |
| Dec. 14 to Jan. 1 | 2624 / 4674.7 | 4590 / 4448.2 | -6.4 (-7.6 to -5.3) | 42.6% (37.1% to 48.2%) |
| Jan. 1 to Jan. 14 | 7203 / 6436.4 | 12595 / 6062.4 | -10.9 (-12.6 to -9.2) | 40.2% (35.4% to 45.0%) |
| Jan. 14 to Feb. 1 | 25409 / 8563.9 | 30713 / 7541.4 | -7.8 (-9.7 to -6.0) | 17.4% (13.7% to 21.1%) |
| Feb. 1 to Feb. 14 | 23225 / 4959.7 | 19568 / 4150.2 | -0.2 (-2.5 to 2.1) | 0.4% (-3.8% to 4.6%) |
| Feb. 14 to Feb. 28 | 15316 / 4281.8 | 10562 / 3608.9 | 4.1 (1.5 to 6.7) | -9.4% (-15.5% to -3.2%) |
| MOD1MOD2MOD3 vs MOD1MOD2 | | | | |
| Dec. 1 to Dec. 14 | 12 / 369.5 | 73 / 362.9 | -2.5 (-3.7 to -1.4) | 85.7% (74.1% to 97.3%) |
| Dec. 14 to Jan. 1 | 190 / 498.3 | 338 / 480.8 | -3.5 (-6.4 to -0.7) | 41.1% (14.4% to 67.7%) |
| Jan. 1 to Jan. 14 | 851 / 777.1 | 1587 / 744.1 | -10.4 (-14.4 to -6.4) | 49.1% (36.8% to 61.4%) |
| Jan. 14 to Feb. 1 | 6895 / 2148.8 | 9622 / 1952.2 | -8.1 (-12.1 to -4.2) | 19.1% (10.8% to 27.4%) |
| Feb. 1 to Feb. 14 | 7124 / 1460.0 | 6757 / 1232.4 | -2.7 (-7.5 to 2.1) | 5.0% (-3.6% to 13.5%) |
| Feb. 14 to Feb. 28 | 4853 / 1282.0 | 3735 / 1066.2 | 3.5 (-1.0 to 8.0) | -7.5% (-17.7% to 2.6%) |

CI denotes confidence interval, CVE comparative vaccine effectiveness, ICU intensive care unit, NE not estimated, PYRS person-years, and RD risk difference. Cell counts lower than 3 in Denmark could not be reported due to national regulations on data privacy protection. The comparisons of AZD1AZD2MOD3 vs AZD1AZD2, BNT1MOD2MOD3 vs BNT1MOD2, MOD1BNT2BNT3 vs MOD1BNT2, BNT1MOD2BNT3 vs BNT1MOD2, MOD1BNT2MOD3 vs MOD1BNT2, and BNT1BNT2BNT3 vs BNT1BNT2 could not be stratified according to calendar periods due to too few individuals.

**Supplementary References.**

1 Schmidt M, Pedersen L, Sørensen HT. The Danish Civil Registration System as a tool in epidemiology. *Eur J Epidemiol* 2014; **29**: 541–9.

2 Krause TG, Jakobsen S, Haarh M, Mølbak K. The Danish vaccination register. *Euro Surveill* 2012; **17**: 20155.

3 Voldstedlund M, Haarh M, Mølbak K, MiBa Board of Representatives. The Danish Microbiology Database (MiBa) 2010 to 2013. *Euro Surveill* 2014; **19**: 20667.

4 Schmidt M, Schmidt SAJ, Sandegaard JL, Ehrenstein V, Pedersen L, Sørensen HT. The Danish National Patient Registry: a review of content, data quality, and research potential. *Clinical Epidemiology* 2015; : 449.

5 Population Information System | Digital and population data services agency. Digi- ja väestötietovirasto. https://dvv.fi/en/population-information-system (accessed March 20, 2022).

6 Register of Social assistance - THL. Finnish Institute for Health and Welfare (THL), Finland. https://thl.fi/en/web/thlfi-en/statistics-and-data/data-and-services/register-descriptions/social-assistance (accessed March 20, 2022).

7 Terhikki Register - valvira englanti. Terhikki Register. http://www.valvira.fi/web/en/healthcare/professional_practice_rights/terhikki_register (accessed March 20, 2022).

8 Baum U, Sundman J, Jääskeläinen S, Nohynek H, Puumalainen T, Jokinen J. Establishing and maintaining the National Vaccination Register in Finland. *Euro Surveill* 2017; **22**: 30520.

9 Finnish National Infectious Diseases Register - THL. Finnish Institute for Health and Welfare (THL), Finland. https://thl.fi/en/web/infectious-diseases-and-vaccinations/surveillance-and-registers/finnish-national-infectious-diseases-register (accessed March 20, 2022).

10 Care Register for Health Care - THL. Finnish Institute for Health and Welfare (THL), Finland. https://thl.fi/en/web/thlfi-en/statistics-and-data/data-and-services/register-descriptions/care-register-for-health-care (accessed March 20, 2022).

11 Register of Primary Health Care visits - THL. Finnish Institute for Health and Welfare (THL), Finland. https://thl.fi/en/web/thlfi-en/statistics-and-data/data-and-services/register-descriptions/register-of-primary-health-care-visits (accessed March 29, 2022).

12 Lindman AES. Emergency preparedness register for COVID-19 (Beredt C19). Norwegian Institute of Public Health. https://www.fhi.no/en/id/infectious-diseases/coronavirus/emergency-preparedness-register-for-covid-19/ (accessed March 20, 2022).

13 State Register of Employers and Employees (Aa-registeret). nav.no. https://www.nav.no/en/home/employers/nav-state-register-of-employers-and-employees (accessed March 20, 2022).

14 Iplos-registeret. Helsedirektoratet. https://www.helsedirektoratet.no/tema/statistikk-registre-og-rapporter/helsedata-og-helseregistre/iplos-registeret (accessed March 20, 2022).

15 Trogstad L, Ung G, Hagerup-Jenssen M, Cappelen I, Haugen IL, Feiring B. The Norwegian immunisation register--SYSVAK. *Euro Surveill* 2012; **17**: 20147.

16 Bakken IJ, Ariansen AMS, Knudsen GP, Johansen KI, Vollset SE. The Norwegian Patient Registry and the Norwegian Registry for Primary Health Care: Research potential of two nationwide health-care registries. *Scand J Public Health* 2020; **48**: 49–55.

17 Registrering i Norsk pandemiregister – informasjon til ansatte. Helse Bergen. https://helse-bergen.no/norsk-pandemiregister/registrering-i-norsk-pandemiregister-informasjon-til-ansatte (accessed Oct 16, 2022).

18 Ludvigsson JF, Almqvist C, Bonamy A-KE, *et al.* Registers of the Swedish total population and their use in medical research. *Eur J Epidemiol* 2016; **31**: 125–36.

19 Brooke HL, Talbäck M, Hörnblad J, *et al.* The Swedish cause of death register. *Eur J Epidemiol* 2017; **32**: 765–73.

20 Ludvigsson JF, Svedberg P, Olén O, Bruze G, Neovius M. The longitudinal integrated database for health insurance and labour market studies (LISA) and its use in medical research. *Eur J Epidemiol* 2019; **34**: 423–37.

21 Registret över insatser till äldre och personer med funktionsnedsättning. Socialstyrelsen. https://www.socialstyrelsen.se/statistik-och-data/register/aldre-och-personer-med-funktionsnedsattning/ (accessed March 20, 2022).

22 Chrapkowska C, Galanis I, Kark M, *et al.* Validation of the new Swedish vaccination register - Accuracy and completeness of register data. *Vaccine* 2020; **38**: 4104–10.

23 Rolfhamre P, Jansson A, Arneborn M, Ekdahl K. SmiNet-2: Description of an internet-based surveillance system for communicable diseases in Sweden. *Euro Surveill* 2006; **11**: 103–7.

24 National Patient Register. Socialstyrelsen. https://www.socialstyrelsen.se/en/statistics-and-data/registers/national-patient-register/ (accessed March 20, 2022).

25 Ludvigsson JF, Andersson E, Ekbom A, *et al.* External review and validation of the Swedish national inpatient register. *BMC Public Health* 2011; **11**: 450.
